# A dynamic transmission model for assessing the impact of pneumococcal vaccination in the United States

**DOI:** 10.1101/2024.06.11.24308671

**Authors:** Tufail M Malik, Kevin M Bakker, Rachel J Oidtman, Oluwaseun Sharomi, Giulio Meleleo, Robert B Nachbar, Elamin H Elbasha

**Author notes:** Corresponding author (TM).

## Abstract

*Streptococcus pneumoniae* (SP) is a bacterial pathogen that kills more than 300,000 children every year across the globe. Multiple vaccines exist that prevent pneumococcal disease, with each vaccine covering a variable number of the more than 100 known serotypes. Due to the high effectiveness of these vaccines, each new pneumococcal conjugate vaccine (PCV) introduction has resulted in a decrease in vaccine-type disease and a shift in the serotype distribution towards non-vaccine types in a phenomenon called serotype replacement. Here, an age-structured compartmental model was created that reproduced historical carriage transmission dynamics in the United States and was used to evaluate the population-level impact of new vaccine introductions into the pediatric population. The model incorporates co-colonization and serotype competition, which drives replacement of the vaccine types by the non-vaccine types. The model was calibrated to historical age- and serotype-specific invasive pneumococcal disease (IPD) data from the United States. Vaccine-specific coverage and effectiveness were integrated in accordance with the recommended timelines for each age group. Demographic parameters were derived from US-population-specific databases, while population mixing patterns were informed by US-specific published literature on age-group based mixing matrices. The calibrated model was then used to project the epidemiological impact of PCV15, a 15-valent pneumococcal vaccine, compared with the status quo vaccination with PCV13 and demonstrated the value of added serotypes in PCV15. Projections revealed that PCV15 would reduce IPD incidence by 6.04% (range: 6.01% to 6.06%) over 10 years when compared to PCV13.

## Introduction

Despite the widespread use of pneumococcal vaccines in the United States (US), *Streptococcus pneumoniae* (SP) caused nearly 17,400 cases of invasive pneumococcal disease (IPD) in 2021, and nearly 3,000 associated deaths [1]. Globally the number of deaths in children under 5 is approximately 300,000 [2].

There are more than 100 SP serotypes (STs) [3] . Only a small fraction of these cause majority of disease which may vary in frequency and severity by age. Vaccines are only able to target a portion of pneumococcal STs, and their effectiveness is serotype-specific. Asymptomatic or symptomatic nasopharyngeal carriage (NPC) of SP in an individual, which is relatively common in young children [5], may result in SP transmission to other individuals through person-to-person contact via respiratory droplets. NPC can lead to multiple disease outcomes, including invasive pneumococcal disease (IPD), non-bacteremic pneumococcal pneumonia (NBPP), and acute otitis media (AOM) [6]. IPD, which includes meningitis and bacteremic pneumonia, primarily afflicts extremes of ages and those with risk factors [3]. AOM is a condition that exclusively affects children. Vaccines play a role in preventing NPC, but the necessary immune response is higher than what is required to prevent disease.

In the US a pediatric vaccination program with a 7-valent pneumococcal conjugate vaccine (PCV) was introduced in 2000, and building on this composition, a 13-valent PCV was introduced in 2010, followed by a 15-valent vaccine in 2022, and a 20-valent vaccine in 2023. Since 1983, a 23-valent pneumococcal polysaccharide vaccine (PPSV23) had been recommended for all adults 65 years or older. Vaccines targeting additional or different serotypes, along with those based on other technologies, are currently in development. Since the introduction of PCVs, the US has seen a dramatic reduction in IPD across all age groups in vaccine-type (VT) serotypes [3]. There has also been replacement of VTs with non-vaccine types (NVTs) [7]. Globally, there have been higher levels of replacement than those seen in the US, sometimes leading to higher overall incidence levels than the pre-vaccine period (for example in France following PCV13 introduction [8]).

Dynamic transmission models (DTMs) have been used extensively to describe the epidemiological and biological dynamics of SP transmission in the presence or absence of vaccination (Table S1 summarizes a review of published models). In this study, a DTM was developed to quantify the epidemiological impact of ST-specific and age-based vaccination programs. The model is driven by carriage transmission and assigns each ST into one of the serotype classes (STCs), which were determined based on their inclusion in different pneumococcal vaccines. Additionally, the model features indirect effects such as herd immunity and competition between ST classes, which determines replacement dynamics following vaccine introduction. The model is flexible, so that it can be used in countries that report disease incidence with different age-groups, incorporated different vaccines into their vaccine schedules (e.g., PCV10), want to examine different vaccination policies, or are interested in other pneumococcal diseases of interest. Here, the model was applied to the US setting by dividing STs into 11 STCs (outlined in Table 1) and dividing the population into 6 age groups (listed in Table 2). The model was fitted to historical (2000–2019) US IPD incidence (per 100,000) data, by incorporating each pneumococcal vaccine in the model according to its time of inclusion in US immunization programs in respective age groups, and the epidemiological impact of the continued use of PCV13 versus implementing PCV15 in the pediatric immunization program was assessed (at the time of this study PCV20 was not yet recommended in peds in the US).

**Table 1.**
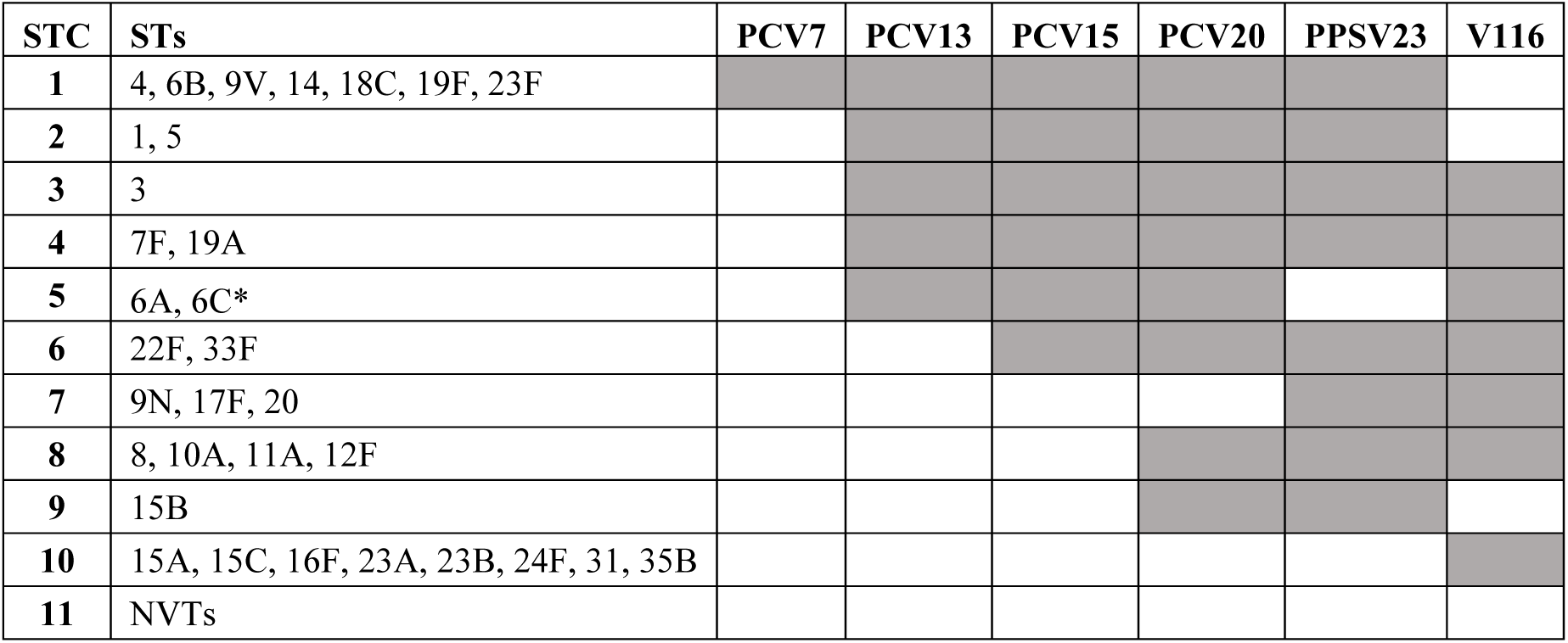
Model serotype classes (STCs). *: cross protection from 6A antigen.

**Table 2.**
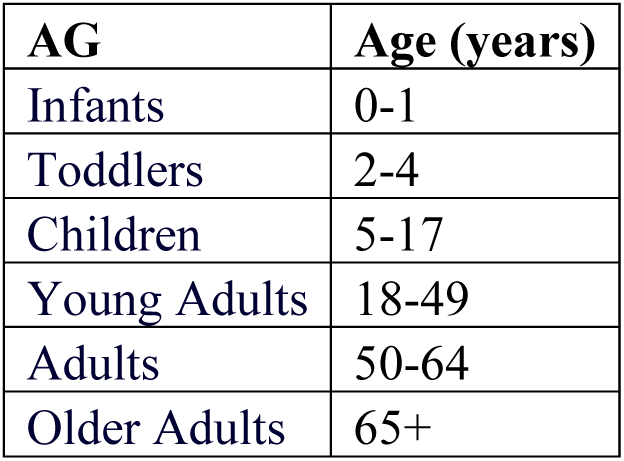
Age groups (AG) used in the model.

## Methods

A deterministic age-structured DTM was developed that described SP carriage transmission dynamics and disease progression in the presence of age- and serotype-specific pneumococcal vaccines. In addition to the brief description of the model here, additional details can be found in the Supplementary Appendix. Real-world data were used to inform parameters for vaccine effectiveness against disease, coverage rates, duration of protection, and carriage clearance rates. Published contact matrices [9] were used with suitable adaptation for model age structure to inform contact matrix within and between age groups. Real-world data are lacking for PCV15 given recent introduction. Population data were used to parameterize the demographic aspects of the model. The model was calibrated to national-level IPD data in the US to estimate the probability of SP carriage acquisition per contact, competition between STCs, progression rate from carriage to disease, and vaccine effectiveness against carriage. Disease incidence data were stratified by age and grouped into STCs, based on inclusion in various vaccines (Table 1).

Previous studies have employed a similar approach of grouping STs into classes and have embraced similar assumptions [10, 11, 12, 13] (see Section 3.2.1 of the Supplementary Appendix for detail). With the calibrated model, epidemiological projections were made under different pediatric vaccination scenarios keeping a realistic mix of vaccination regimen in the adult population.

### Model overview

The model was based on a system of ordinary differential equations and distinguished pneumococcal carriage from disease as well as their vaccine protection status, which varied by age and vaccine type (Fig 1). For example, an unvaccinated person can be either susceptible non-colonized or colonized with up to three STCs. A fraction of infants is born into the unvaccinated susceptible non-colonized compartment and remain there until they are vaccinated, become colonized, or die from causes other than pneumococcal disease. Individuals acquire one or two STCs simultaneously at an age-specific and time-dependent rate (i.e., per capita force of carriage acquisition), progressing from non-colonized status to single or double colonization, from single to double or triple colonization, and from double to triple colonization. The model assumes simultaneous acquisition of up to two STCs because the acquisition of multiple STCs is exceedingly unlikely. It was assumed that an individual could be colonized by up to three STCs concurrently.

**Fig 1.**
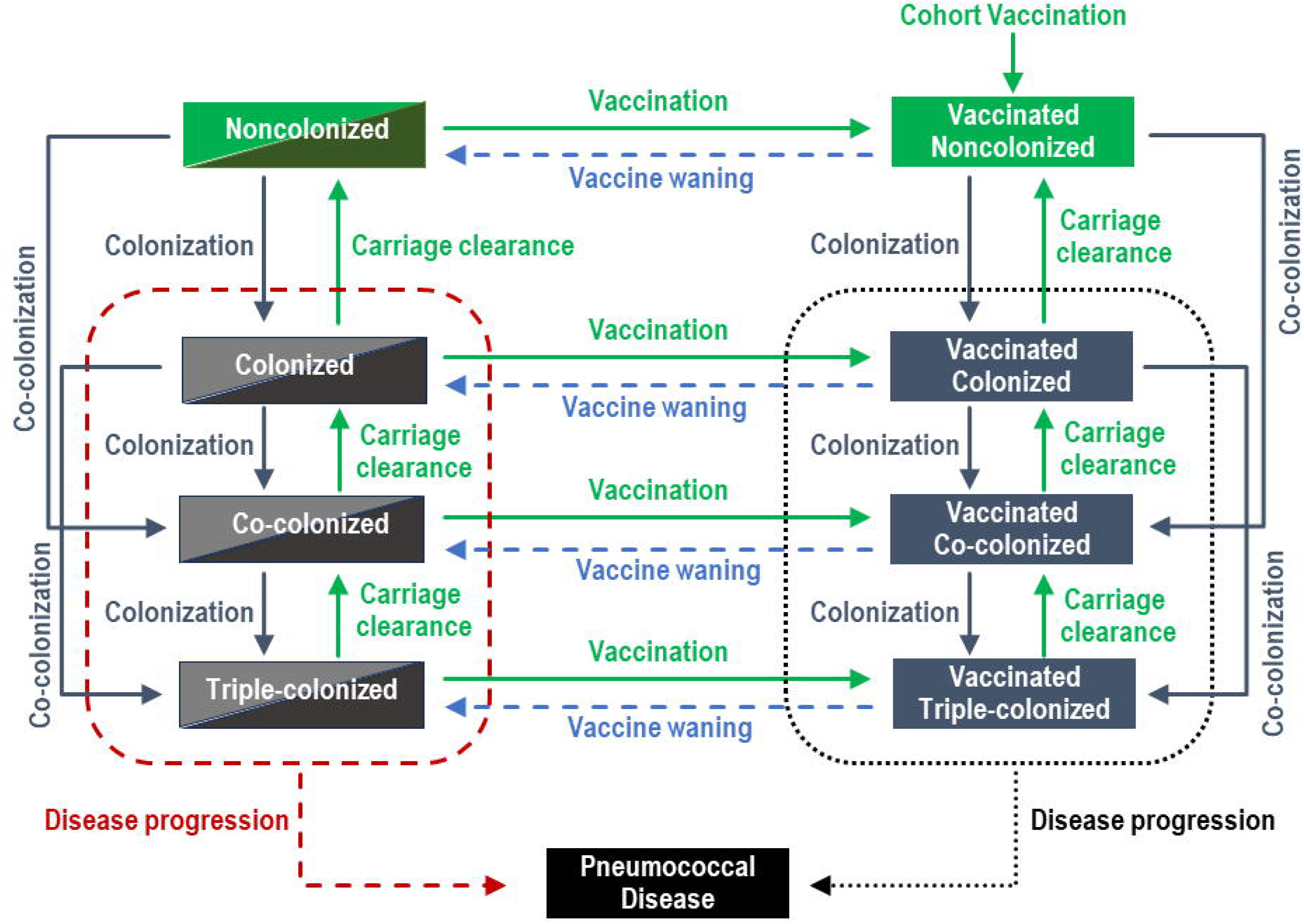
Model flowchart. Individuals are born into the non-colonized compartments and can move throughout the system during their lifetime. Upon carriage acquisition, individuals move to the colonized (with a single ST), co-colonized with two STs, or triple-colonized compartments. Colonized individuals can clear carriage of one ST at a time. A proportion of colonized individuals will develop a pneumococcal disease. Vaccination (right arm of flowchart) reduces the chance of carriage acquisition and disease development for vaccine STCs. Vaccine can wane over time, but vaccination history is tracked. Compartments in the left arm of the flowchart are composed of vaccine-naïve and vaccine-waned individuals. Detailed model flowchart is provided in Figure S3-S5.

The rate at which uncolonized persons of a given age group acquire SP and become colonized depends on the number and type of contacts, the fraction of colonized, co-colonized, and triple- colonized contacts, and the transmission probability per contact, which varies by age. The number of contacts are governed by a mixing matrix, where each cell represents the number of contacts a person of a given age class makes with a person in another age class (Table S4). Upon recovery, it is assumed that the person does not develop immunity and becomes completely susceptible again.

Individuals leave the unvaccinated compartments through cohort and continuous vaccination at given rates and enter the appropriate vaccinated compartment, based on their colonization status. They remain in a given vaccinated compartment until they die, acquire an additional STC, or their immunity wanes. Since typical colonization is asymptomatic, individuals can be vaccinated from any of the four (uncolonized, colonized, double colonized, or triple-colonized) unvaccinated compartments. Vaccination reduces the risk of carriage acquisition and disease progression.

In the DTM, the rate of acquisition of a second or third STC is reduced due to competition from the currently carried STC(s). The competition parameter is determined by the relationship between the current and the invading STC, and can range from no competition (i.e., independence) to complete competition (i.e., exclusion), where the acquisition of the invading STC is completely blocked by the currently carried STC(s).

Disease occurrence was modeled using a case-carrier ratio that varies by disease manifestation, age, STC, and vaccine-status. Progression to IPD, AOM, or NBPP was included here, while other manifestations (such as sinusitis) or further classification (such as bacteremia) could be included in future analyses.

### Model application to the US

Individual serotypes were grouped into 11 serotype classes (STCs), based on inclusion in various current, past, and future vaccines, including PCV7, PCV13, PCV15, PCV20, PCV21 (V116), and PPSV23 (Table 1).

The classification is based on whether a ST is targeted by an individual vaccine so that the assessment of each vaccine can be carried out by grouping some of the classes. For example, PPSV23 includes STCs 1-4 and STCs 6-9, while V116 includes STCs 3-8 and STC 10. Corresponding parameters were estimated according to this age and ST grouping.

It is worth noting that the current model offers flexibility in the aggregation of serotypes into classes. In particular, it includes a broader disaggregation of STs than those present in literature. Table S1 summarizes the serotype classifications or serogroupings used in published pneumococcal models.

Each STC was further categorized into 6 age groups described in Table 2.

This age grouping is chosen based on the availability of historic IPD incidence data in the US, which was used as the calibration target. Additionally, these age groups align with vaccination policy recommendations by the Advisory Committee on Immunization Practices (ACIP) of the Centers for Disease Control and Prevention (CDC). For instance, pediatric vaccination historically targets infants, adult vaccination covers older adults, and at-risk groups include young adults and adults. Future policy considerations related to adult vaccination may involve expanding age-based vaccination to include adults and older adults.

In addition to the scenarios evaluated here, the model is flexible and can evaluate other vaccine scenarios, such as reducing the vaccination age from older adults to adults, having sequential vaccination of different vaccines in an age group, other vaccines, or other potential policy questions.

Numerical computations, including model calibration and projections, were implemented using Mathematica® 14.0 (https://reference.wolfram.com/mathematica, https://reference.wolfram.com/language/ref/NDSolve.html, https://reference.wolfram.com/language/ref/NIntegrate.html, https://reference.wolfram.com/language/ref/NMinimize.html).

### Model parameters and data sources

Baseline values for demographic, epidemiologic, and vaccine inputs of the model were identified through a targeted search of the published literature and public databases.

### Demographic parameters

A stable equilibrium age distribution (with 2017 as the baseline year) and constant mortality hazards for each age group were assumed, using data from the US Census Bureau website (https://www.census.gov/). Since the implied growth rate was insignificant, constant population was assumed over the analysis time horizon to reduce model complexity.

### Contact mixing matrix

Mixing patterns among the population affect the transmission dynamics of an infectious pathogen. Mixing patterns from Prem *et al*. [9] were adapted to create the 6-age group contact matrix (see Section 3.1.3 of the Supplementary Appendix). Although transmission among adult age groups was allowed, it was assumed that there was no transmission of SP from adults to the pediatric population due to limited evidence of adult-to-child transmission and vast evidence that child-to-child transmission drives SP dynamics [14, 15, 16].

### Vaccine parameters

The model assumed full vaccine compliance (i.e., 3+1 doses in pediatric population) and used the respective age-specific observed vaccine coverage rates (VCRs) over the historical period and anticipated VCRs for projections. VCR in each vaccine-target age group is discussed in Supplementary Appendix. In the model, a distinction between two types of vaccine effectiveness (VE) was made: VEc against carriage acquisition and VEd against pneumococcal disease (VEd was unique for each disease outcome of IPD, AOM, and NBPP). VEs vary by vaccine (PCV13, PCV15, PCV20, and PPSV23), STC, and age group. VEds against NBPP and AOM can be seen in Supplementary Table S10 and S11. VEcs were calibrated in the model.

STC- and age-specific VEds against IPD of all vaccines included in the model were based on clinical trials and real-world-evidence studies (Table 3). VEd of PCV13 against STC1-type (i.e., PCV7 STs) IPD in children was assumed to be identical to published PCV7 VEd [17]. ST- specific VEds for the additional PCV13 serotypes were taken from Moore *et al*. [18]. Since VEds within an STC may vary, VEds for each STC in children were estimated as the incidence- weighted average of individual VEds. There is no consensus regarding the VEd of PCV13 on ST3 [19] (which constitutes STC3). Despite high pediatric coverage levels, real world evidence reveals that the long-term impact of PCV13 has had little to no effect on population-level ST3 incidence. Therefore, VEd against STC3 in the base case was assumed to be equal to the left end point of the 95% confidence interval from Moore *et al*. [18].

**Table 3.** Vaccine effectiveness against IPD by age and serotype class. (PCV20 was only considered for adult vaccination).

|  | STC | Pediatric age groups | Young adults | Adults | Older adults |
| --- | --- | --- | --- | --- | --- |
| **PCV13** | **1** | 0.96 | 0.739 | 0.713 | 0.677 |
|  | **2** | 0.87 | 0.739 | 0.713 | 0.677 |
|  | **3** | 0.303* | 0.256 | 0.247 | 0.235 |
|  | **4** | 0.883 | 0.739 | 0.713 | 0.677 |
|  | **5** | 0.86 | 0.739 | 0.713 | 0.677 |
|  | **6-11** | 0 |  |  |  |
| **PCV15** | **1-5** | Identical to PCV13 VE |  |  |  |
|  | **6** | 0.86 | 0.739 | 0.713 | 0.677 |
|  | **7-11** | 0 |  |  |  |
| **PCV20** | **1-6** | Identical to PCV15 VE |  |  |  |
|  | **7** | 0 |  |  |  |
|  | **8, 9** | NA | 0.739 | 0.713 | 0.677 |
|  | **10, 11** | 0 |  |  |  |
| **PPSV23** | **1-4, 6-9** | NA | 0.585 | 0.559 | 0.521 |
|  | **5,10,11** | NA | 0 | 0 | 0 |

IPD: Invasive Pneumococcal Disease; STC: Serotype Class; NA: Not Applicable;
PCV13: 13-valent pneumococcal conjugate vaccine; PCV15: 15-valent pneumococcal conjugate vaccine; PCV20: 20-valent pneumococcal conjugate vaccine; PPSV23: 23-valent pneumococcal polysaccharide vaccine; STCs = serotype classes;
\*: left end point of 95%CI.

Populations in each adult age group include those at low risk for pneumococcal disease (healthy individuals) as well as those with increased risk due to immunocompromising conditions. The VEd of PCV13 for IPD in healthy adults was based on Bonten *et al*. [20] and in high-risk adults on Cho *et al.* [21], and VEd of PPSV23 was based on [22]. These individual efficacies were incorporated into the age-based population-level VEds based on risk distribution by age (Table S9).

Equivalency was assumed for the new vaccines, which means that new PCVs with additional STs (e.g PCV15 and PCV20) have equivalent VEds as previous PCVs (PCV13) for shared serotypes. Overall VEd of PCV13 [18] was used as VEd of PCV15 against STC6-type IPD in pediatric age groups. PCV13 VEd in adult age groups was not ST-specific and was extended to additional PCV15 and PCV20 STCs (Table 3).

In a scenario analysis STC-specific VEd for PCV15 was based on immune response of PCV15 relative to PCV13. Results of this scenario analysis are presented in the Supplementary Appendix.

It was assumed that mean duration of protection from any PCV was 10 years, while PPSV23 was assumed to have a mean duration of protection of 7.5 years [23].

### Carriage clearance rates

Annual carriage clearance rates for a given serotype were calculated as the inverse of carriage duration. A literature review was performed to collect data on age and serotype-specific carriage durations. For each age-STC combination the total carriage duration across studies was divided by the total population across studies and normalized by age group. More details on these derivations are available in Section 3.2.4.1.1 of the Supplementary Appendix.

### Model calibration

To infer the values of the age- and STC-specific probability of acquisition, age- and STC-specific case-carrier ratios, STC-specific competition parameters, and the STC-specific VEc for the VTs, the model was calibrated to historical US IPD incidence data from 2000–2019. The model’s fit to historical data of IPD incidence was conducted to minimize the sum of squared errors. Figure S11 of the Supplementary Appendix provides a visual comparison of the model’s fit to the data, with the fit error shown in each plot. The calibration results, i.e., the values of fitted parameters, such as invasiveness and carriage transmission probability are higher in extreme age groups, consistent with conventional wisdom and existing modeling literature.

Pneumococcal dynamics were severely affected by the COVID-19 pandemic [24] through reduced transmission due to non-pharmaceutical interventions and disruption of routine healthcare services including vaccination. Since the model does not account for such effects, data after 2019 were not included in model calibration. Calibration methodology and detailed results are described further in the Supplementary Appendix.

### Calibration target

Age-stratified STC-specific IPD incidence per 100,000 in the US from 1998-2019 was used as calibration target. The annual IPD incidence per 100,000 in the US by age group and STC was derived from two sources: (i) the available overall (all serotypes) incidence data and (ii) the serotype distribution data. Overall incidence was obtained from the annual CDC surveillance reports [25] and the annual serotype distribution data came from Active Bacterial Core surveillance, CDC [26].

For calibration to NBPP data, pediatric NBPP incidence per 100,000 from 1998–2018 was obtained from Hu *et al*. [27]. To obtain the counterpart data in adults, all-cause pneumonia (ACP) incidence was acquired from Truven MarketScan Commercial Claims and Encounters (January 1, 2012 to December 2020) – a healthcare claims database that contains information about clinical utilization over time [28]. It was assumed that 11% of ACP had pneumococcal etiology—NBPP—in US adults [29]. For pediatrics, it was assumed that ST distribution for NBPP followed that of IPD. For adults, the proportion of NBPP attributable to STCs was calculated using data from two recent studies [30, 31] and used to construct the STC- and age-specific calibration data (see Section 5.1 of Supplementary Appendix for additional details).

For AOM calibration, pediatric all-cause AOM incidence per 100,000 from 1998-2018 was obtained from Hu *et al*. [32] of which 23.8% was pneumococcal attributable [33]. Studies with ST distribution information for AOM cases [33, 34, 35] were used to construct the STC- and age-specific data (see Section 5.2 in Supplementary Appendix for additional details).

Using the carriage prevalence profile that corresponded to calibrated IPD incidence, case-carrier ratios for NBPP and AOM were then calibrated using the historical incidence of each manifestation. Details on NBPP and AOM calibration, procedure, and results are provided in the Supplementary Appendix.

### Model projections

The calibrated model was then used to compare pneumococcal disease burden assuming either pediatric PCV13 or PCV15 vaccination. In adults, both PCV15 with PPSV23 or PCV20 have been recommended for use [36]. A hypothetical combination of these vaccines was constructed. In addition, given the historic practice, we assumed that healthcare providers may continue to administer PPSV23 alone. Therefore, to design an adult vaccination scenario (which would be kept same while comparing the two pediatric vaccination scenarios) it was assumed that of the vaccinated US adults 80% receive PCV20, 10% receive PCV15+PPSV23, and 10% receive

PPSV23 alone. Since adult vaccination was identical between these two scenarios, the differences in model outcomes were solely caused by pediatric vaccination. A different adult vaccination scenario of 50% vaccinated adults receiving PCV20, 25% receiving PCV15+PPSV23 and 25% receiving PPSV23 (50-25-25 scenario) was examined in an additional scenario analysis.

The two scenarios are summarized below:

1. Scenario PCV13: Infants receive PCV13 according to assumed VCR. Given assumed VCR among adults 18+, 80% receive PCV20, 10% receive PPSV23, and 10% receive PCV15+PPSV23.
2. Scenario PCV15: Infants receive PCV15 according to assumed VCR. Given assumed VCR among adults 18+, 80% receive PCV20, 10% receive PPSV23, and 10% receive PCV15+PPSV23.

## Results

### IPD data calibration results

As shown in Fig 2 and 3, the model was able to capture historical trends in each of the STCs for each age group in the US (detailed model fit by STC/age group is given by Figure S11 in the Supplementary Appendix). Before the introduction of a PCV in the US, PCV7 serotypes (STC1) dominated the IPD landscape. However, after the introduction of PCV7 in 2000 and prior to the introduction of PCV13 in 2010, replacement occurred, with the additional PCV13 (excluding ST3) serotypes increasing in incidence. This replacement behavior was captured by the model by careful adjustment of the competition parameters. In 2010 PCV13 was introduced and all PCV13 STCs, other than ST3, declined. ST3, both in the model and data, remained relatively constant across the calibration period. Following PCV13 introduction, all non-vaccine STCs trended upward.

**Fig 2.**
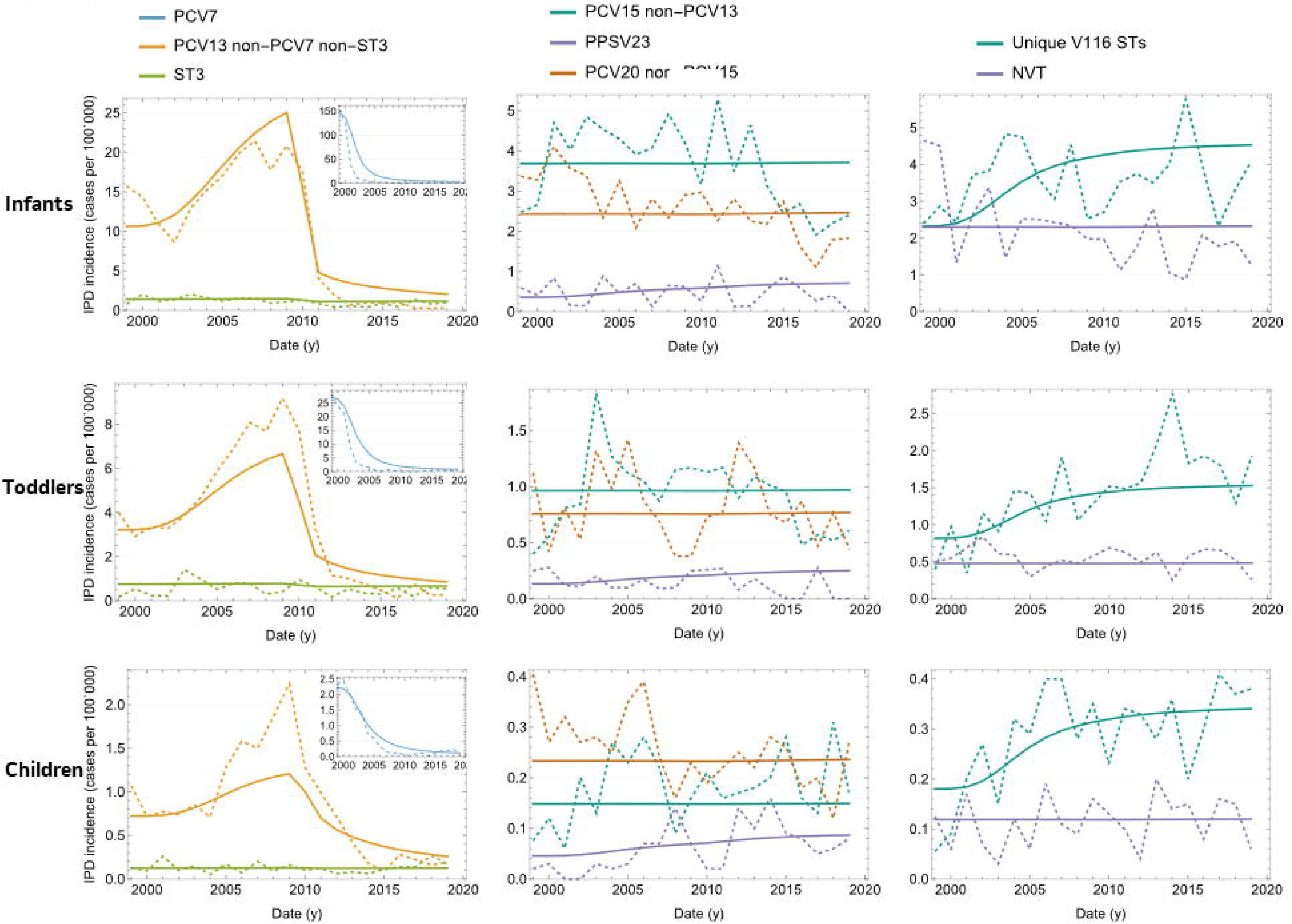
Calibrated versus observed IPD incidence per 100,000 over time in children age groups. Dashed lines: data; Solid lines: model outcome.

**Fig 3.**
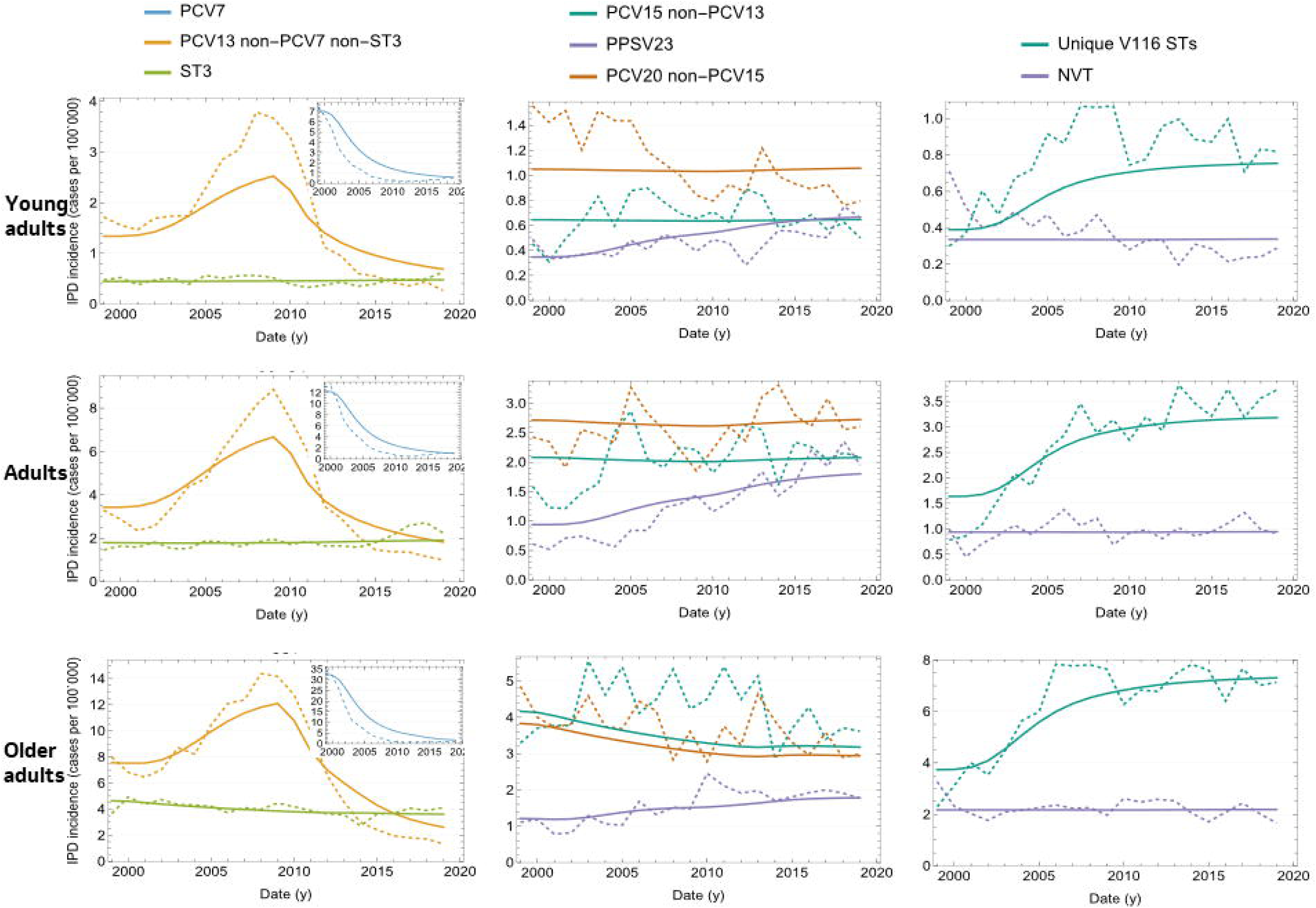
Calibrated versus observed IPD incidence per 100,000 over time in adult age groups. Dashed lines: data; Solid lines: model outcome. IPD = invasive pneumococcal disease; PCV7 = 7-valent pneumococcal conjugate vaccine; PCV13 = 13-valent pneumococcal conjugate vaccine; PCV15 = 15-valent pneumococcal conjugate vaccine; PPSV23 = 23-valent pneumococcal polysaccharide vaccine; ST3 = serotype 3; STCs = serotype classes; V116 = an investigational 21- valent pneumococcal conjugate vaccine.

Prior to PCV7, STC1 outcompeted other STs, but following PCV7 introduction, and the reduction in STC1 STs, STCs 4,7, and10 were able to expand due to the reduced competitive pressure from STC1 STs

Several years of real-world data following the introduction of new vaccines will be necessary to estimate how new vaccine type STs, such as 22F and 33F (STC6) compete and interact with other STCs.

The model estimated age- and serotype-specific VEc, which revealed PCV7 to have a greater impact against the seven serotypes it targeted than PCV13 had on the additional six serotypes it targeted (TableS18). The model also calculated age- and serotype-specific case-to-carrier ratios for IPD in the US (Table S17). Results indicated that these case-carrier ratios were highest in the infants and older adults.

Following the calibration of the model to IPD incidence, which generated age- and STC-specific carriage acquisition rates, the model was then calibrated to estimate the age- and STC-specific case-carrier ratios for each of AOM and NBPP. The model captured changes in the serotype distribution associated with AOM after different vaccine introductions, such as the decline of five of the additional serotypes included in PCV13 (excluding STC3) (Figure S14). Similarly, the model replicated reductions in PCV7 STs in 2000 and STC4 (7F, 19A) in 2010 in pediatric NBPP following the introduction of PCV7 and PCV13 (Figure S13).

### Epidemiological projections

The calibrated model was used to make projections in the US for the two scenarios described above (Fig 4). Projections revealed that PCV15 prevented more cases of IPD, AOM, and NBPP than PCV13 across all age groups. The reduced incidence of IPD in pediatric age groups under the PCV15 scenario reflected the direct and indirect benefit due to the added vaccine type serotypes 22F and 33F, while the lower incidence in adult age groups was due to indirect protection (i.e., herd immunity) from the pediatric population. It was estimated that over 10 years, PCV15 would reduce about 6% more cases of IPD in the US when compared to PCV13. This includes around 13.54 more cases averted in children under 2 and 4.82% more cases averted in adults over the age of 65. With the 50-25-25 scenario (50% vaccinated adults receiving PCV20 and 25% receiving PCV15+PPSV23 and 25% receiving PPSV23), PCV15 saved about 6.1% more cases of IPD than PCV13 over 10 years, including 4.9% more cases saved in adults over 65 years of age. As expected, the number of pediatric IPD cases averted remained unchanged.

**Fig 4.**
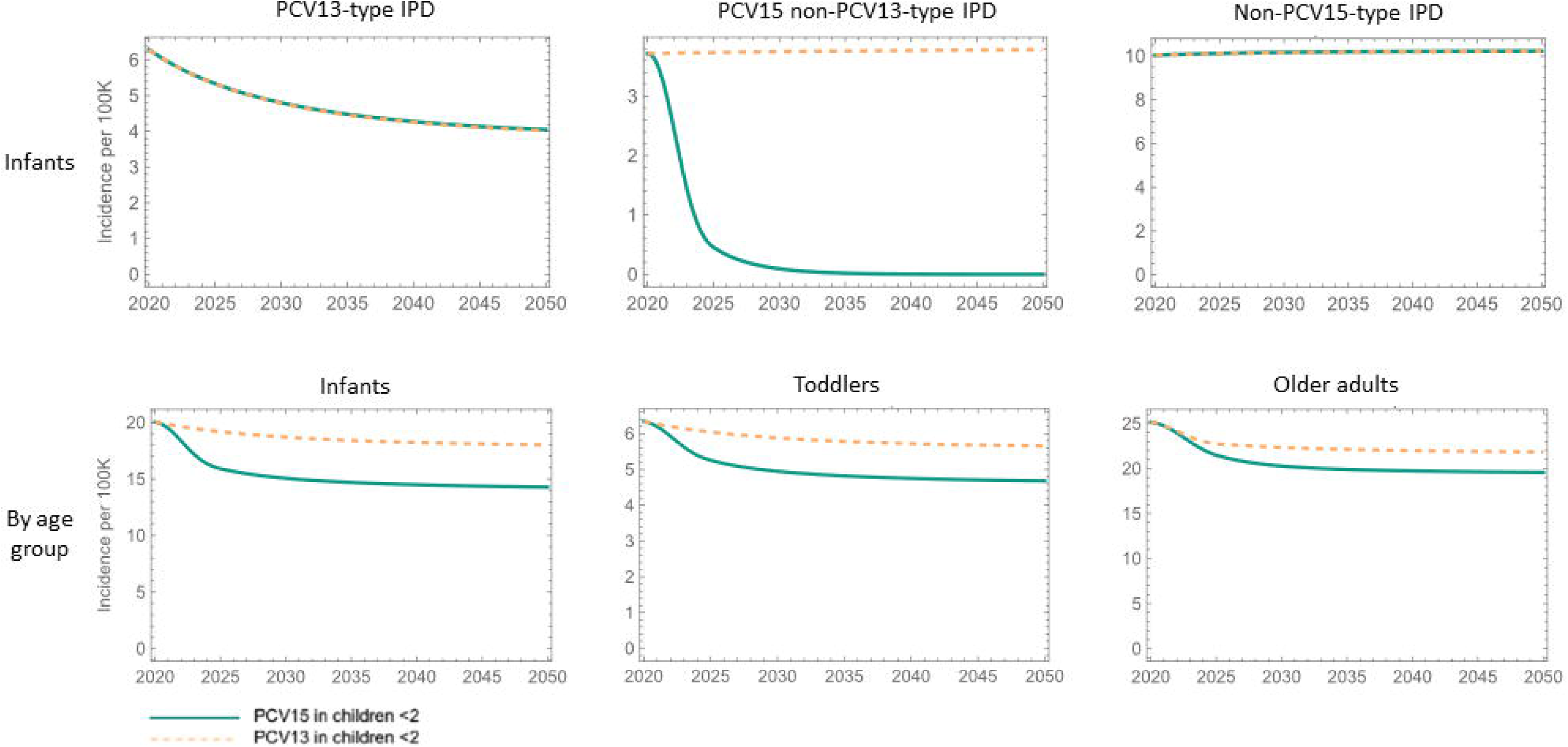
IPD incidence per 100,000 projected by the calibrated model. Top: IPD burden, infants and toddlers; Bottom: Overall IPD burden by age group. Vaccination in adults 18+: 80% receive PCV20, 10% receive PPSV23, and 10% receive PCV15+PPSV23. Vaccine coverage used in these projections is given by Figure S6. Reduced IPD incidence in adults is a result of indirect protection of pediatric PCV15 vaccination. IPD = invasive pneumococcal disease; PCV13 = 13-valent pneumococcal conjugate vaccine; PCV15 = 15-valent pneumococcal; STs = serotypes; VE = vaccine effectiveness

The model estimated that after 10 years, compared to PCV13, PCV15 will also reduce the number of NBPP cases by 5.5% and the number of AOM cases by about 3%.

### Projected IPD STC distribution

Figure 5 shows the pre-PCV, 2019, and 10 year-projections of IPD incidence in the US. By 2019, PCV7 and PCV13 reduced overall IPD, however ST-distributions shifted with NVTs replacing VTs over this same period. The third column shows the projected shift in this distribution following 10 years of PCV15 utilization. STs 22F and 33F (PCV15 non-PCV13) are diminished with PCV15. There is a clear shift from vaccine-types towards NVTs (Other STs) through ST replacement. Since PCV20 was implemented in adult age groups (18 and above) in projection period, IPD in the PCV20 non-PCV15 STC was reduced in adults.

**Fig 5.**
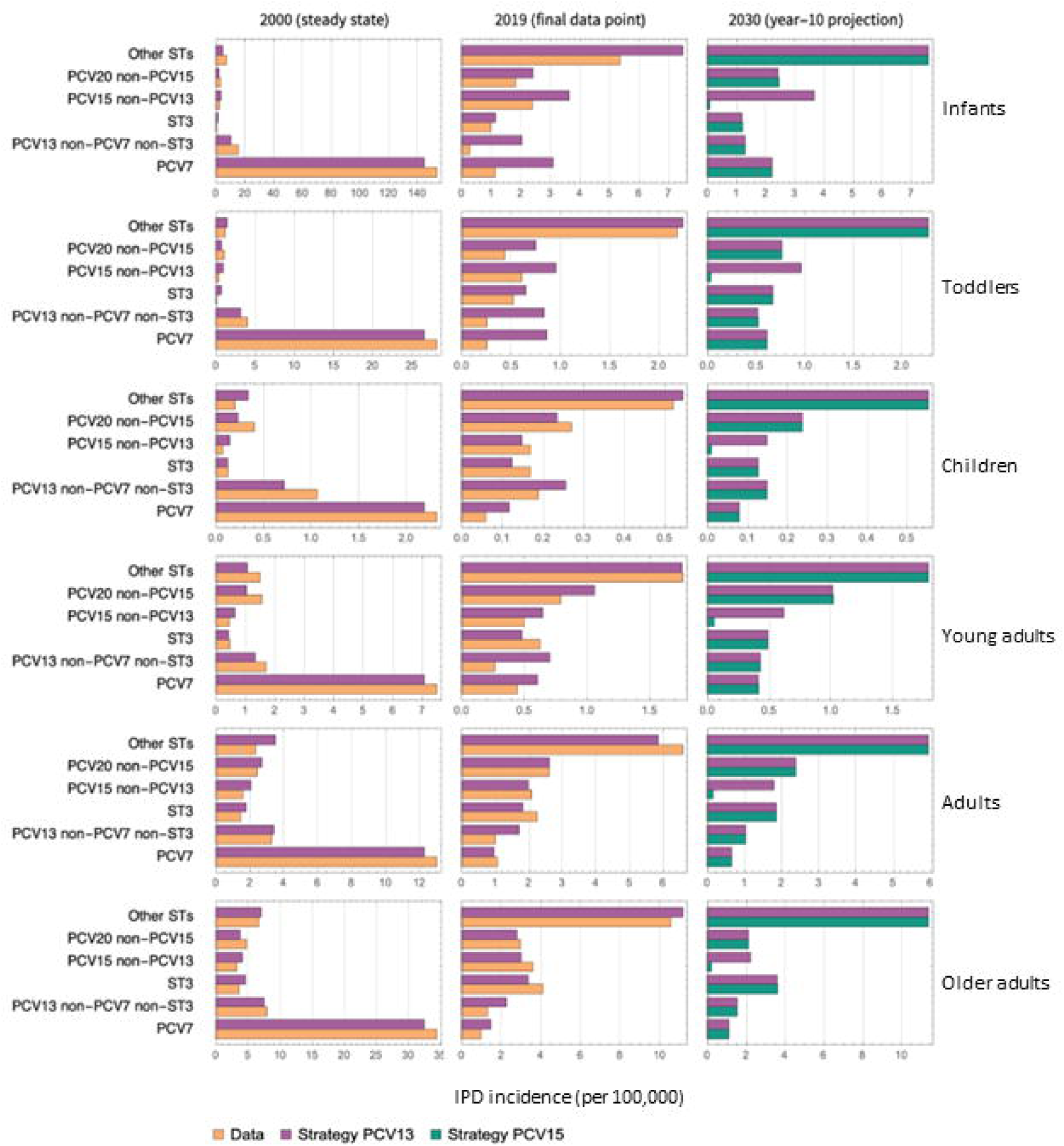
IPD incidence (cases per 100,000) attributed to each STC grouped by vaccine in each model age group. Model calibrated STC distributions and data at the start and end of calibration period (2000 and 2019 respectively), and model projected STC distributions after 10 years of PCV15 in infants. IPD = invasive pneumococcal disease; PCV7 = 7-valent pneumococcal conjugate vaccine; PCV13 = 13-valent pneumococcal conjugate vaccine; PCV15 = 15-valent pneumococcal conjugate vaccine; PPSV23 = 23-valent pneumococcal polysaccharide vaccine; ST3 = serotype 3; STCs = serotype classes

## Discussion

An age-structured dynamic model of pneumococcal carriage transmission was developed to evaluate the epidemiological impact of age-based vaccine policies on pneumococcal diseases. In this study, we calibrated the model to historical data from the United States, though the model could be adapted to other countries if data are available. Serotypes were aggregated into serotype classes (STCs) which allowed disease incidence to be examined by each age and STC combination.

The model was calibrated to the historical age-specific US IPD incidence data from 2000 to 2019 and used to project future outcomes under different vaccine strategies. The results of this calibration revealed age- and serotype-specific carriage acquisition probabilities, competition parameters between STCs, age- and serotype-specific vaccine efficacies against carriage, and age- and serotype specific case-to-carrier ratios for invasive and non-invasive pneumococcal disease (IPD, NBPP, and AOM). Calibrated values fell within the range of published estimates where available or were aligned with calibration estimates of published models.

Probability of carriage acquisition per contact with a colonized individual, which is an age- and STC-specific parameter, was highest in infants and older adults due to weak immune systems (i.e., immature in infants [37] and deficient due to age in older adults [38]). This implies that infants and toddlers (aged 0-4) and older adults are most likely to acquire carriage upon contact with a colonized individual of any age. These results are consistent with other literature (for example, see [11, 16, 39]). Case-to-carrier ratios for IPD followed a similar pattern, with infants and older adults most likely to develop disease from carriage acquisition [40]. Calibration estimates of case-to-carrier ratios for IPD also fall within the range of published estimates [40, 41, 42, 43].

The calibrated competition parameter estimates correctly reflected the ST replacement in the US whereby NVTs increased to fill the depleted niche due to target STs declining following PCV7 and PCV13 implementation. Modeling literature highlights the role of ST competition [44] and some pneumococcal transmission models have included this feature at a higher level or in some aggregation [12, 45, 46]. However, calibrated ST competition between PCV7 STs against non- PCV7 STs, and PCV13 STs against non-PCV13 STs, to our knowledge, has not been examined at such fine granularity before.

Our parameter estimates of STC-specific VEc aligned with those reported in published literature [47, 48]. Like pneumococcal disease, PCVs are also less protective against ST3 acquisition [49]. This is evident from the relatively low calibrated VEc against ST3 which was also the case with other published model calibrations that reported ST-specific VEc [11]. Additionally, VEc in adult age groups is lower than the respective STC-specific VEc in children age groups. Since carriage acquisition is a prerequisite for pneumococcal disease, protection against acquisition is the first level of defense that PCVs offer; reduction in acquisition will lead to reduction in disease.

Since dynamic transmission models estimate the time-dependent dynamics of population-level susceptibility along with factors that affect the transmission rate (e.g., vaccination status and carriage acquisition in response to mixing patterns), they are well suited to provide insights into the indirect protection provided from interventions, such as including vaccination. For example, a recent study [10] used dynamic transmission models to compare the epidemiologic impact of switching from PCV13 to PCV15 or PCV20 in infant schedule in England. One of the research findings of this study reveals that transitioning from PCV13 to PCV15 in the pediatric vaccination program in England resulted in the unintended consequence of increased incidence of IPD. However, the implementation of our model to the US data using a different structure and assumptions demonstrated a decline in IPD cases if a transition from PCV13 to PCV15 were to occur. Indirect protection of pediatric vaccination on adults which has been observed in programs where pneumococcal vaccines have been implemented in pediatric programs [50], can be analyzed in detail using this model. Here, identical vaccination strategies in adults were kept and the two different vaccination programs in infants, PCV13 versus PCV15, were compared.

Due to indirect protection (i.e., herd immunity), the implementation of PCV15 in pediatrics led to additional reductions in adult disease attributed to 22F and 33F.

The model does have some limitations. First, because of lack of data to estimate some parameters such as STC-specific probability of SP acquisition we inferred the values of some of these parameters by calibrating the model outcomes to historical data. Second, certain serotypes, such as serotype 3, have evolved under pressure from vaccination [51, 52] but the model does not account for such evolution, assuming the same properties of STCs throughout the calibration and projection horizons. Third, since there is very little evidence of meaningful transmission from adults to children, our model assumes no transmission from adults to children [14] so there is no indirect effect of vaccinating adults on children. Though this conservative assumption helps prevent any overstatement of the potential value for adult vaccines, it may limit any implications on model projections arising from possible adults-to-children transmission. Fourth, including several STs into a single STC entails strong assumption that STs in the same STC have identical properties. Fifth, it is assumed that infants either receive full vaccination series or no vaccination at all. This is a mathematically simplifying assumption to limit parameterization and model complexity. Sixth, due to lack of data for the efficacy of PCV15 and PCV20, these values were assumed based on the efficacy of PCV13. Seventh, the model assumes that no immunity is gained after infection.

The flexibility and accuracy of the model demonstrates promise for future adaptions in other country settings, where local pneumococcal epidemiology may vary, such as different age- reporting, vaccine history, vaccine dosing, vaccine efficacies, different STs of interest, or different policy questions. The model implemented here focused on epidemiological outcomes, but in future work the model could additionally be used to evaluate the cost-effectiveness of different vaccines or vaccination strategies. Understanding that certain viral pathogens, such as respiratory syncytial virus, influenza, and human metapneumovirus may facilitate IPD [53, 54, 55, 56], historical data from these pathogens can also be included into the model to capture some of the unexplained inter-annual variation in IPD incidence.

## Data Availability

ST distribution in annual invasive pneumococcal disease (IPD) incidence in the US which was used to generate IPD incidence model calibration target (orange curve, Figure S11) is publicly available at: https://data.cdc.gov/Public-Health-Surveillance/1998-2022-Serotype-Data-for-Invasive-Pneumococcal-/qvzb-qs6p/about_data All other data produced in the present work are contained in the manuscript and supplementary appendix.

## Acknowledgements

The authors acknowledge the support of Eric Sarpong in the acquisition of NBPP incidence data in adults, John Lang for his constructive feedback on the manuscript and supplementary appendix, and the Scientific Advisory Committee composed of multiple experts for their advice throughout model development and application.

## Supporting information

S1 Text. Supplementary Appendix

