## Supplementary Appendix for "A dynamic transmission model for assessing the impact of pneumococcal vaccination in the United States"

### Supplementary Appendix
### A technical Report Accompanying Manuscript:

#### A dynamic transmission model for assessing the impact of pneumococcal vaccination in the United States.

Tufail M Malik, Kevin M Bakker, Rachel J Oidtman, Oluwaseun Sharomi, Giulio Meleleo, Robert B Nachbar, Elamin Elbasha.

January 30, 2025

### CONTENTS

1.1 List of Figures ... 4
1.2 List of Tables ... 5
1.3 List of Abbreviations ... 6
2 Introduction ... 8
2.1 Pneumococcal dynamic transmission models - a review of published models ... 8
2.2 The model ... 16
3 Model description ... 16
3.1 The demographic model ... 16
3.1.1 Demographic model structure ... 16
3.1.2 Estimates of the demographic model parameters ... 17
3.1.3 Population mixing patterns ... 19
3.2 The epidemiologic model ... 23
3.2.1 Major modeling assumptions ... 24
3.2.2 Epidemiologic model structure ... 25
3.2.3 Model flowchart ... 29
3.2.4 Model equations ... 33
3.2.5 Estimates of the epidemiologic model parameters ... 41
4 Model calibration to vaccination coverage data and projected coverage ... 51
4.1 History of ACIP recommendations of pneumococcal vaccines ... 51
4.2 Implications of ACIP recommendations for the model ... 51
4.3 Vaccine-only model ... 52
5 Model calibration to IPD data ... 53
5.1 IPD incidence ... 54
5.2 Calibration procedure ... 54
5.3 Calibration results ... 61
5.3.1 Calibrated parameters ... 61
5.3.2 Results of model fitting ... 63
6 Model calibration to NBPP and AOM data ... 69

#### 1.1 List of Figures

Figure S1. Age distribution for 2017 US population and model projection with different annual growth rate assumptions. (a) *q* = 0%, (b) *q*=0.5%, (c) *q*=1.7%. 18

Figure S2. Step-by-step transformation of the mixing matrix, from the originally published age groups in Prem et al. (30) to the model age groups. 21

Figure S3. Model flowchart describing carriage transmission and vaccination dynamics in unvaccinated compartments. 30

Figure S4. Model flowchart describing carriage transmission and vaccination/waning dynamics among vaccinated compartments. 31

Figure S5. Model flowchart describing carriage transmission and vaccination dynamics among compartments with waned vaccine protection. 32

Figure S6. Fitted and projected vaccination coverage in respective age groups. 53

Figure S7. US pneumococcal vaccine recommendation timings and IPD incidence. $T_0$, $T_1$ and $T_2$ represent calibration periods. Red denotes new or changed recommendation. RG: Risk Group. 54

Figure S8. Carriage prevalence at steady state. Left: Carriage data from Cleary *et al*. [82] was used as a reference point for carriage values by vaccine type (blue). This was compared to the carriage levels in the model at pre-PCV steady state (orange). Right: Carriage levels assumed in model age groups at pre-PCV steady state. 58

Figure S9. Initial fitting of the steady-state IPD incidence (per 100,000) by age group for each STC. Orange denotes the data and green denotes the model. 59

Figure S10. Model calibration versus IPD incidence (per 100,000) in 2000 by age group for each STC. Orange denotes the data and green denotes the model outcome. 64

Figure S11. Model outputs versus IPD incidence (per 100,000) data by year for each STC and each age group. Vertical green lines denote the time of implementation of the PCVs (PCV7 in 2000 and PCV13 in 2010). Mean square difference for each plot was minimized. The 'fit error' shows this mean square difference at the fit. 66

Figure S12. Model calibration versus IPD incidence (per 100,000) data by year for each age group. Dashed denotes the data and solid line denotes the model outcome. 67

Figure S13. Model fitting of the overall IPD incidence (per 100,000) aggregated over age group and STC. Orange denotes the data and green denotes the model. 68

Figure S 14. Model validation: Comparison of carriage prevalence in the US children under 5 as generated by the model with data from Sharma et al. and Desai et al. for children under 5, and Lee et al. for children under 7. 69

Figure S15. Model calibration to the NBPP incidence data by year for each age group. Pediatric data from 1998 was used for calibration, but adult data was only available beginning in 2012, which was used for calibration. 73

Figure S16. Model calibration to the AOM incidence data by year for each age group. 75

#### 1.3 List of Abbreviations

| | |
|---|---|
| ABCs | Active Bacterial Core Surveillance |
| ACIP | Advisory Committee on Immunization Practices |
| AG | Age group |
| AOM | Acute otitis media |
| AR | Antibiotic resistant |
| AS | Antibiotic susceptible |
| CDC | Centers for Disease Control and Prevention |
| CFR | Case fatality rate |
| CI | Confidence interval |
| CMC | Chronic medical conditions |
| DTM | Dynamic transmission model |
| IBM | Individual based model |
| IC | Immunocompromised |
| IPD | Invasive pneumococcal disease |
| MMWR | Morbidity and Mortality Weekly Report |
| NBPP | Non-bacteremic pneumococcal pneumonia |
| NIP | National immunization program |
| NIS | National Immunization Survey |

| | |
|---|---|
| NVT | Non-vaccine-type |
| ODE | Ordinary differential equation |
| PCV | Pneumococcal conjugate vaccine |
| PCV10 | 10-valent pneumococcal conjugate vaccine |
| PCV13 | 13-valent pneumococcal conjugate vaccine |
| PCV20 | 20-valent pneumococcal conjugate vaccine |
| PPSV23 | 23-valent pneumococcal polysaccharide vaccine |
| PD | Pneumococcal disease |
| SCDM | Shared clinical decision-making |
| SEI | Susceptible-Exposed-Infected |
| SI | Susceptible-Infected |
| SIRS | Susceptible-Infected-Recovered-Susceptible |
| SIS | Susceptible-Infected-Susceptible |
| SIR | Susceptible-Infected-Recovered |
| SP | *Streptococcus pneumoniae* |
| ST | Serotype |
| STC | Serotype class |
| SVIRD | Susceptible-Vaccinated-Infected-Recovered-Dead |
| UK | United Kingdom |
| US | United States |
| VE | Vaccine efficacy |
| VEd | Vaccine efficacy against disease |
| VEc | Vaccine efficacy against carriage acquisition |
| VT | Vaccine-type |
| WHO | World Health Organization |

### 2 INTRODUCTION

This supplementary technical appendix comprises background information related to the development of "A dynamic transmission model for assessing the impact of pneumococcal vaccination". It includes detailed descriptions of previous literature, methodological considerations, including model's assumptions, equations, outcomes, and inputs.

#### 2.1 Pneumococcal dynamic transmission models - a review of published models

A targeted literature review was carried out to identify the published dynamic transmission models of pneumococcal vaccination. This non-exhaustive review covered pneumococcal dynamic transmission models published since 2000 (apart from the articles by Marc Lipsitch in 1997 and 1999 [1, 2] which provided a foundational structure of pneumococcal transmission dynamics).

Numerous dynamic transmission models of pneumococcal carriage and disease have been published that divide the population into compartments of individuals who are susceptible (S), vaccinated (V), exposed (E), infected (I), recovered (R), or dead due to SP-induced death (D). They range from deterministic to stochastic to agent-based models. The review article by Lochen *et al*. [3] observes that deterministic, dynamic models of varying structure have been used to model pneumococcal carriage and disease, including SIS, SIR, SEI, SI, SIRS and SVIRD though the majority adopts an SIS structure [1, 2, 4, 5, 6, 7, 8, 9, 10, 11]. This diversity affirms the complexity of natural immunity against SP and scarcity of data on recovery from pneumococcal carriage and pneumococcal disease. Furthermore, Lochen *et al*. [3] notes that typically the economic evaluations are not based on transmission dynamic models, thereby either ignoring or arbitrarily imputing the herd immunity effects and argues in favor of using both the dynamic transmission models and economic evaluations in conjunction for decision making on designing vaccination programs. Table S1 provides an overview of different modeling approaches and their major findings.

Table S1. Dynamic models of pneumococcal transmission.

| Study | Model structure | Setting | Objective(s) | Salient features of the model | Major finding(s) | Age structure? | Vaccination? | Co-colonization? | No. of STs |
|---|---|---|---|---|---|---|---|---|---|
| Bottomley *et al*. 2013 [12] | SIR | The Gambia | To predict the prevalence of VT and NVT serotypes following PCV7 introduction in The Gambia. | A proportion of infected individuals acquires lifelong serotype-specific immunity after infection | Vaccination eliminates low and medium prevalence serotypes, but overall carriage prevalence is reduced slightly due to serotype replacement | N | Y | N[a] | 6 groups:<br><br>VT/NVT, low/medium /high prevalence types |
| De Cao *et al*. 2014 [13] | SIS | Netherlands | To assess the impact of PCV13 on IPD | Model of Melegaro *et al*. [4] was adapted to Dutch data | PCV13 reduces IPD to levels greater than previously determined if herd protection considered | Y | Y | Y[b] | 2 groups:<br><br>VT/NVT |
| Choi *et al*. 2011 [5] | SIS | England and Wales | To predict the impact of vaccine on VT- and NVT-related IPD incidence | Co-colonization included but simultaneous acquisition ignored. | Despite serotype replacement, PCV7 vaccination could result in a decrease in IPD | N | Y | Y[b] | 2 groups:<br><br>VT/NVT |

| | | | | | | | | | |
|---|---|---|---|---|---|---|---|---|---|
| Choi *et al*. 2012 [6] | SIS + IBM | England and Wales | To assess the effect of switching from PCV7 to PCV13 and that of no vaccination on IPD | Age structure, competition effect between two serotype groups | Reduction in IPD after PCV7 to PCV13 switch. | Y | Y | Y[b] | 3 groups: PCV7, PCV13-PCV7, NVT |
| Choi 2019 *et al*. [14] | SIS | England and Wales | To assess the impact of 2+1 to 1+1 dosing schedule | Model in Choi *et al*. [6] fitted to septicaemia and meningitis incidence | Dropping one vaccine dose results in extremely small pneumococcal disease | Y | Y | Y[b] | 3 groups: PCV7, PCV13-PCV7, NVT |
| Cobey *et al*. 2012 [7] | SIS | Developed country | To investigate how weak serotype-specific immune response can support coexistence | Colonization, co-colonization, antibiotic and vaccination | Interaction of acquired serotype-specific & nonspecific immunity produces coexistence | Y | Y | Y[c] | 25 (default) Analysed 15, 35 |
| Colijn *et al*. 2010 [15] | SIS | - | To investigate the mechanisms of coexistence of antibiotic susceptible (AS) and antibiotic resistant (AR) serotypes | Structurally neutral models with heterogeneity promoting coexistence – as opposed to implicit mechanisms leading to coexistence | Coexistence is rare, except with (i) simultaneous dual transmission of AS and AR strains, and (ii) stronger self-immunity than cross-immunity | N | N | Y | 2 Drug-susceptible, drug-resistant |

| | | | | | | | | | |
|---|---|---|---|---|---|---|---|---|---|
| Flasche *et al*. 2013 [16] | SIRS | High income | To demonstrate that competition/coexistence could be determined by serotype-specific and nonspecific immunity | Individual-based model; a bivalent & a universal vaccine; | Interaction of acquired serotype-specific & nonspecific immunity produces coexistence (similar to Cobey *et al*. [7]) | Y | Y | Y | 20 |
| Flasche *et al*. 2017 [17] | SIS | Kilifi, Kenya | To estimate the dose efficiency of catch-up PCV campaigns. | Includes IPD; VT/NVT; Catch-up vaccination | Catch-up campaigns can increase efficiency of PCVs | Y | Y | Y | 2<br>VT/NVT |
| Gaivao *et al*. 2017 [18] | SIS | - | To examine competition and facilitation among two co-colonizing serotypes. | No vaccine or other intervention. | Heterogeneity in clearance and transmission favors endemic persistence. Backward bifurcation at $R_0$=1; | N | N | Y | 2 |
| Gjini 2016 [19] | SI, SIS | NA | To illustrate how prevalence data post-licensure can be used to estimate vaccine efficacy | Two serotype groups considered | How prevalence ratios in vaccinated/non-vaccinated hosts depend on VE, VT/NVT competition and transmission intensity | N | Y | Y | |
| Gjini *et al*. 2016 [20] | SIS | Portugal | To understand coexistence | Competition between VT-VT, VT-NVT, NVT-NVT; | Competition for carriage mediates stable coexistence only when competitive abilities satisfy certain pairwise asymmetries | N | Y | Y | 2 VT/NVT |

| | | | | | | | | | |
|---|---|---|---|---|---|---|---|---|---|
| Gjini 2017 [21] | SIS | Multi-country | A report on the application of model in Gjini *et al*. [19] | | Multiple factors impact performance of a vaccine. Applying mechanistic approaches to data retrospectively can help address not only pathogen-intrinsic variability but also of host populations. | N | N | Y | 2 |
| Iannelli *et al*. [8] | SIS | NA | To study the dynamics of a two-strain disease with superinfection (strain A infects those already infected by strain B), no coinfection. | Superinfection; no co-colonization; both strains are VT | If regulated by superinfection, serotype replacement possible even with perfect vaccine. | N | N | N | 2 |
| Lipsitch 1997 [1] | SIS | NA | To analyse competition in a two-serotype carriage with co-colonization. To predict factors and extent of serotype replacement. | Co-colonization; Strain A is VT, B is partially or fully VT, or NVT. | The presence of an NVT synergizes with the vaccine to reduce the VT transmission. | N | Y | Y | 2 |
| Lipsitch 1999 [2] | SIS | NA | To highlight the benefit of sero-replacement. To suggest factors to consider in choosing serotype composition of vaccines | Effects of competitive interactions can be analysed easily. | Sero-replacement depends on many factors including VT prevalence pre-vaccine, and vaccine coverage. | N | Y | Y | 2<br>Non-specific groups |

| | | | | | | | | | |
|---|---|---|---|---|---|---|---|---|---|
| Malik *et al*. 2021 [22] | SI | NA (with application to US data) | To assess cohort vaccination of pediatric population | Tracking natural history of carriage. Partial natural immunity following carriage. Co-carriage/competition. | Forecast of carriage prevalence. Basic and effective reproduction numbers derived. Conditions for coexistence and serotype-replacement theoretically derived. | N | Y | Y | 2<br><br>VT/NVT |
| Melegaro *et al*. 2010 [4] | SIS | UK/US | To predict the PCV7 impact on IPD incidence | Same model as in Choi 2012, fitted to US data. | It is important to include VT-NVT competition in a model | Y | Y | Y | 2<br><br>VT/NVT |
| Snedecor *et al*. 2009 [9] | SIS | US | To assess the direct and indirect effects of infants PCV7 in other age groups | Age structure (6 groups); doses considered. | Model fits IPD incidence data in <5 and >=65. There is no PCV7 threshold coverage level (after which there will be no IPD incidence decline). | Y | Y | N | 1<br><br>No specific STs were considered |
| Sutton *et al*. 2008 [23] | SEI | Australia | To assess the impact of pneumococcal vaccines provided to large risk groups | Seasonal infection rate; age stratification. | Parameters such as vaccine efficacy and infection rate depend on relative risk of unvaccinated to vaccinated individuals. | N | Y | N | 1<br><br>No specific STs were considered |

| | | | | | | | | | |
|---|---|---|---|---|---|---|---|---|---|
| Sutton *et al*. 2010 [23] | SEIS | Developed country | To study the population-level impact of vaccination strategies on pneumococcal disease | Age-structure; seasonal infection rate | Targeting carriage (vs infection) has biggest impact | Y | Y | N | 1<br><br>No specific STs were considered |
| Temime *et al*. 2004 [10] | SIS | France | To assess the impact of PCV7 and a hypothetical PCV11 on carriage and antibiotic resistance | Treatment and vaccination; antibiotic resistance. | PCV's are expected to control the selection of antibiotic resistance since by preventing carriage, they lessen antibiotic pressure. | Y | Y | N | 2<br><br>VT/NVT |
| van Effelterre *et al*. 2010 [11] | SIS | High income country | To assess the role of vaccination and treatment on IPD incidence | PCV7; antibiotic treatment; co-colonization. | Increase in 19A is more attributable to antibiotic pressure than PCV7. | Y | Y | Y | 18<br><br>(18 STs or groups of STs modelled individually) |

| | | | | | | | | | |
|---|---|---|---|---|---|---|---|---|---|
| Wasserman *et al*. 2018 [24] | SIS | UK | To assess how much IPD burden will increase after changing 2+1 dosing to 1+1 schedule | Vaccination; doses; | IPD incidence will increase substantially with the change in dosing schedule | N | Y | N | 5<br><br>19A, 3, remaining PCV13 serotypes not covered by PCV7 (1, 5, 7F, 6A), PCV7, NVT |
| Wu *et al*. 2012 [25] | SVIRD | Taiwan | To carry out long-term cost-effectiveness of PCV13 in Taiwan | Age structure; PCV13; a compartment of SP induced death | PCV13 is cost-effective in Taiwan with the WHO-recommended cost-effectiveness threshold from both the payer and societal perspectives | Y | Y | N | 1<br><br>No specific STs were considered |

a: Co-colonization was considered in supplementary appendix for sensitivity analysis. b: Co-colonization with 2 ST groups. c: Co-colonization with non-specific number of STs.

#### 2.2 The model

The framework constructed in this manuscript is a dynamic transmission model which describes pneumococcal carriage transmission dynamics and the potential disease progression in the presence of age- and serotype class-specific pneumococcal vaccines. The model, which is based on a system of ordinary differential equations, is constructed by dividing the population into age-specific compartments based on their pneumococcal carriage status, and vaccine protection status. It extends many of the published models by incorporating additional functionalities and features.

### 3 MODEL DESCRIPTION

The model describes the transmission of SP between humans and has two major components. The first part includes the demographic aspects of the population. This component is intended to mimic the age structure of a population such as births and deaths in each age group, contact patterns among different age groups, and growth from one age group to the next. The second component of the model consists of the epidemiologic model and includes carriage transmission.

#### 3.1 The demographic model

##### 3.1.1 Demographic model structure

The demographic model is based on the initial-boundary-value problem for age-dependent population growth described in [26]. The total population at time t, $\mathcal{P}(t)$, is divided into nA age groups in the age intervals ($A_0$, $A_1$), [$A_1$, $A_2$), …, [$A_{nA-1}$, $A_{nA}$). The total number of individuals $\mathcal{P}_a(t)$ at time t in the age group a given by the interval [$A_{a-1}$, $A_a$) is the integral of the age distribution function over this interval.

The differential equation representing the rates of population changes in the $a$th age group $[A_{a-1}, A_a]$ can be written as follows [26] (where prime represents differentiation with respect to time):

$$\mathcal{P}'_a = \delta_{1a}\Lambda + (1-\delta_{1a})m_{a-1}\mathcal{P}_{a-1} - [(1-\delta_{nA\,a})m_a + \mu_a]\mathcal{P}_a, \quad 1, 2, \dots, a, \dots, nA, \tag{1}$$

where, $\delta_{ij}$ is the Kronecker Delta (i.e., unity if $i = j$, and $0$ otherwise), $\mu_a$ is mortality hazard of age group $a$, and $m_a$ represents the maturation rate from age group $a$ to age group $a$+1, given by [26]

$$m_a = \frac{\mu_a + q}{exp[(\mu_a + q)(A_a - A_{a-1})] - 1}. \tag{2}$$

Here, *q* is the annual population growth or decay rate. Solving equation (1) at steady-state and using the fact that at any time $t, \mathcal{P}(t) = \sum_a \mathcal{P}_a(t)$, yields

$$\Lambda = \bar{\mathcal{P}}(m_1 + \mu_1)\left[1 + \sum_{a=2}^{nA}\left(\prod_{j=2}^{a}\frac{m_{j-1}}{m_j + \mu_j}\right)\right]^{-1}, \tag{3}$$

where $\bar{\mathcal{P}}$ is total population at the steady state.

The demographic parameters are described in Table S2.

Table S2. Description of the demographic model variables and parameters.

| Symbol | Description |
|---|---|
| $\mathcal{P}_a$ | Population size (number of individuals) in age group *a* |
| $\Lambda$ | Birth rate (recruitment into the model) |
| $\mu_a$ | Mortality hazard in age group *a*. |
| $m_a$ | Aging rate from age group *a* to *a+1*. |
| $f_a$ | Fertility rate in age group *a*. |
| *q* | Annual population growth rate. |

##### 3.1.2 Estimates of the demographic model parameters

For US calibration, the model divides the population into six age groups, < 2, 2-4, 5-17, 18-49, 50-64, and 65+. In all relevant analyses we use the interval 65-100 to represent the last age group. Mortality hazard in an age group is the probability of death from natural causes in the given age group. Age-specific life tables for the baseline year 2017 were obtained from the US Census Bureau website [27], from which mortality hazards ($\mu_a$) for each model age group were estimated, and are given in Table S3.

The annual growth rate *q* of the demographic model satisfies a modified age-group form of the Lotka characteristic equations [26]:

$$1 = \left[f_1 + f_2\frac{m_1}{q + m_2 + \mu_2} + \cdots + f_n\frac{m_{n-1} \dots m_1}{(q + m_n + \mu_n) \dots (q + m_2 + \mu_2)}\right] / (q + m_1 + \mu_1), \tag{4}$$

Using the birth rates per woman in US in 2017 (US Census Bureau data), we estimate the fertility rates for age groups 5-17 and 18-49 to be 0.00253386 and 0.0565967, respectively. Fertility rates for other age groups are assumed to be 0. Solving equations (2) and (4) simultaneously with these fertility rates yields the growth rate $q$ = 1.7% for 2017.

Population distributions in the six model age groups in 2017 with $q$ = 0%, 0.5% and 1.7% growth rates, together with data from the 2017 population census are plotted in Figure S1. With all growth rate assumptions, the model fits well for ages 0 to 18. With 0% and 0.5% growth rates it underestimates the population in age groups 18-49 and 50-64 and overestimates in age group 65+. With the 1.7% growth rate assumption, the model overestimates for age group 18-49, underestimates for 50-64 and again slightly overestimates for 65+. The model does not take into consideration special characteristics of the population such as immigration.

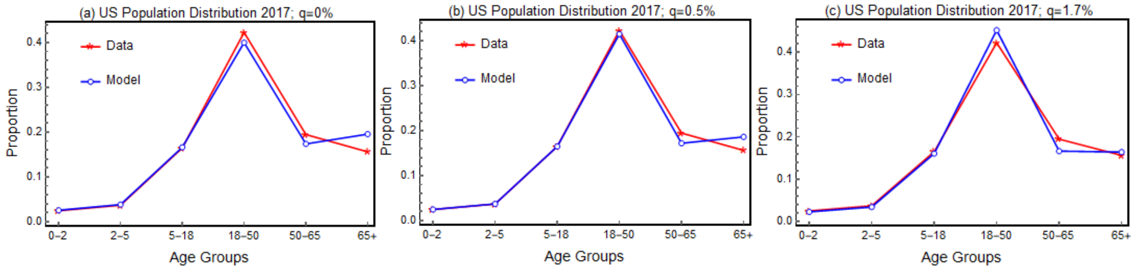

Figure S1. Age distribution for 2017 US population and model projection with different annual growth rate assumptions. (a) $q$ = 0%, (b) $q$=0.5%, (c) $q$=1.7%.

Because of this, and to be certain all results are due to vaccination and not strange demographics, we kept the population constant across the calibration and projection periods (i.e., we assume $q$ = 0). It is worth highlighting that the recent estimate of US Census Bureau indicates the slowing growth of the US resident population, with annual growth in 2017 being only about 0.79% [27]. With $q = 0$ and mortality hazards $\mu_a$ provided in Table S3, Equation (2) yields age-group specific maturation rate $m_a$, given in Table S3.

Table S3. Mortality hazards and aging rates for model groups [27] .

| Age group | < 2 | 2 - 4 | 5 - 17 | 18 - 49 | 50 - 64 | 65+ |
|---|---|---|---|---|---|---|
| $\mu_a$ | 0.00294255 | 0.000252028 | 0.000185549 | 0.00141466 | 0.00706467 | 0.0561529 |
| $m_a$ | 0.49853 | 0.333207 | 0.0768303 | 0.030548 | 0.0631967 | 0 |

Using a total population $\mathcal{P}(t) = 325{,}511{,}184$ in the year 2017 [27] with the mortality hazards and aging rates provided in Table S3, the annual birth rate $\Lambda$ is estimated to be 4,198,555 using equation (3).

##### 3.1.3 Population mixing patterns

Since the model incorporates age structure, it is important to account for the heterogeneity due to mixing patterns between different age groups. Inter-age mixing patterns impact the transmission dynamics of infectious diseases, particularly when the epidemiology is a characteristic of age as is the case with pneumococcal carriage acquisition and transmission as well as susceptibility to, and burden, severity and duration of the underlying disease. The classical vaccine-induced herd protection threshold assuming a homogeneous population can be substantially higher than the threshold in the presence of such heterogeneities in the populations [28].

Mixing patterns among a population are typically based on diary-based survey data. They provide a good approximation in the absence of mixing data specific to the transmission of certain disease. The major complication in adapting these mixing patterns arises from the mismatch between age groups of the survey population versus the age groups of the model. For the current model, estimates of the mixing matrix are derived from those published by Prem *et al*. [29], which are in turn projected from the POLYMOD study in Mossong *et al*. [30] wherein the daily estimates correspond to mixing patterns among 5-year age groups. The derived mixing matrix for the model is shown in Table S4, where each entry of the matrix is an estimate of the number of contacts an individual in an age group represented by a row makes in one day, on the average, with an individual from an age group listed in a column. The detailed derivation of Table S4 is given in Section 3.1.3.1, using a general framework that transforms an available mixing matrix among certain age groups to a mixing matrix in the desired age groups.

Table S4. Mixing matrix for US model age groups. Each number in the table represents the average number of contacts an individual in an age group in the row makes per day with an individual from an age group in the column.

| **Age Group** | **0-1** | **2-4** | **5-17** | **18-49** | **50-64** | **65+** |
|---|---|---|---|---|---|---|
| **0-1** | 1.041976 | 1.556262 | 1.789593 | 4.120562 | 0.776557 | 0.248696 |
| **2-4** | 1.041976 | 1.556262 | 1.789593 | 4.120562 | 0.776557 | 0.248696 |
| **5-17** | 0.21374 | 0.319236 | 9.121789 | 5.553655 | 0.662356 | 0.191759 |
| **18-49** | 0.188274 | 0.2812 | 2.537017 | 9.979767 | 1.518593 | 0.180047 |
| **50-64** | 0.172911 | 0.258254 | 1.933061 | 6.391688 | 2.961316 | 0.425319 |
| **65+** | 0.082389 | 0.123054 | 0.900301 | 2.118562 | 1.016864 | 1.120341 |

###### *3.1.3.1 Mixing matrix age group adaptation*

Age-structured dynamic transmission epidemiology models (DTMs) with discrete age groups are often used when modeling continuous age is not feasible. One of the necessary components of the force of infection in these models is the age-by-age mixing matrix that gives the number of contacts per person per unit time for a person in age group $i$ with persons in age group $j$.

Published age-by-age mixing matrices are square, (often) symmetric matrices. The age groups are typically uniform in size (number of years), except, of course, for the oldest age group that has no upper bound (i.e., it is ∞). The age group boundaries of published mixing matrices seldom align completely with the age group boundaries of a DTM, and therefore the mixing matrix needs to be adapted for use in a DTM.

The method used here to adapt the mixing matrix is based on the conservation of contacts in the population. Therefore, the population age density is required, and should be of sufficient resolution to align with the age group boundaries of the initial mixing matrix and also with the age group boundaries of the model, and thus the final mixing matrix.

The initial mixing matrix is transformed in four steps to create the age group-adapted mixing matrix:

1. Multiply by the initial age group density to get contacts per unit time.
2. Expand into a larger matrix (more rows and columns) by partitioning some of the age groups into smaller age groups with their boundaries aligned with the final age groups; the proportions of contacts in the new partition exactly matches the age group density proportions in the new partition.
3. Contract into a smaller matrix (fewer rows and columns) by merging some of the age groups into larger age groups to align their boundaries with the final age groups.
4. Divide by the final age group density to get contacts per person per unit time.

Panel **A** of Figure S2 shows the adaptation used where the mixing matrix age groups are $\{(0,5), (5,10), (10, 15), \ldots, (70,75), (75, \infty)\}$ and the model age groups are $\{(0,2), (2,5), (5,18), (18,50), (50,65), (65, \infty)\}$. The union of the age group boundaries of the initial mixing and final model age groups gives the joint age groups for the expansion step, where the first age group is split into three, the second is split into two, and the last three are unchanged. In the contraction step, the third and fourth joint age groups are joined and the seventh and eighth joint age groups are joined.

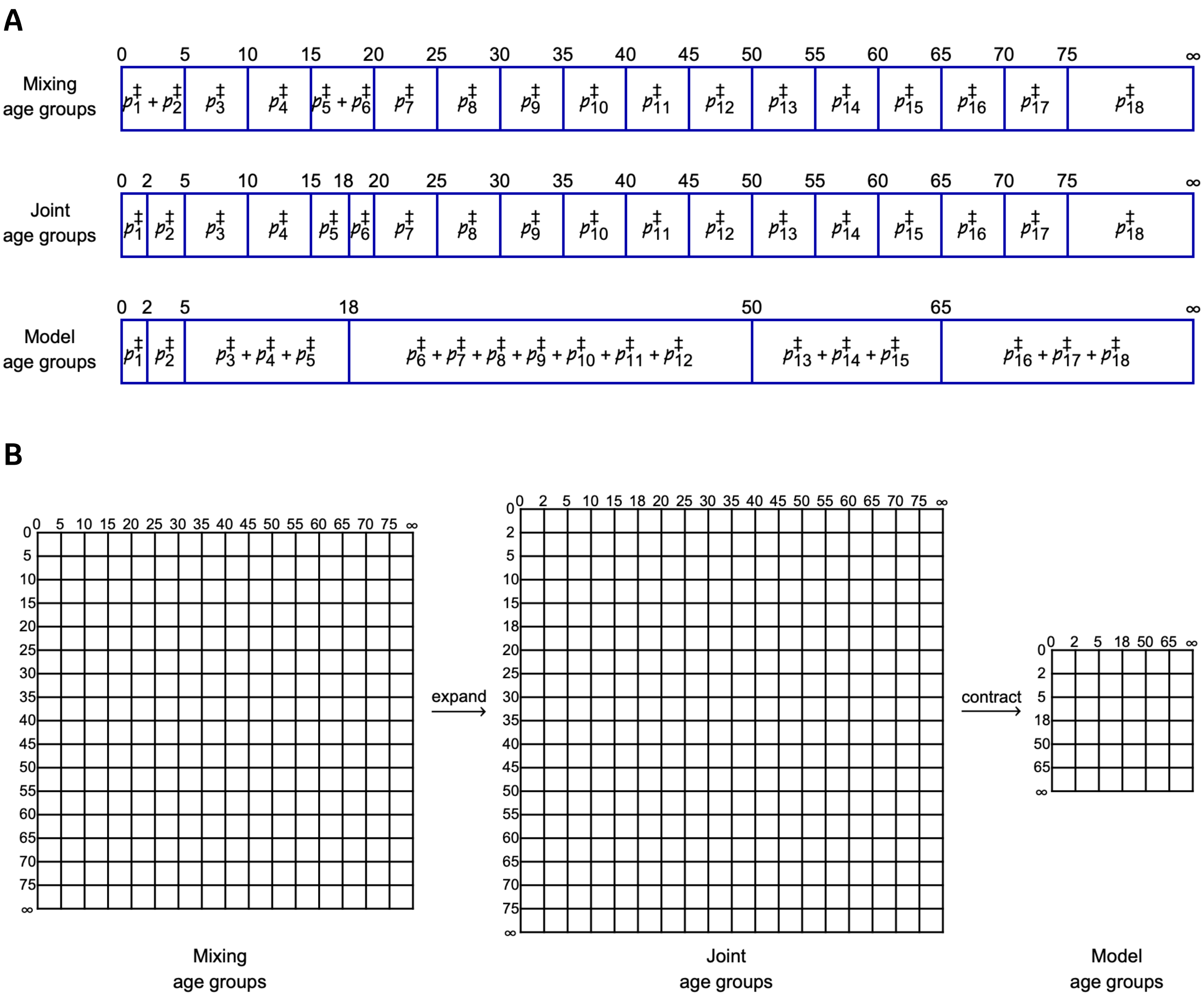

Figure S2. Step-by-step transformation of the mixing matrix, from the originally published age groups in Prem et al. (30) to the model age groups.

Panel **B** of Figure S2 shows steps 2 and 3 of the repartitioning of the mixing matrix. The population density for the joint age groups $\mathcal{p}_1^\ddagger, \dots, \mathcal{p}_{18}^\ddagger$ are used for the expansion step 2 and contraction step 3.

The expansion and contraction operations are most-easily carried out with linear algebra. Let $\mathcal{M}$ be the initial $n \times n$ mixing matrix, $\mathcal{M}^\ddagger$ be the joint $n^\ddagger \times n^\ddagger$ mixing matrix, and $\mathcal{M}^*$ be the final (model) $n^* \times n^*$ mixing matrix. Define the age group matrix $\mathcal{A}$ for the initial mixing matrix $\mathcal{M}$ with $a_{i,1} \le \text{age} < a_{i,2}$ for the $i$-th age group where $a_{1,1} = 0$, $a_{i,2} = a_{i+1,1}$, and $a_{n,2} = \infty$. Define also similar age group matrices $\mathcal{A}^\ddagger$ for $\mathcal{M}^\ddagger$ and $\mathcal{A}^*$ for $\mathcal{M}^*$.

Furthermore, let $\mathcal{p}$ be the initial population density vector of length $n$, $\mathcal{p}^\ddagger$ be the joint population density vector of length $n^\ddagger$, and $\mathcal{p}^*$ be the final (model) population density vector of length $n^*$. The following general relation holds

$$\sum_{i=1}^{n} \mathscr{p}_i = \sum_{i=1}^{n^\ddagger} \mathscr{p}_i^\ddagger = \sum_{i=1}^{n^*} \mathscr{p}_i^*$$

and, for the specific example above, these individual relations hold

$$\mathscr{p}_1 = \mathscr{p}_1^\ddagger + \mathscr{p}_2^\ddagger$$
$$\mathscr{p}_2 = \mathscr{p}_3^\ddagger$$
$$\mathscr{p}_3 = \mathscr{p}_4^\ddagger$$
$$\mathscr{p}_4 = \mathscr{p}_5^\ddagger + \mathscr{p}_6^\ddagger$$
$$\mathscr{p}_5 = \mathscr{p}_7^\ddagger$$
$$\mathscr{p}_6 = \mathscr{p}_8^\ddagger$$
$$\mathscr{p}_7 = \mathscr{p}_9^\ddagger$$
$$\mathscr{p}_8 = \mathscr{p}_{10}^\ddagger$$
$$\mathscr{p}_9 = \mathscr{p}_{11}^\ddagger$$
$$\mathscr{p}_{10} = \mathscr{p}_{12}^\ddagger$$
$$\mathscr{p}_{11} = \mathscr{p}_{13}^\ddagger$$
$$\mathscr{p}_{12} = \mathscr{p}_{14}^\ddagger$$
$$\mathscr{p}_{13} = \mathscr{p}_{15}^\ddagger$$
$$\mathscr{p}_{14} = \mathscr{p}_{16}^\ddagger$$
$$\mathscr{p}_{15} = \mathscr{p}_{17}^\ddagger$$
$$\mathscr{p}_{16} = \mathscr{p}_{18}^\ddagger$$
$$\mathscr{p}_1^\ddagger = \mathscr{p}_1^*$$
$$\mathscr{p}_2^\ddagger = \mathscr{p}_2^*$$
$$\mathscr{p}_3^\ddagger + \mathscr{p}_4^\ddagger + \mathscr{p}_5^\ddagger = \mathscr{p}_3^*$$
$$\mathscr{p}_6^\ddagger + \mathscr{p}_7^\ddagger + \mathscr{p}_8^\ddagger + \mathscr{p}_9^\ddagger + \mathscr{p}_{10}^\ddagger + \mathscr{p}_{11}^\ddagger + \mathscr{p}_{12}^\ddagger = \mathscr{p}_4^*$$
$$\mathscr{p}_{13}^\ddagger + \mathscr{p}_{14}^\ddagger + \mathscr{p}_{15}^\ddagger = \mathscr{p}_5^*$$
$$\mathscr{p}_{16}^\ddagger + \mathscr{p}_{17}^\ddagger + \mathscr{p}_{18}^\ddagger = \mathscr{p}_6^*$$

Now, define the mapping matrix $M^\ddagger$ from $\mathcal{A}$ to $\mathcal{A}^\ddagger$ where

$$m_{i,j}^\ddagger = \begin{cases} 1 & a_{i,1} \le a_{j,1}^\ddagger \wedge a_{j,2}^\ddagger \le a_{i,2} \\ 0 & \text{True} \end{cases} \qquad \text{with } 1 \le i \le n \text{ and } 1 <= j <= n^\ddagger$$

and likewise for $M^*$ from $\mathcal{A}^\ddagger$ to $\mathcal{A}^*$ where

$$m_{i,j}^\ddagger = \begin{cases} 1 & a_{i,1}^\ddagger \le a_{j,1}^* \wedge a_{j,2}^* \le a_{i,2}^\ddagger \\ 0 & \text{True} \end{cases} \qquad \text{with } 1 \le i \le n^\ddagger \text{ and } 1 <= j <= n^*$$

For step 1 of the algorithm, define the matrix of contacts per unit time $\mathcal{C}$ as

C = diag($\mathscr{p}$)·$M$

For the expansion step 2, define the weight matrix $W^\ddagger$

Proprietary

$$W^{\ddagger} = \left(\frac{\mathscr{p}^{\ddagger}}{\mathscr{p} \cdot M^{\ddagger}}\right) \cdot \left(\frac{\mathscr{p}^{\ddagger}}{\mathscr{p} \cdot M^{\ddagger}}\right)^{T}$$

The joint matrix of contacts per unit time $\mathcal{C}^{\ddagger}$ is defined as the Hadamard product

$$\mathcal{C}^{\ddagger} = W^{\ddagger}\left(M^{\ddagger^{T}} \cdot \mathcal{C} \cdot M^{\ddagger}\right)$$

For the contraction step 3, define the weight matrix $W^{*}$

$$W^{*} = \left(\frac{\mathscr{p}^{*}}{\mathscr{p}^{\ddagger} \cdot M^{*}}\right) \cdot \left(\frac{\mathscr{p}^{*}}{\mathscr{p}^{\ddagger} \cdot M^{*}}\right)^{T}$$

The model matrix of contacts per unit time $\mathcal{C}^{*}$ is defined as the Hadamard product

$$\mathcal{C}^{*} = W^{*}\left(M^{*^{T}} \cdot \mathcal{C}^{\ddagger} \cdot M^{*}\right)$$

The final step 4 gives the model matrix of contacts per person per unit time

$$\mathcal{M}^{*} = \mathrm{diag}\left(\frac{1}{\mathscr{p}^{*}}\right) \cdot \mathcal{C}^{*} \, .$$

#### 3.2 The epidemiologic model

The epidemiologic model distinguishes between pneumococcal carriage and disease. Carriage transmission is modeled via differential equations, and carriage may progress to a pneumococcal disease. This is modeled through a linear relation which depends on age, serotype, and vaccine-status. At some point, most children will acquire *Streptococcus pneumoniae* (SP). This colonization will typically reside asymptomatically in the nasopharynx for prolonged periods of time. Carriage does not always result in a pneumococcal disease. On the other hand, transmission dynamics revolve entirely around carriage transmission. Children younger than 5 years and adults 65 years or older are at increased risk for pneumococcal disease [31].

The model incorporates the occurrence of pneumococcal disease with carriage acquisition, including invasive pneumococcal disease (IPD), acute otitis media (AOM), and non-bacteremic pneumococcal pneumonia (NBPP). However, other manifestations (for example bacteremia or further classification of AOM, namely simple or recurrent) can be analyzed using this framework, which entails adjusting parameters relevant to the diseases under consideration.

ST classes (STCs) are summarized in Table 1 of the main text. Model age groups were chosen according to the available US data and vaccine recommendations. Of note is the age boundary of 18 years. The ABCs surveillance reports break the age groups at 18 (5-17, 18-34) for the IPD incidence and CFR estimates. On the other hand, vaccine recommendations for risk group start at 19 years of age (and not 18). The parameters are stratified according to the model age groups (as described above) to align with the data, hence we use 18 as an age-group boundary.

###### 3.2.1 Major modeling assumptions

1. Each new case of pneumococcal disease is assumed to develop from acquisition, based on the invasiveness or case-to-carrier ratio of the serotype. For example, serotype 1 (ST1), a common cause of IPD, is rarely observed in carrier state in the nasopharynx, implying its high invasiveness for IPD that results in fast development of IPD [32].
2. The model is not stratified based on risk categorization of the population. Therefore, risk-specific inputs are aggregated to represent the population-level estimates. For example, the risk-group specific vaccination rates and vaccine efficacies are used to estimate the corresponding values in the overall age groups 18-64 and 65+, without risk-based distinction. Proportions of the population attributed to healthy, chronic medical conditions, and immunocompromising conditions were obtained from Pelton *et al*. 2019 [33].
3. Since carriage is mostly asymptomatic and likely undetected at the time of vaccination, non-colonized as well as colonized individuals are vaccinated, according to the corresponding vaccination rate in the respective age group.
4. PPSV23 does not protect against carriage acquisition. Individuals who have received both vaccines are assumed to have protection against acquisition by the PCV.
5. Protection against disease development in individuals who have received both PCV and PPSV23 are assumed to have protection against disease as maximum VEd of the two vaccines.
6. We assumed that double- and triple-colonized individuals clear serotypes sequentially.
7. We assumed that a person can be colonized with at most three serotypes at a given time, and that there is no simultaneous transmission of three STCs.

8. Given the lack of detailed data to parameterize the model for all relevant serotypes, we grouped serotypes into 11 categories (or serotype classes) and assumed that parameter values (competition, acquisition probability, duration of carriage, and invasiveness) do not change within a serotype class. Modeling papers which include multiple STs typically aggregate them into groups in a similar fashion for tractability. For example, Choi, et al. [34] grouped STs by the vaccines. The first group included PCV7 STs, second contained the additional PCV13 STs (except ST1 and ST3), and the third group included the additional PCV15 or PCV20 STs depending on the model used. All the remaining STs comprised the final group. Parameter values (such as vaccine efficacy (VE)) were assumed constant across STs in the same group. In Wasserman, et al. [24], STs were divided into 5 serotype groups: (i) ST19A, (ii) ST3, (iii) ST1,5,7F,6A, (iv) PCV7-STs, (v) NVTs. Each serotype group was assumed to have a common value for probability of IPD given carriage acquisition, duration of carriage, duration of immunity, PCV13 VE against IPD, and PCV13 VE against carriage. Similarly, Melegaro,

et al. [4] divided STs into two broad groups, vaccine-type (VT) and nonvaccine-type (NVT) and a single value for each of age-specific proportion of population that recovers from infection, competition parameters, VE against VT carriage, and age-specific force of infection was assumed for the entire VT (respectively, NVT) group. STs are divided into six classes (low, medium and high prevalence VT/NVT) in Bottomley, et al. [12], where all STs from the same class are assumed to be acquired and cleared at the same rate.

##### 3.2.2 Epidemiologic model structure

The deterministic, age-structured, multi-serotype SP transmission model is constructed by dividing the population into age-specific compartments based on their pneumococcal carriage status, disease status, and vaccine protection status, describing the associated carriage transmission dynamics and disease progression in the presence of age- and serotype-specific pneumococcal vaccines.

###### *3.2.2.1 Description of model compartments*

Model compartments are described in detail below. Model variables and epidemiologic parameters are summarized in Table S5 & Table S6.

Table S5. Description of model variables. Subscript $\sigma$ represents vaccination status v (vaccinated with a PCV), w (vaccinated with PPSV23) or vw (vaccinated with both PCV and PPSV23).

| Variable | Description |
|---|---|
| $\mathcal{N}_{u,a}$ | Unvaccinated non-colonized individuals in age group *a*. |
| $\mathcal{N}_{\sigma,a}$ | Non-colonized individuals in age group *a* with vaccination status $\sigma$. |
| $\mathcal{C}_{u,a,i}$ | Unvaccinated individuals in age group *a* colonized with serotype class *i*. |
| $\mathcal{C}_{\sigma,a,i}$ | Individuals in age group *a* with vaccination status $\sigma$ colonized with serotype class *i*. |
| $\mathcal{CC}_{u,a,i,j}$ | Unvaccinated individuals in age group *a* co-colonized with serotype classes *i* and *j*. |
| $\mathcal{CC}_{\sigma,a,i,j}$ | Individuals in age group *a* with vaccination status $\sigma$ co-colonized with serotype classes *i* and *j*. |

| | |
|---|---|
| $\mathcal{CCC}_{u,a,i,j,k}$ | Unvaccinated individuals in age group *a* co-colonized with serotype classes *i, j* and *k*. |
| $\mathcal{CCC}_{\sigma,a,i,j,k}$ | Individuals in age group *a* with vaccination status $\sigma$ co-colonized with serotype classes *i, j* and *k*. |
| $\mathcal{M}_{\sigma,a}$ | Non-colonized individuals in age group *a* with waned $\sigma$ −vaccine protection. |
| $\mathcal{Q}_{\sigma,a,i}$ | Individuals in age group *a* colonized with serotype class *i* with waned $\sigma$ −vaccine protection. |
| $\mathcal{QQ}_{\sigma,a,i,j}$ | Individuals in age group *a* co-colonized with serotype classes *i* and *j* with waned $\sigma$ −vaccine protection. |
| $\mathcal{QQQ}_{\sigma,a,i,j,k}$ | Individuals in age group *a* co-colonized with serotype classes *i, j* and *k* with waned $\sigma$ −vaccine protection. |
| $\mathcal{D}_{\sigma,a,i}$ | Individuals in age group *a* with vaccination status $\sigma$ and pneumococcal disease with serotype class *i.* |
| $\mathcal{P}_a$ | Total population in age group *a*. |

Table S6. Description of epidemiologic model parameters.

| **Parameter** | **Description** |
|---|---|
| $c_{a,b}$ | Average number of contacts an individual in age group *a* makes with an individual in age group *b* per year. |
| $\beta_{a,i}$ | Probability of acquisition of carriage of serotype class *i* per contact in age group *a*. |
| $\gamma_{\sigma,a,i}$ | Serotype class *i* clearance rate of $\sigma$-vaccinated individuals in age group *a*. |
| $\theta_{i,j}$ | Competition parameter if currently colonized with serotype class *i* (risk reduction for second carriage with *j*) |
| $\theta_{i,j,k}$ | Competition parameter if currently co-colonized with serotype classes *i* and *j* (risk reduction for acquisition of a third serotype *k*) |
| $\theta\theta_{i,j,k}$ | Competition parameter if currently colonized with serotype class *i* (risk reduction for simultaneous acquisition of two serotypes *j* and *k*) |

| | |
|---|---|
| $\rho_{a,i}$ | Case-to-carrier ratio (the probability of developing a pneumococcal disease given carriage with serotype class *i* in age group *a*). |
| $\epsilon_{\sigma,a,i}$ | σ-vaccine efficacy against carriage acquisition of serotype class *i* in age group *a.* |
| $\epsilon_{D,\sigma,a,i}$ | σ-vaccine efficacy against developing pneumococcal disease due to serotype class *i* in age group *a.* |
| $\omega_{\sigma,a}$ | σ-vaccine waning rate in age group *a.* |
| $\phi_{v,1}$ | Cohort vaccination coverage in the first age group of the model. |
| $\phi c_{v,a}, \phi c_{w,a}$ | Cohort vaccination coverage in age group *a* > 1 with a PCV and PPSV23 respectively. |
| $\phi cv_{w,a}, \phi cw_{v,a}$ | Cohort rate of PPSV23 (PCV) vaccination of those who were previously vaccinated with PCV (PPSV23). |
| $\psi_{v,a}, \psi_{w,a}$ | Continuous rate of vaccination of the vaccine-naïve individuals in age group *a* with a PCV or PPSV23 respectively. |
| $\psi v_{w,a}, \psi w_{v,a}$ | Continuous rate of PPSV23 (PCV) vaccination of those who were previously vaccinated with PCV (PPSV23). |
| $\alpha_{a,b}$ | Transmissibility of carriage from age group *b* to age group *a*. |

##### ***3.2.2.2 Carriage Transmission Dynamics***

###### ***3.2.2.2.1*** Unvaccinated individuals $\mathcal{N}_{u,a}, \mathcal{C}_{u,a,i}, \mathcal{CC}_{u,a,i,j}$ and $\mathcal{CCC}_{u,a,i,j,k}$

Newborns enter the model in first age group at a constant rate Λ. A fraction (1 – $\phi_{v,1}$) that does not receive vaccination (assuming childhood vaccination program) joins the compartment of unvaccinated non-colonized individuals, $\mathcal{N}_{u,1}$. Unvaccinated non-colonized individuals $\mathcal{N}_{u,a}$ in age group *a* may acquire single carriage with STC *i* at a force of colonization $\lambda_{a,i}$, moving to the compartments $\mathcal{C}_{u,a,i}$ or simultaneous co-colonization with two serotypes *i* and *j*, at a rate $\lambda_{a,i,j}$, moving to the compartments $\mathcal{CC}_{u,a,i,j}$. Individuals with single carriage (ST *i*) may acquire a second ST j, moving from $\mathcal{C}_{u,a,i}$ to $\mathcal{CC}_{u,a,i,j}$ at a rate $\theta_{i,j}(\lambda_{a,j} + \lambda_{a,i,j})$. Rate of carriage acquisition of a ST *j* in presence of a currently carried ST *i* is reduced due to ST competition $\theta_{i,j}$ (< 1).

Singly or doubly colonized individuals may acquire triple colonization with STC *k* at a force of colonization $\theta\theta_{i,j,k}\lambda_{a,j,k}$ or $\theta_{i,j,k}\left(\lambda_{a,k}+\lambda_{a,i,k}+\lambda_{a,j,k}\right)$, respectively, joining the compartment $\mathcal{CCC}_{u,a,i,j,k}$ of triple colonization. In the case of acquiring two STs *j* and *k* in the presence of a single ST *i*, we assume that competition between *i* and *j* is independent of that between *i* and *k*. therefore, we assume that $\theta\theta_{i,j,k}=\theta_{i,j}\theta_{i,k}$ (impacting the transition from $\mathcal{C}_{u,a,i}$ to $\mathcal{CCC}_{u,a,i,j,k}$). On the other hand, competition parameter when there are two current STs *i* and *j* and a single ST *k* is invading (which applies to transition from $\mathcal{CC}_{u,a,i,j}$ to $\mathcal{CCC}_{u,a,i,j,k}$) the competition parameter (assuming similar independence between *i*-*k* competition and *j*-*k* competition) is given by $\theta_{i,j,k}=\theta_{i,k}\theta_{j,k}$.

Carriage clearance of a STC *i* (at rate $\gamma_{u,a,i}$) moves individuals from compartments of triple carriage to corresponding compartments of double carriage with remaining STCs, double to single, and single to non-colonized.

Unvaccinated individuals in an age group *a* may receive vaccination of a PCV or PPSV23 through cohort vaccination at an age boundary (at a rate $\phi c_{v,a}$ or $\phi c_{w,a}$, respectively) or through continuous vaccination (at a rate $\psi_{v,a}$ or $\psi_{w,a}$, respectively), leaving the unvaccinated compartments. Individuals who do not become colonized or get vaccinated remain in compartment $\mathcal{N}_{u,a}$.

###### *3.2.2.2.2* Vaccinated individuals $\mathcal{N}_{\sigma,a}, \mathcal{C}_{\sigma,a,i}, \mathcal{CC}_{\sigma,a,i,j}$ and $\mathcal{CCC}_{\sigma,a,i,j,k}, (\sigma=v,w,vw)$

Vaccinated non-colonized individuals $\mathcal{N}_{\sigma,a}$ in age group *a* may acquire single carriage with STC *i* at a force of colonization $\lambda v_{\sigma,a,i}$, moving to the compartments $\mathcal{C}_{\sigma,a,i}$ or simultaneous co-colonization with two serotypes *i* and *j*, at a rate $\lambda v_{\sigma,a,i,j}$, moving to the compartments $\mathcal{CC}_{\sigma,a,i,j}$. Those singly or doubly colonized individuals may acquire a third STC *k* at a force of colonization $\theta\theta_{i,j,k}\lambda v_{\sigma,a,j,k}$ or $\theta_{i,j,k}\left(\lambda v_{\sigma,a,k}+\lambda v_{\sigma,a,i,k}+\lambda v_{\sigma,a,j,k}\right)$, respectively, joining the compartment $\mathcal{CCC}_{\sigma,a,i,j,k}$ of triple colonization with vaccination status $\sigma$. Carriage clearance of a STC *i* (at rate $\gamma_{\sigma,a,i}$) moves individuals from compartments of triple carriage to corresponding compartments of double carriage with remaining STCs, double to single, and single to non-colonized.

PCV-vaccinated (respectively, PPSV23-vaccinated) individuals in an age group *a* may receive PPSV23 (respectively, PCV) vaccination through cohort vaccination at an age boundary at a rate $\phi cv_{w,a}$ (respectively, $\phi cw_{v,a}$) or through continuous vaccination at a rate $\psi v_{w,a}$ (respectively, $\psi w_{v,a}$), leaving the single vaccination compartments and joining the corresponding compartments with dual vaccination status $(\sigma=vw)$.

Waning of vaccine protection moves individuals out of vaccine-protected compartments at waning rate $\omega_{\sigma,a}$.

###### *3.2.2.2.3* Individuals with waned vaccine protection $\mathcal{M}_{\sigma,a}$, $\mathcal{Q}_{\sigma,a,i}$, $\mathcal{QQ}_{\sigma,a,i,j}$, $\mathcal{QQQ}_{\sigma,a,i,j,k}$, $(\sigma = v, w, vw)$

These compartments are populated by waning of the vaccine protection in the corresponding vaccinated compartments. Uncolonized individuals move from compartments $\mathcal{N}_{\sigma,a}$ into $\mathcal{M}_{\sigma,a}$ and colonized individuals from $\mathcal{C}_{\sigma,a,i}$, $\mathcal{CC}_{\sigma,a,i,j}$ and $\mathcal{CCC}_{\sigma,a,i,j,k}$ respectively move into $\mathcal{Q}_{\sigma,a,i}$, $\mathcal{QQ}_{\sigma,a,i,j}$, $\mathcal{QQQ}_{\sigma,a,i,j,k}$. The purpose of subscript $\sigma$ in the notation for waned compartments is to keep track of history of the corresponding vaccination status $(v, w, vw)$. Since there is no more vaccine protection, for each vaccination status $(v, w, or\ vw)$ the dynamics within waned compartments pertaining to carriage acquisition and clearance are analogous to those of the unvaccinated compartments.

###### *3.2.2.3 Pneumococcal Disease*

###### *3.2.2.3.1* **Individuals with pneumococcal disease:** $\mathcal{D}_{\sigma,a,i}$ $(\sigma = v, w, vw)$

Individuals with a pneumococcal disease due to serotype *i* are represented by $\mathcal{D}_{\sigma,a,i}$. Development of a pneumococcal disease *D* (IPD, NBPP, AOM etc.) in the $\sigma$-vaccinated, colonized individuals, is diminished by a factor $\left(1 - \epsilon_{D,\sigma,a,i}\right)$ due to the vaccine efficacy $\epsilon_{D,\sigma,a,i}$ against the particular disease by ST *i* in age group *a.* These individuals are part of the pool of i-colonized individuals with the same vaccination status. To estimate disease burden, we look at the fraction of colonized individuals who develop the pneumococcal disease, determined by the disease-specific case-to-carrier ratio, $\rho_{a,i}$. Thus, upon recovery from disease these individuals remain in the pool of colonized individuals, do not lose their vaccine status, and the immunity is identical to someone who is colonized.

##### 3.2.3 Model flowchart

The model compartments and transitions are described by the flowchart in Figure S3 (unvaccinated individuals), Figure S4 (vaccinated individuals) and Figure S5 (individuals with waned vaccine).

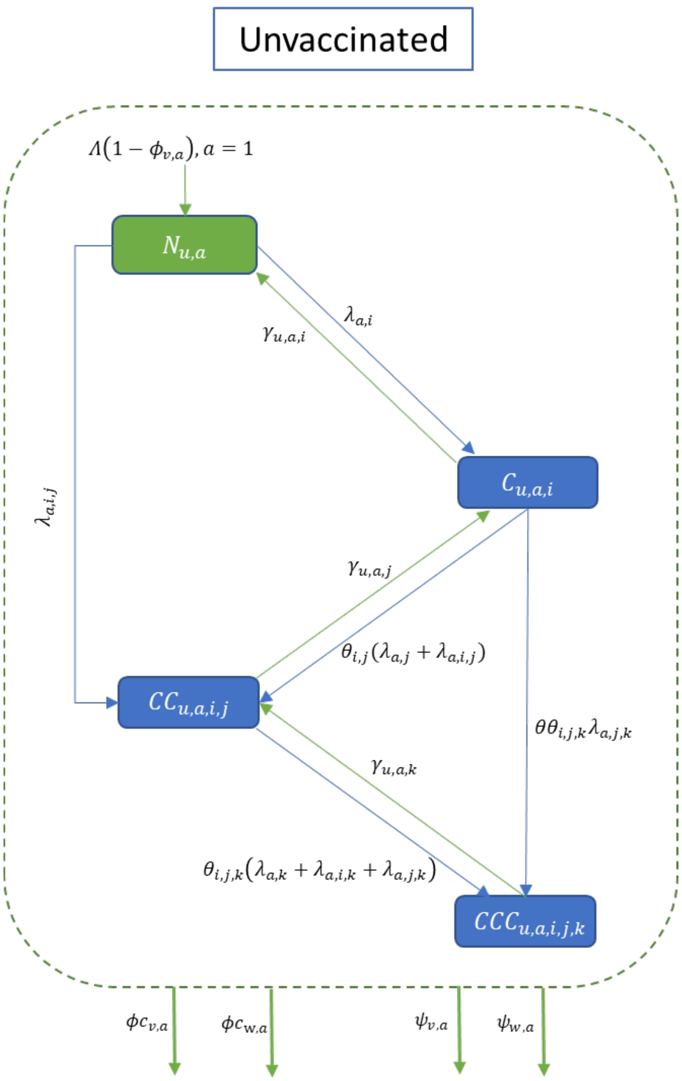

Figure S3. Model flowchart describing carriage transmission and vaccination dynamics in unvaccinated compartments.

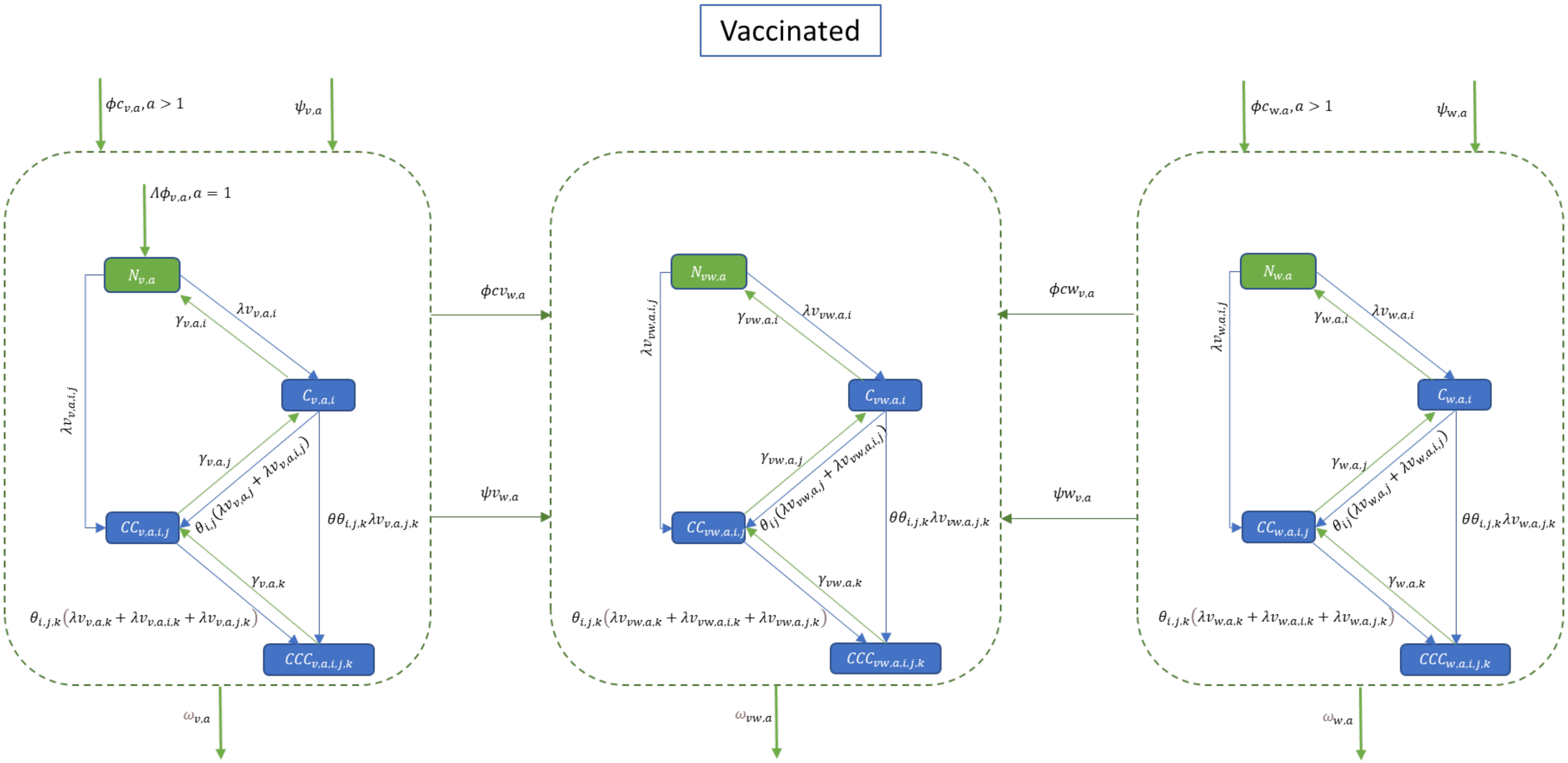

Figure S4. Model flowchart describing carriage transmission and vaccination/waning dynamics among vaccinated compartments.

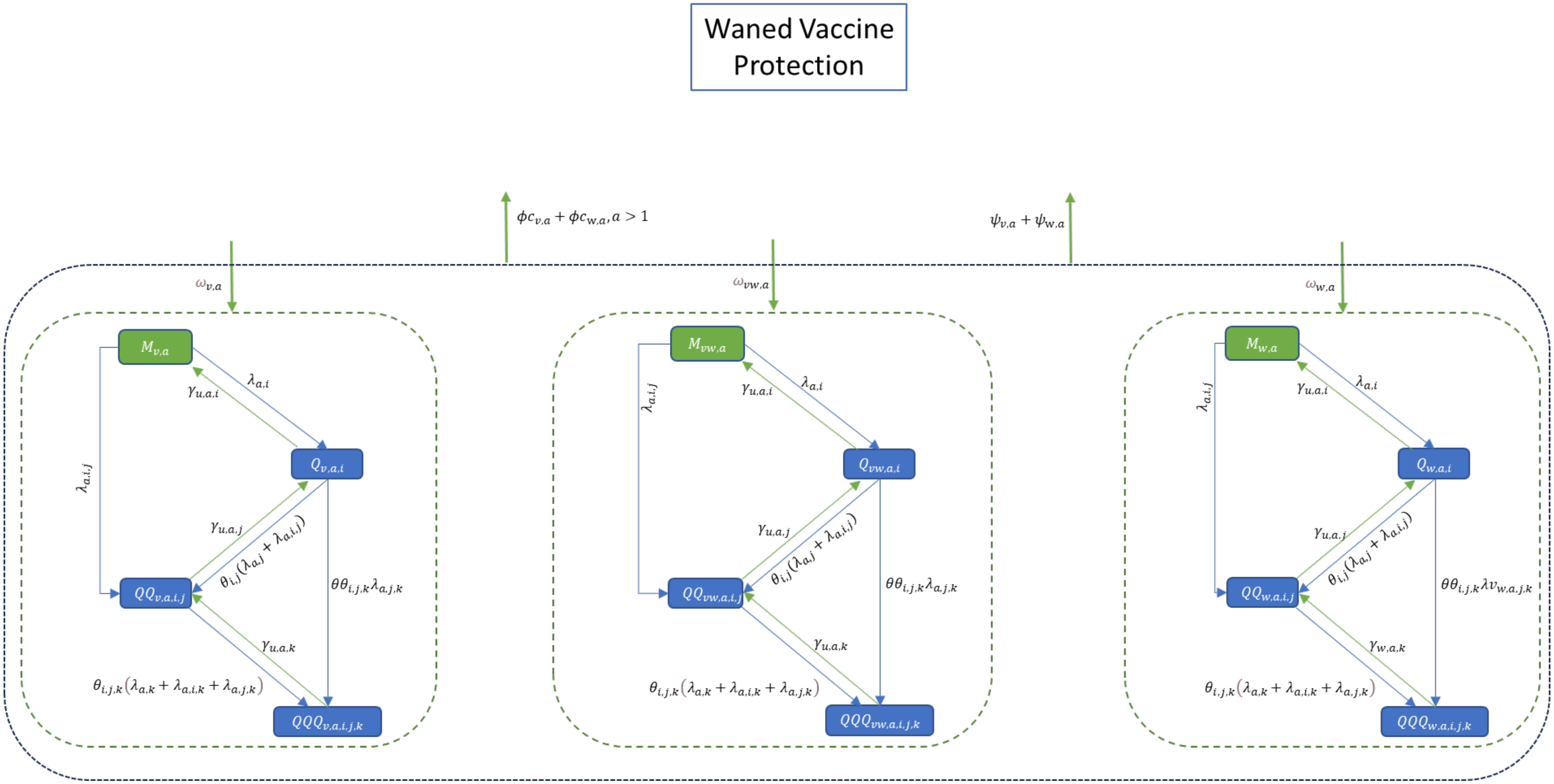

Figure S5. Model flowchart describing carriage transmission and vaccination dynamics among compartments with waned vaccine protection.

##### 3.2.4 Model equations

The model consists of the following ordinary differential equations (ODEs), describing the dynamics associated with each of the compartments described in the flowchart (Figure S3-S5), where $\delta_{x,y}$ is Kronecker delta ($\delta_{x,y} = 1\ if\ x = y, 0\ if\ x \neq\ y$) and prime represents differentiation with respect to time:

$$\mathcal{N}'_{u,a} = \delta_{1,a}\Lambda(1-\phi_{v,a}) + (1-\delta_{1,a})(1-\phi c_{v,a} - \phi c_{w,a})m_{a-1}\mathcal{N}_{u,a-1} + \sum_{i=1}^{nS}\gamma_{u,a,i}\mathcal{C}_{u,a,i}$$

$$-\left(m_a + \psi_{v,a} + \psi_{w,a} + \sum_{i=1}^{nS}\lambda_{a,i} + \sum_{i=1}^{nS-1}\sum_{j=i+1}^{nS}\lambda_{a,i,j} + \mu_a\right)\mathcal{N}_{u,a},$$

$$\mathcal{N}'_{v,a} = \delta_{1,a}\Lambda\phi_{v,a} + (1-\delta_{1,a})m_{a-1}(1-\phi cv_{w,a})\mathcal{N}_{v,a-1} + (1-\delta_{1,a})\phi c_{v,a}m_{a-1}\mathcal{N}_{u,a-1} + \psi_{v,a}\mathcal{N}_{u,a}$$

$$+\sum_{i=1}^{nS}\gamma_{v,a,i}\mathcal{C}_{v,a,i} - \left(m_a + \mu_a + \omega_{v,a} + \psi v_{w,a} + \sum_{i=1}^{nS}\lambda v_{v,a,i} + \sum_{i=1}^{nS-1}\sum_{j=i+1}^{nS}\lambda v_{v,a,i,j}\right)\mathcal{N}_{v,a},$$

$$\mathcal{N}_{w,a}{}' = (1-\delta_{1,a})m_{a-1}\left[(1-\phi cw_{v,a})\mathcal{N}_{w,a-1} + \phi c_{w,a}\mathcal{N}_{u,a-1}\right] + \psi_{w,a}\mathcal{N}_{u,a} + \sum_{i=1}^{\mathrm{nS}}\gamma_{w,a,i}\mathcal{C}_{w,a,i}$$

$$-\left(m_a + \mu_a + \omega_{w,a} + \psi w_{v,a} + \sum_{i=1}^{\mathrm{nS}}\lambda_{a,i} + \sum_{i=1}^{nS-1}\sum_{j=i+1}^{nS}\lambda_{a,i,j}\right)\mathcal{N}_{w,a},$$

$$\mathcal{N}_{vw,a}{}' = (1-\delta_{1,a})m_{a-1}(\mathcal{N}_{vw,a-1} + \phi cv_{w,a}\mathcal{N}_{v,a-1} + \phi cw_{v,a}\mathcal{N}_{w,a-1}) + \psi v_{w,a}\mathcal{N}_{v,a} + \psi w_{v,a}\mathcal{N}_{w,a}$$

$$+\sum_{i=1}^{\mathrm{nS}}\gamma_{vw,a,i}\mathcal{C}_{vw,a,i} - \left(m_a + \mu_a + \omega_{vw,a} + \sum_{i=1}^{\mathrm{nS}}\lambda v_{v,a,i} + \sum_{i=1}^{\mathrm{nS}-1}\sum_{j=i+1}^{\mathrm{nS}}\lambda v_{v,a,i,j}\right)\mathcal{N}_{vw,a},$$

$$\mathcal{C}'_{u,a,i} = \left(1-\delta_{1,a}\right)\left(1-\phi c_{v,a} - \phi c_{vw,a}\right)m_{a-1}\mathcal{C}_{u,a-1,i} + \lambda_{a,i}\mathcal{N}_{u,a} + \sum_{j=i+1}^{\mathrm{nS}}\gamma_{u,a,j}\mathcal{CC}_{u,a,i,j}$$

$$+\sum_{j=1}^{\mathrm{i}-1}\gamma_{u,a,j}\mathcal{CC}_{u,a,j,i}$$

$$-\left(m_a + \mu_a + \psi_{v,a} + \psi_{w,a} + \gamma_{u,a,i} + \sum_{\substack{j=1,\\ j\neq i}}^{nS}\theta_{i,j}\lambda_{a,j} + \sum_{j=i+1}^{nS}\theta_{i,j}\lambda_{a,i,j} + \sum_{j=1}^{i-1}\theta_{i,j}\lambda_{a,j,i}\right.$$

$$\left.+\sum_{\substack{j=1,\\ j\neq i}}^{nS-1}\sum_{\substack{k=j+1,\\ k\neq i}}^{nS}\theta\theta_{i,j,k}\lambda_{a,j,k}\right)\mathcal{C}_{u,a,i},$$

$$
\begin{aligned}
\mathcal{C}'_{v,a,i} = {} & \left(1-\delta_{1,a}\right) m_{a-1}\left[\left(1-\phi c v_{w,a}\right)\mathcal{C}_{v,a-1,i} + \phi c_{v,a} m_{a-1}\mathcal{C}_{u,a-1,i}\right] + \psi_{v,a}\mathcal{C}_{u,a,i} + \lambda v_{v,a,i}\mathcal{N}_{v,a} \\
& + \sum_{j=i+1}^{\mathrm{nS}} \gamma_{v,a,j}\mathcal{CC}_{v,a,i,j} + \sum_{j=1}^{\mathrm{i-1}} \gamma_{v,a,j}\mathcal{CC}_{v,a,j,i} \\
& - \left( m_a + \mu_a + \omega_{v,a} + \psi v_{w,a} + \gamma_{v,a,i} + \sum_{\substack{j=1,\\ j\neq i}}^{\mathrm{nS}} \theta_{i,j}\lambda v_{v,a,j} + \sum_{j=i+1}^{\mathrm{nS}} \theta_{i,j}\lambda v_{v,a,i,j} \right. \\
& \left. + \sum_{j=1}^{\mathrm{i-1}} \theta_{i,j}\lambda v_{v,a,j,i} + \sum_{\substack{j=1,\\ j\neq i}}^{nS-1} \sum_{\substack{k=j+1,\\ k\neq i}}^{nS} \theta\theta_{i,j,k}\lambda v_{v,a,j,k} \right) \mathcal{C}_{v,a,i},
\end{aligned}
$$

$$
\begin{aligned}
\mathcal{C}_{w,a,i}{}' = {} & (1-\delta_{1,a}) m_{a-1}\left[(1-\phi c w_{v,a})\mathcal{C}_{w,a-1,i} + \phi c_{w,a}\mathcal{C}_{u,a-1,i}\right] + \lambda v_{w,a,i}\mathcal{N}_{w,a} + \psi_{w,a}\mathcal{C}_{u,a,i} \\
& + \sum_{j=i+1}^{\mathrm{nS}} \gamma_{w,a,j}\mathcal{CC}_{w,a,i,j} + \sum_{j=1}^{\mathrm{i-1}} \gamma_{w,a,j}\mathcal{CC}_{w,a,j,i} \\
& - \left( m_a + \mu_a + \omega_{w,a} + \psi w_{v,a} + \gamma_{w,a,i} + \sum_{\substack{j=1,\\ j\neq i}}^{\mathrm{nS}} \theta_{i,j}\lambda v_{w,a,j} + \sum_{j=i+1}^{\mathrm{nS}} \theta_{i,j}\lambda v_{w,a,i,j} \right. \\
& \left. + \sum_{j=1}^{\mathrm{i-1}} \theta_{i,j}\lambda v_{w,a,j,i} + \sum_{\substack{j=1,\\ j\neq i}}^{nS-1} \sum_{\substack{k=j+1,\\ k\neq i}}^{nS} \theta\theta_{i,j,k}\lambda v_{w,a,j,k} \right) \mathcal{C}_{w,a,i},
\end{aligned}
$$

$$
\begin{aligned}
\mathcal{C}_{vw,a,i}{}' = {} & (1-\delta_{1,a}) m_{a-1}(\mathcal{C}_{vw,a-1,i} + \phi c v_{w,a}\mathcal{C}_{v,a-1,i} + \phi c w_{v,a}\mathcal{C}_{w,a-1,i}) + \lambda v_{v,a,i}\mathcal{N}_{vw,a} + \psi v_{w,a}\mathcal{C}_{v,a,i} \\
& + \psi w_{v,a}\mathcal{C}_{w,a,i} + \sum_{j=i+1}^{\mathrm{nS}} \gamma_{vw,a,j}\mathcal{CC}_{vw,a,i,j} + \sum_{j=1}^{\mathrm{i-1}} \gamma_{vw,a,j}\mathcal{CC}_{vw,a,j,i} \\
& - \left( m_a + \mu_a + \omega_{vw,a} + \gamma_{vw,a,i} + \sum_{\substack{j=1,\\ j\neq i}}^{\mathrm{nS}} \theta_{i,j}\lambda v_{vw,a,j} + \sum_{j=i+1}^{\mathrm{nS}} \theta_{i,j}\lambda v_{vw,a,i,j} \right. \\
& \left. + \sum_{j=1}^{\mathrm{i-1}} \theta_{i,j}\lambda v_{vw,a,j,i} + \sum_{\substack{j=1,\\ j\neq i}}^{nS-1} \sum_{\substack{k=j+1,\\ k\neq i}}^{nS} \theta\theta_{i,j,k}\lambda v_{vw,a,j,k} \right) \mathcal{C}_{vw,a,i},
\end{aligned}
$$

$$\mathcal{CC}'_{u,a,i,j} = (1-\delta_{1,a})[1-(\phi c_{v,a}+\phi c_{w,a})]m_{a-1}\mathcal{CC}_{u,a-1,i,j} + \theta_{i,j}(\lambda_{a,j}+\lambda_{a,i,j})\mathcal{C}_{u,a,i}$$

$$+\theta_{j,i}(\lambda_{a,i}+\lambda_{a,i,j})\mathcal{C}_{u,a,j} + \lambda_{a,i,j}\mathcal{N}_{u,a} + \sum_{k=j+1}^{\mathrm{nS}}\gamma_{u,a,k}\mathcal{CCC}_{u,a,i,j,k} + \sum_{k=i+1}^{\mathrm{j}-1}\gamma_{u,a,k}\mathcal{CCC}_{u,a,i,k,j}$$

$$+\sum_{k=1}^{i-1}\gamma_{u,a,k}\mathcal{CCC}_{u,a,k,i,j}$$

$$-\left(m_a+\mu_a+\gamma_{u,a,i}+\gamma_{u,a,j}+\psi_{v,a}+\psi_{w,a}+\sum_{\substack{k=1,\\k\neq i,k\neq j}}^{\mathrm{nS}}\theta_{i,j,k}\lambda_{a,k}+\sum_{\substack{k=1,\\k>i,k\neq j}}^{\mathrm{nS}}\theta_{i,j,k}\lambda_{a,i,k}\right.$$

$$\left.+\sum_{\substack{k=1,\\k<i,k\neq j}}^{\mathrm{nS}}\theta_{i,j,k}\lambda_{a,k,i}+\sum_{\substack{k=1,\\k\neq i,k>j}}^{\mathrm{nS}}\theta_{i,j,k}\lambda_{a,j,k}+\sum_{\substack{k=1,\\k\neq i,k<j}}^{\mathrm{nS}}\theta_{i,j,k}\lambda_{a,k,j}\right)\mathcal{CC}_{u,a,i,j},$$

$$\mathcal{CC}'_{v,a,i,j} = (1-\delta_{1,a})m_{a-1}(1-\phi c v_{w,a})\mathcal{CC}_{v,a-1,i,j} + (1-\delta_{1,a})\phi c_{v,a}\mathcal{CC}_{u,a,i,j} + \psi_{v,a}\mathcal{CC}_{u,a,i,j}$$

$$+\theta_{i,j}(\lambda v_{v,a,j}+\lambda v_{v,a,i,j})\mathcal{C}_{v,a,i} + \theta_{j,i}(\lambda v_{v,a,i}+\lambda v_{v,a,i,j})\mathcal{C}_{v,a,j} + \lambda v_{v,a,i,j}\mathcal{N}_{v,a}$$

$$+\sum_{k=j+1}^{nS}\gamma_{v,a,k}\mathcal{CCC}_{v,a,i,j,k} + \sum_{k=i+1}^{j-1}\gamma_{v,a,k}\mathcal{CCC}_{v,a,i,k,j} + \sum_{k=1}^{i-1}\gamma_{v,a,k}\mathcal{CCC}_{v,a,k,i,j}$$

$$-\left(m_a+\mu_a+\omega_{v,a}+\psi v_{w,a}+\gamma_{v,a,i}+\gamma_{v,a,j}+\sum_{\substack{k=1,\\k\neq i,k\neq j}}^{nS}\theta_{i,j,k}\lambda v_{v,a,k}\right.$$

$$+\sum_{\substack{k=1,\\k>i,k\neq j}}^{nS}\theta_{i,j,k}\lambda v_{v,a,i,k}+\sum_{\substack{k=1,\\k<i,k\neq j}}^{nS}\theta_{i,j,k}\lambda v_{v,a,k,i}+\sum_{\substack{k=1,\\k\neq i,k>j}}^{nS}\theta_{i,j,k}\lambda v_{v,a,j,k}$$

$$\left.+\sum_{\substack{k=1,\\k\neq i,k<j}}^{nS}\theta_{i,j,k}\lambda v_{v,a,k,j}\right)\mathcal{CC}_{v,a,i,j},$$

Proprietary

$$\mathcal{CC}'_{w,a,i,j} = \left(1-\delta_{1,a}\right)m_{a-1}(1-\phi cw_{v,a})\mathcal{CC}_{w,a-1,i,j} + \left(1-\delta_{1,a}\right)\phi c_{w,a}\mathcal{CC}_{u,a,i,j} + \psi_{v,a}\mathcal{CC}_{u,a,i,j}$$
$$+ \theta_{i,j}\left(\lambda v_{w,a,j} + \lambda v_{w,a,i,j}\right)\mathcal{C}_{w,a,i} + \theta_{j,i}\left(\lambda v_{w,a,i} + \lambda v_{w,a,i,j}\right)\mathcal{C}_{w,a,j} + \lambda v_{w,a,i,j}\mathcal{N}_{w,a}$$
$$+ \sum_{k=j+1}^{nS} \gamma_{w,a,k}\mathcal{CCC}_{w,a,i,j,k} + \sum_{k=i+1}^{j-1} \gamma_{w,a,k}\mathcal{CCC}_{w,a,i,k,j} + \sum_{k=1}^{i-1} \gamma_{w,a,k}\mathcal{CCC}_{w,a,k,i,j}$$
$$-\left( m_a + \mu_a + \omega_{w,a} + \psi w_{v,a} + \gamma_{w,a,i} + \gamma_{w,a,j} + \sum_{\substack{k=1,\\ k\neq i, k\neq j}}^{nS} \theta_{i,j,k}\lambda v_{w,a,k} \right.$$
$$+ \sum_{\substack{k=1,\\ k>i, k\neq j}}^{nS} \theta_{i,j,k}\lambda v_{w,a,i,k} + \sum_{\substack{k=1,\\ k<i, k\neq j}}^{nS} \theta_{i,j,k}\lambda v_{w,a,k,i} + \sum_{\substack{k=1,\\ k\neq i, k>j}}^{nS} \theta_{i,j,k}\lambda v_{w,a,j,k}$$
$$\left. + \sum_{\substack{k=1,\\ k\neq i, k<j}}^{nS} \theta_{i,j,k}\lambda v_{w,a,k,j} \right)\mathcal{CC}_{w,a,i,j},$$

$$\mathcal{CC}'_{vw,a,i,j} = (1-\delta_{1,a})m_{a-1}(\mathcal{CC}_{vw,a-1,i,j} + \phi cv_{w,a}\mathcal{CC}_{v,a-1,i,j} + \phi cw_{v,a}\mathcal{CC}_{w,a-1,i,j}) + \psi v_{w,a}\mathcal{CC}_{v,a,i,j}$$
$$+ \psi w_{v,a}\mathcal{CC}_{w,a,i,j} + \theta_{i,j}(\lambda v_{v,a,j} + \lambda v_{v,a,i,j})\mathcal{C}_{vw,a,i} + \theta_{j,i}(\lambda v_{v,a,i} + \lambda v_{v,a,i,j})\mathcal{C}_{vw,a,j}$$
$$+ \lambda v_{v,a,i,j}\mathcal{N}_{vw,a} + \sum_{k=j+1}^{nS} \gamma_{vw,a,k}\mathcal{CCC}_{vw,a,i,j,k} + \sum_{k=i+1}^{j-1} \gamma_{vw,a,k}\mathcal{CCC}_{vw,a,i,k,j}$$
$$+ \sum_{k=1}^{i-1} \gamma_{vw,a,k}\mathcal{CCC}_{vw,a,k,i,j}$$
$$-\left( m_a + \mu_a + \omega_{vw,a} + \gamma_{vw,a,i} + \gamma_{vw,a,j} + \sum_{\substack{k=1,\\ k\neq i, k\neq j}}^{nS} \theta_{i,j,k}\lambda v_{vw,a,k} + \sum_{\substack{k=1,\\ k>i, k\neq j}}^{nS} \theta_{i,j,k}\lambda v_{vw,a,i,k} \right.$$
$$\left. + \sum_{\substack{k=1,\\ k<i, k\neq j}}^{nS} \theta_{i,j,k}\lambda v_{vw,a,k,i} + \sum_{\substack{k=1,\\ k\neq i, k>j}}^{nS} \theta_{i,j,k}\lambda v_{vw,a,j,k} + \sum_{\substack{k=1,\\ k\neq i, k<j}}^{nS} \theta_{i,j,k}\lambda v_{vw,a,k,j} \right)\mathcal{CC}_{vw,a,i,j},$$

$$\mathcal{CCC}'_{u,a,i,j,k} = (1-\delta_{1,a})(1-(\phi c_{v,a} + \phi c_{vw,a}))m_{a-1}\mathcal{CCC}_{u,a-1,i,j,k} + \theta_{j,k,i}(\lambda_{a,i} + \lambda_{a,i,j} + \lambda_{a,i,k})\mathcal{CC}_{u,a,j,k}$$
$$+ \theta_{i,k,j}(\lambda_{a,j} + \lambda_{a,i,j} + \lambda_{a,j,k})\mathcal{CC}_{u,a,i,k} + \theta_{i,j,k}(\lambda_{a,k} + \lambda_{a,i,k} + \lambda_{a,j,k})\mathcal{CC}_{u,a,i,j}$$
$$+ \theta\theta_{i,j,k}\lambda_{a,j,k}\mathcal{C}_{u,a,i} + \theta\theta_{j,i,k}\lambda_{a,i,k}\mathcal{C}_{u,a,j} + \theta\theta_{k,i,j}\lambda_{a,i,j}\mathcal{C}_{u,a,k} - (m_a + \mu_a + \psi_{v,a} + \psi_{w,a}$$
$$+ \gamma_{u,a,i} + \gamma_{u,a,j} + \gamma_{u,a,k})\mathcal{CCC}_{u,a,i,j,k},$$

$$\begin{aligned}\mathcal{CCC}'_{v,a,i,j,k} = {} & (1-\delta_{1,a})m_{a-1}\left((1-\phi cv_{w,a})\mathcal{CCC}_{v,a-1,i,j,k} + \phi c_{v,a}\mathcal{CCC}_{u,a-1,i,j,k}\right) + \psi_{v,a}\mathcal{CCC}_{u,a,i,j,k} \\ & + \theta_{j,k,i}\left(\lambda v_{v,a,i} + \lambda v_{v,a,i,j} + \lambda v_{v,a,i,k}\right)\mathcal{CC}_{v,a,j,k} \\ & + \theta_{i,k,j}\left(\lambda v_{v,a,j} + \lambda v_{v,a,i,j} + \lambda v_{v,a,j,k}\right)\mathcal{CC}_{v,a,i,k} \\ & + \theta_{i,j,k}\left(\lambda v_{v,a,k} + \lambda v_{v,a,i,k} + \lambda v_{v,a,j,k}\right)\mathcal{CC}_{v,a,i,j} + \theta\theta_{i,j,k}\lambda v_{v,a,j,k}\mathcal{C}_{v,a,i} \\ & + \theta\theta_{j,i,k}\lambda v_{v,a,i,k}\mathcal{C}_{v,a,j} + \theta\theta_{k,i,j}\lambda v_{v,a,i,j}\mathcal{C}_{v,a,k} \\ & - \left(m_a + \mu_a + \omega_{v,a} + \gamma_{v,a,i} + \gamma_{v,a,j} + \gamma_{v,a,k}\right)\mathcal{CCC}_{v,a,i,j,k},\end{aligned}$$

$$\begin{aligned}\mathcal{CCC}_{w,a,i,j,k}{}' = {} & (1-\delta_{1,a})m_{a-1}((1-\phi cw_{v,a})\mathcal{CCC}_{w,a-1,i,j,k} + \phi c_{w,a}\mathcal{CCC}_{u,a-1,i,j,k}) + \psi_{w,a}\mathcal{CCC}_{u,a,i,j,k} \\ & + \theta_{j,k,i}(\lambda_{a,i} + \lambda_{a,i,j} + \lambda_{a,i,k})\mathcal{CC}_{w,a,j,k} + \theta_{i,k,j}(\lambda_{a,j} + \lambda_{a,i,j} + \lambda_{a,j,k})\mathcal{CC}_{w,a,i,k} + \theta_{i,j,k}(\lambda_{a,k} \\ & + \lambda_{a,i,k} + \lambda_{a,j,k})\mathcal{CC}_{w,a,i,j} + \theta\theta_{i,j,k}\lambda_{a,j,k}\mathcal{C}_{w,a,i} + \theta\theta_{j,i,k}\lambda_{a,i,k}\mathcal{C}_{w,a,j} + \theta\theta_{k,i,j}\lambda_{a,i,j}\mathcal{C}_{w,a,k} \\ & - (m_a + \mu_a + \omega_{w,a} + \psi w_{v,a} + \gamma_{w,a,i} + \gamma_{w,a,j} + \gamma_{w,a,k})\mathcal{CCC}_{w,a,i,j,k},\end{aligned}$$

$$\begin{aligned}\mathcal{CCC}_{vw,a,i,j,k}{}' = {} & (1-\delta_{1,a})m_{a-1}(\mathcal{CCC}_{vw,a-1,i,j,k} + \phi cv_{w,a}\mathcal{CCC}_{v,a-1,i,j,k} + \phi cw_{v,a}\mathcal{CCC}_{w,a-1,i,j,k}) \\ & + \psi v_{w,a}\mathcal{CCC}_{v,a,i,j,k} + \psi w_{v,a}\mathcal{CCC}_{w,a,i,j,k} + \theta_{j,k,i}(\lambda v_{v,a,i} + \lambda v_{v,a,i,j} + \lambda v_{v,a,i,k})\mathcal{CC}_{vw,a,j,k} \\ & + \theta_{i,k,j}(\lambda v_{v,a,j} + \lambda v_{v,a,i,j} + \lambda v_{v,a,j,k})\mathcal{CC}_{vw,a,i,k} + \theta_{i,j,k}(\lambda v_{v,a,k} + \lambda v_{v,a,i,k} \\ & + \lambda v_{v,a,j,k})\mathcal{CC}_{vw,a,i,j} + \theta\theta_{i,j,k}\lambda v_{v,a,j,k}\mathcal{C}_{vw,a,i} + \theta\theta_{j,i,k}\lambda v_{v,a,i,k}\mathcal{C}_{vw,a,j} \\ & + \theta\theta_{k,i,j}\lambda v_{v,a,i,j}\mathcal{C}_{vw,a,k} - (m_a + \mu_a + \omega_{vw,a} + \gamma_{vw,a,i} + \gamma_{vw,a,j} + \gamma_{vw,a,k})\mathcal{CCC}_{vw,a,i,j,k},\end{aligned}$$

$$\begin{aligned}\mathcal{M}_{\sigma,a}{}' = {} & \omega_{\sigma,a}\mathcal{N}_{\sigma,a} + (1-\delta_{1,a})m_{a-1}(1-\phi c_{v,a} - \phi c_{w,a})\mathcal{M}_{\sigma,a-1} + \sum_{i=1}^{\mathrm{nS}}(\gamma_{u,a,i}\mathcal{Q}_{\sigma,a,i}) \\ & - \left(m_a + \mu_a + \psi_{v,a} + \psi_{w,a} + \sum_{i=1}^{\mathrm{nS}}\lambda_{a,i} + \sum_{i=1}^{\mathrm{nS}-1}\sum_{j=i+1}^{\mathrm{nS}}\lambda_{a,i,j}\right)\mathcal{M}_{\sigma,a}, \qquad \sigma = v, w, vw\end{aligned}$$

$$\begin{aligned}\mathcal{Q}_{\sigma,a,i}{}' = {} & \omega_{\sigma,a}\mathcal{C}_{\sigma,a,i} + (1-\delta_{1,a})m_{a-1}(1-\phi c_{v,a} - \phi c_{w,a})\mathcal{Q}_{\sigma,a-1,i} + \lambda_{a,i}\mathcal{M}_{\sigma,a} + \sum_{j=i+1}^{nS}\gamma_{u,a,j}\mathcal{QQ}_{\sigma,a,i,j} \\ & + \sum_{j=1}^{i-1}\gamma_{u,a,j}\mathcal{QQ}_{\sigma,a,j,i} \\ & - \left(m_a + \mu_a + \psi_{v,a} + \psi_{w,a} + \gamma_{u,a,i} + \sum_{\substack{j=1,\\ j\neq i}}^{nS}\theta_{i,j}\lambda_{a,j} + \sum_{j=i+1}^{nS}\theta_{i,j}\lambda_{a,i,j} + \sum_{j=1}^{i-1}\theta_{i,j}\lambda_{a,j,i}\right. \\ & \left. + \sum_{\substack{j=1,\\ j\neq i}}^{nS-1}\sum_{\substack{k=j+1,\\ k\neq i}}^{nS}\theta\theta_{i,j,k}\lambda_{a,j,k}\right)\mathcal{Q}_{\sigma,a,i}, \qquad \sigma = v, w, vw,\end{aligned}$$

$$\begin{aligned}
\mathcal{QQ}_{\sigma,a,i,j}{}' = {} & \omega_{\sigma,a}\mathcal{CC}_{\sigma,a,i,j} + (1-\delta_{1,a})m_{a-1}(1-\phi c_{v,a}-\phi c_{w,a})\mathcal{QQ}_{\sigma,a-1,i,j} + \theta_{i,j}(\lambda_{a,j}+\lambda_{a,i,j})\mathcal{Q}_{\sigma,a,i} \\
& + \theta_{j,i}(\lambda_{a,i}+\lambda_{a,i,j})\mathcal{Q}_{\sigma,a,j} + \lambda_{a,i,j}\mathcal{M}_{\sigma,a} + \sum_{k=j+1}^{nS}\gamma_{u,a,k}\mathcal{QQQ}_{\sigma,a,i,j,k} \\
& + \sum_{k=i+1}^{j-1}\gamma_{u,a,k}\mathcal{QQQ}_{\sigma,a,i,k,j} + \sum_{k=1}^{i-1}\gamma_{u,a,k}\mathcal{QQQ}_{\sigma,a,k,i,j} \\
& - \Bigg( m_a + \mu_a + \psi_{v,a} + \psi_{w,a} + \gamma_{u,a,i} + \gamma_{u,a,j} + \sum_{\substack{k=1,\\ k\neq i,k\neq j}}^{\mathrm{nS}} \theta_{i,j,k}\lambda_{a,k} + \sum_{\substack{k=1,\\ k>i,k\neq j}}^{\mathrm{nS}} \theta_{i,j,k}\lambda_{a,i,k} \\
& + \sum_{\substack{k=1,\\ k<i,k\neq j}}^{\mathrm{nS}} \theta_{i,j,k}\lambda_{a,k,i} + \sum_{\substack{k=1,\\ k\neq i,k>j}}^{\mathrm{nS}} \theta_{i,j,k}\lambda_{a,j,k} + \sum_{\substack{k=1,\\ k\neq i,k<j}}^{\mathrm{nS}} \theta_{i,j,k}\lambda_{a,k,j} \Bigg)\mathcal{QQ}_{\sigma,a,i,j}, \\
& \sigma = v, w, vw,
\end{aligned}$$

$$\begin{aligned}
\mathcal{QQQ}_{\sigma,a,i,j,k}{}' = {} & \omega_{\sigma,a}\mathcal{CCC}_{\sigma,a,i,j,k} + (1-\delta_{1,a})m_{a-1}(1-\phi c_{v,a}-\phi c_{w,a})\mathcal{QQQ}_{\sigma,a-1,i,j,k} + \theta_{j,k,i}(\lambda_{a,i}+\lambda_{a,i,j} \\
& + \lambda_{a,i,k})\mathcal{QQ}_{\sigma,a,j,k} + \theta_{i,k,j}(\lambda_{a,j}+\lambda_{a,i,j}+\lambda_{a,j,k})\mathcal{QQ}_{\sigma,a,i,k} + \theta_{i,j,k}(\lambda_{a,k}+\lambda_{a,i,k} \\
& + \lambda_{a,j,k})\mathcal{QQ}_{\sigma,a,i,j} + \theta\theta_{i,j,k}\lambda_{a,j,k}\mathcal{Q}_{\sigma,a,i} + \theta\theta_{j,i,k}\lambda_{a,i,k}\mathcal{Q}_{\sigma,a,j} + \theta\theta_{k,i,j}\lambda_{a,i,j}\mathcal{Q}_{\sigma,a,k} - (m_a \\
& + \mu_a + \psi_{v,a} + \psi_{w,a} + \gamma_{u,a,i} + \gamma_{u,a,j} + \gamma_{u,a,k})\mathcal{QQQ}_{\sigma,a,i,j,k}, \qquad \sigma = v, w, vw.
\end{aligned}$$

###### 3.2.4.1 *Forces of colonization $\lambda_{a,i}$, $\lambda_{a,i,j}$, $\lambda v_{\sigma,a,i}$ and $\lambda v_{\sigma,a,i,j}$*

The transmission rate of a single serotype *i* from an individual with nasopharyngeal carriage (or co- or triple carriage) of serotype *i* to an *i*-susceptible individual (per capita force of colonization) is age- and time-dependent. It also depends on mixing patterns among individuals of different age groups (incorporated here as the average number of contacts $(c_{a,b})$ that a susceptible individual of age group *a* makes with a colonized individual from age group *b*, per unit time). The parameter $\alpha_{a,b}$ (0 or 1) controls transmissibility of carriage from age group *b* to age group *a* regardless of vaccination status and independent of serotype.

We first define the proportions of population in age group *b* who are colonized with one, two, or three STs.

$$\mathscr{p}_{b,i} = (\mathcal{C}_{u,b,i} + \mathcal{C}_{v,b,i} + \mathcal{C}_{w,b,i} + \mathcal{C}_{vw,b,i} + \mathcal{Q}_{v,b,i} + \mathcal{Q}_{w,b,i} + \mathcal{Q}_{vw,b,i})/\mathcal{P}_b$$

$$\mathscr{pp}_{b,i,j} = (\mathcal{CC}_{u,b,i,j} + \mathcal{CC}_{v,b,i,j} + \mathcal{CC}_{w,b,i,j} + \mathcal{CC}_{vw,b,i,j} + \mathcal{QQ}_{v,b,i,j} + \mathcal{QQ}_{w,b,i,j} + \mathcal{QQ}_{vw,b,i,j})/\mathcal{P}_b$$

$$\begin{aligned}
& \mathscr{ppp}_{b,i,j,k} \\
& = (\mathcal{CCC}_{u,b,i,j,k} + \mathcal{CCC}_{v,b,i,j,k} + \mathcal{CCC}_{w,b,i,j,k} + \mathcal{CCC}_{vw,b,i,j,k} + \mathcal{QQQ}_{v,b,i,j,k} + \mathcal{QQQ}_{w,b,i,j,k} + \mathcal{QQQ}_{vw,b,i,j,k})/\mathcal{P}_b
\end{aligned}$$

The force of acquisition of carriage of a single serotype *i*, $\lambda_{a,i}$, by an unvaccinated individual of age group *a*, is given as follows.

$$
\begin{aligned}
\lambda_{a,i} = \sum_{b=1}^{nA} \alpha_{a,b} c_{a,b} \Bigg( & \beta_{a,i} \wp_{b,i} + \sum_{j=i+1}^{nS} (\beta_{a,i} - \beta_{a,i,j}) \wp\wp_{b,i,j} + \sum_{j=1}^{i-1} (\beta_{a,i} - \beta_{a,j,i}) \wp\wp_{b,j,i} \\
& + \sum_{j=i+1}^{nS-1} \sum_{k=j+1}^{nS} (\beta_{a,i} - \beta_{a,i,j} - \beta_{a,i,k} - \beta_{a,i,j,k}) \wp\wp\wp_{b,i,j,k} \\
& + \sum_{j=1}^{i-1} \sum_{k=i+1}^{nS} (\beta_{a,i} - \beta_{a,j,i} - \beta_{a,i,k} - \beta_{a,j,i,k}) \wp\wp\wp_{b,j,i,k} \\
& + \sum_{j=1}^{i-2} \sum_{k=j+1}^{i-1} (\beta_{a,i} - \beta_{a,j,i} - \beta_{a,k,i} - \beta_{a,j,k,i}) \wp\wp\wp_{b,j,k,i} \Bigg),
\end{aligned}
$$

$$i = 1, \dots, nS.$$

Rate of simultaneous acquisition of two STs by the unvaccinated individuals is given by:

$$
\begin{aligned}
\lambda_{a,i,j} = \sum_{b=1}^{nA} \alpha_{a,b} c_{a,b} \Bigg( & \beta_{a,i,j} \wp\wp_{b,i,j} + \sum_{k=j+1}^{\mathrm{nS}} (\beta_{a,i,j} - \beta_{a,i,j,k}) \wp\wp\wp_{b,i,j,k} + \sum_{k=i+1}^{j-1} (\beta_{a,i,j} - \beta_{a,i,k,j}) \wp\wp\wp_{b,i,k,j} \\
& + \sum_{k=1}^{i-1} (\beta_{a,i,j} - \beta_{a,k,i,j}) \wp\wp\wp_{b,k,i,j} \Bigg)
\end{aligned}
$$

Similarly, the force of single colonization of a $\sigma$- vaccinated individual is given by:

$$\lambda v_{\sigma,a,i} = (1-\epsilon_{\sigma,a,i})\sum_{b=1}^{nA}\alpha_{a,b}c_{a,b}\Bigg(\beta_{a,i}\wp_{b,i} + \sum_{j=i+1}^{nS}\left(\beta_{a,i}-(1-\epsilon_{\sigma,a,j})\beta_{a,i,j}\right)\wp\wp_{b,i,j}$$

$$+\sum_{j=1}^{i-1}\left(\beta_{a,i}-(1-\epsilon_{\sigma,a,j})\beta_{a,j,i}\right)\wp\wp_{b,j,i}$$

$$+\sum_{j=i+1}^{nS-1}\sum_{k=j+1}^{nS}\big(\beta_{a,i}-(1-\epsilon_{\sigma,a,j})\beta_{a,i,j}-(1-\epsilon_{\sigma,a,k})\beta_{a,i,k}$$

$$-(1-\epsilon_{\sigma,a,j})(1-\epsilon_{\sigma,a,k})\beta_{a,i,j,k}\big)\wp\wp\wp_{b,i,j,k}$$

$$+\sum_{j=1}^{i-1}\sum_{k=i+1}^{nS}\big(\beta_{a,i}-(1-\epsilon_{\sigma,a,j})\beta_{a,j,i}-(1-\epsilon_{\sigma,a,k})\beta_{a,i,k}$$

$$-(1-\epsilon_{\sigma,a,j})(1-\epsilon_{\sigma,a,k})\beta_{a,j,i,k}\big)\wp\wp\wp_{b,j,i,k}$$

$$+\sum_{j=1}^{i-2}\sum_{k=j+1}^{i-1}\big(\beta_{a,i}-(1-\epsilon_{\sigma,a,j})\beta_{a,j,i}-(1-\epsilon_{\sigma,a,k})\beta_{a,k,i}$$

$$-(1-\epsilon_{\sigma,a,j})(1-\epsilon_{\sigma,a,k})\beta_{a,j,k,i}\big)\wp\wp\wp_{b,j,k,i}\Bigg),$$

$$i = 1,\dots,nS.$$

The rate of simultaneous transmission in the vaccinated individuals is given by:

$$\lambda v_{\sigma,a,i,j} = (1-\epsilon_{\sigma,a,i})(1-\epsilon_{\sigma,a,j})\sum_{b=1}^{nA}\alpha_{a,b}c_{a,b}\Bigg(\beta_{a,i,j}\wp\wp_{b,i,j}$$

$$+\sum_{k=j+1}^{\mathrm{nS}}\left(\beta_{a,i,j}-(1-\epsilon_{\sigma,a,k})\beta_{a,i,j,k}\right)\wp\wp\wp_{b,i,j,k}$$

$$+\sum_{k=i+1}^{j-1}\left(\beta_{a,i,j}-(1-\epsilon_{\sigma,a,k})\beta_{a,i,k,j}\right)\wp\wp\wp_{b,i,k,j}$$

$$+\sum_{k=1}^{i-1}\left(\beta_{a,i,j}-(1-\epsilon_{\sigma,a,k})\beta_{a,k,i,j}\right)\wp\wp\wp_{b,k,i,j}\Bigg)$$

$$i = 1,\dots,nS-1,\; j = i+1,\dots,nS,$$

Rate of transmission of ST *i* from the colonized to the susceptible individual is determined by the following:

(i) $\alpha_{a,b}$, the likelihood (0 or 1) of transfer of carriage from age group *b* to age group *a*. In a longitudinal household study by Althouse *et al*. [34] age-specific transmission probabilities were found to be negligible from adults to other adults and from adults to children. However, other literature reports transmission of pneumococcal carriage from adults to other adults [35, 36, 37]. Consequently, we assume transmission of carriage from children to children and adults, from adults to adults, and no transmission from adults to children.

(ii) $\beta_{a,i}$, the probability of acquisition of ST *i* by a susceptible individual in age group *a* if he/she makes a single contact with an *i*-colonized individual. Note that when a contact with a co-colonized individual carrying STs *i* and *j* is made, both serotypes can be acquired. If, however, only ST *i* is acquired in such contact, the possibility of simultaneous acquisition of ST *j* should be excluded. This is represented by the factor $(1 - \beta_{a,j})$. Assuming that acquisition of one STC is independent of acquisition of another, the probability of simultaneous acquisition of double carriage is given by $\beta_{a,i,j} = \beta_{a,i}\beta_{a,j}$, and probability of simultaneous acquisition of triple carriage is assumed to be zero ($\beta_{a,i,j,k} = 0$).

(iii) $c_{a,b}$, the average number of contacts a susceptible individual makes with age group *b,* per unit time.

(iv) $p_{b,i}$, $pp_{b,i,j}$ and $ppp_{b,i,j,k}$, the proportions of population in age group *b* colonized, respectively, by one, two or three STs.

(v) $\epsilon_{\sigma,a,i}$, the vaccine-efficacy against acquisition of ST *i*.

##### 3.2.5 Estimates of the epidemiologic model parameters

Data on carriage transmission is reported primarily in pediatric populations. If age appeared to be the determinant of input values (for example clearance rate), known trends were projected to estimate the values for certain age groups. The event of carriage acquisition is not directly observed and is only detected through active sampling [38]. Therefore, the parameters related to colonization such as carriage prevalence, the probability of carriage transfer per contact, competition parameters, and carriage clearance rates can generally be derived based only on sampling studies. In the modeling literature most of these parameters are estimated through model calibrations.

###### *3.2.5.1 Carriage parameters*

Transmission dynamics governed by the model ODEs are solely related to carriage transmission. Since most nasopharyngeal carriage is among children, most sampling studies are carried out in child populations. Furthermore, carriage studies require active sampling, making such studies a challenging task. Unlike pneumococcal disease data (which gets reported/recorded due to symptoms resulting in individuals seeking medical care), there is limited carriage data available from specific research groups, carried out in a small population over a short period of time.

###### *3.2.5.1.1* Carriage Clearance Rate $\gamma_{\sigma a i}$

Annual carriage clearance rates for a given serotype were calculated as the inverse of carriage duration. A literature search was performed to collect data on age and serotype specific carriage durations (Table S7). These manuscripts focused on study populations in the Gambia [39, 40], Kenya [41, 42], South Africa [43], Thailand [44], Myanmar [44], France [45], Greece [46], UK [47], Sweden [48], Denmark [49], Portugal [50], and Finland [50]. While most of these studies focused on infants, some included older children and mothers.

Table S7. Literature review to estimate carriage clearance rates by ST and age.

| Source | Location | Ages | Notes |
|---|---|---|---|
| Chaguza et al., 2021 [40] | Gambia | 0-1 years | Contributed to the 0-2 age group |
| Abdullahi et al., 2012 [41, 42] | Kenya | 0-5 years | 40% went to our 0-2 age group, 60% went to the 2-5 a |
| Dube et al., 2018 [43] | South Africa | 0-1 years | Contributed to the 0-2 age group |
| Turner et al., 2012 [44] | Thailand & Myanmar | 0-1 years and mothers | Infants contributed to the 0-2 age group and mothers to the 18-50 age group |
| Cauchemez et al., 2006 [45] | France | 3-6 years | 50% went to the 2-5 age group, 50% went to the 5-18 |
| Grivea et al., 2014 [46] | Greece | 2-5 years | Contributed to the 2-5 age group |
| Sleeman et al., 2006 [47] | UK | 0-2 years | Contributed to the 0-2 age group |

| | | | |
|---|---|---|---|
| Hogberg et al., 2007 [48] | Sweden | All ages | The under 1 age group went into our 0-2 age group, their 1-2 age group was split between our 0-2 and 2-5 age group, their 3-4 age group went into our age 2-5 age group, their 5-6 and 7-18 age group went into our 5-18 age group, their greater than 18 went into our 18-50 age group |
| Auranen et al., 2009 [49] | Denmark | 0-3 years | 66% went into the 0-2 age group and 33% went into the 2-5 age group |
| Pessoa et al., 2013 [50] | Portugal and Finland | 0-18 years | In Portugal (mean age 2, range 1-3) 66% of individuals went into the 0-2 age group and 33% went into the 2-5 age group. In Finland (mean age 4, range 1-7), 16.6% of individuals went into the 0-2 age group, 66% went into the 2-5 age group, and 16.6% went into the 5-18 age group |

For each age-serotype class combination we added the total carriage duration across studies and divided it by the total N across studies, normalized by age group. Below is a detailed description for serotype class 1 (serotypes 18C, 9V, 19F, 14, 23F, 4, and 6B) for individuals aged 0-2. For serotype classes, all serotypes total carriage duration within each group were summed and divided by the total N for that group.

Below is an example walkthrough for the carriage duration calculation for a single serotype. For serotype 18C [40], which solely focused on infants aged 0-1, documented 7 individuals carried this serotype for an average of 24 days. The total person-days carried for this study is (7x24)=168. Abdullahi *et al*. [41, 42], which studied children aged 0-5 documented 29 individuals carried 18C for an average of 18.3 days. The total person-days carried was calculated as (29x18.3x0.4)=212.28, with an N of (29x0.4)=11.6. The multiplier of 0.4 (or 2/5) represents the number of children from the study (aged 0-5), who fit the 0-2 age group. Sleeman *et al*. [47] had 14 individuals carry 18C, with a mean duration of 94.5 days. However, they defined the start of carriage as halfway between a negative and their first positive swab. They used two studies to collect these data, one with monthly sampling intervals and another with bi-weekly sampling intervals. To account for their methodological differences, we subtracted 10 days from the mean duration (midpoint between the halfway points of the sample periods). So the total duration from Sleeman *et al*. [47] for 18C was (14x(94.5-10))=1183 person-days. Finally, Pessoa *et al*. [50] examined children in day care centers in Portugal and Finland. The mean ages in each were 2 and 4, though the ranges were 1-3 (Portugal) and 1-7 (Finland). There were no occurrences of 18C in Portugal, however 11 children acquired 18C in Finland for a mean duration of 55 days. We estimated that 1/6 of the children were under 2 in this age group, so the total person-days carried was (11x55x1/6)=100.83, with an N of 11. The totals for carriage person-days were summed and divided by the total N, which then provided the mean carriage duration in days. These values were then inverted and multiplied by 365.25 to calculate the annual clearance rate. While Dube *et al*. [43], Turner *et al*. [44], Hogberg *et al*. [48], Auranen *et al*. [49], and Hill *et al*. [39] covered this age group, these studies did not have data for 18C. Meanwhile Cauchemez *et al*. [45] and Grivea *et al*. [46] did not have any individuals in this age group.

The resulting estimates for carriage clearance rate, $\gamma_{\sigma,a,i}$, are given in Table S8.

Table S8. Carriage clearance rates ($\boldsymbol{\gamma_{\sigma,a,i}}$) by STC and age group.

| STC | 0-2 | 2-5 | 5-18 | 18-50 | 50-65 | 65+ |
|---|---|---|---|---|---|---|
| 1 | 5.19993 | 11.9372 | 15.1879 | 14.4653 | 13.6145 | 13.6145 |
| 2 | 27.3067 | 38.1223 | 39.3453 | 55.7076 | 55.7076 | 55.7076 |
| 3 | 14.8476 | 16.9884 | 16.3789 | 8.02747 | 8.02747 | 8.02747 |
| 4 | 6.29913 | 14.9045 | 16.8781 | 17.0253 | 17.0253 | 17.0253 |
| 5 | 6.81437 | 10.0898 | 16.6023 | 14.1348 | 14.1348 | 14.1348 |
| 6 | 4.17131 | 5.95508 | 5.98541 | 8.47454 | 8.47454 | 8.47454 |
| 7 | 10.5161 | 14.5681 | 14.9021 | 22.024 | 22.0239 | 22.024 |
| 8 | 9.28252 | 13.7244 | 14.1435 | 19.0039 | 17.0089 | 17.0089 |
| 9 | 8.64709 | 11.4573 | 12.7064 | 19.4462 | 16.7931 | 16.7931 |
| 10 | 9.97376 | 14.0261 | 16.8916 | 23.0298 | 20.8943 | 20.8943 |
| 11 | 10.3588 | 11.8974 | 14.6864 | 11.703 | 11.703 | 11.703 |

###### *3.2.5.1.2* Probability of carriage acquisition per contact, $\boldsymbol{\beta_{a,i}}$

We calibrate for the probability of carriage acquisition per contact and provide a detailed description in Section 5.3.1.2.

###### *3.2.5.1.3* Competition Parameter $\boldsymbol{\theta_{i,j}}$

Co-carriage with multiple STs has been observed to occur. For example, of all enrollees, over 40% of children were co-colonized in an infants study in Gambia [40]. Similarly, 22% of children in Netherlands [51], 29.8% in Indonesia [52], 20.1% in Portugal [53] and 16.6% in the UK [54] were found co-colonized with multiple STs. Evidence of co-carriage with 3 or more STs in literature is worth highlighting. 24/1159 (2.1%) co-colonized 13-48 months old children in an England study [54], and 4/17 co-colonized 6-34 months old children in a Spain study [55] were found to carry 3 or more STs. Furthermore, multiple ST detection is hampered due to low sensitivity in detection of low-prevalence STs. Tiley *et al*. [54] found simultaneous co-carriage of 6 STs.

Current carriage with a certain serotype may reduce the risk of acquiring another [1, 5, 11, 16, 56]. Such competition plays an important role in serotype replacement. Selective pressure due to pneumococcal vaccines reduces the prevalence of vaccine type (VT) carriage, thereby providing a competitive advantage to the non-vaccine type (NVT) over the VT [1, 57, 58].

Competition parameter $\theta_{ij}$ was used to denote the risk reduction factor for carriage of serotype *j* due to current carriage of serotype *i*. Competition manifests via reduction in the rate at which new serotypes are acquired, or through accelerated recovery of co-colonized individuals [49]. If there is some level of competition between a current serotype *i* and an invading ST *j* (so that $\theta_{ij} < 1$) then a reduction in the levels of *i* resulting from vaccination will make the niche available for *j*, which consequently increases replacing *i*. The stronger the competition, the more intense replacement is to be expected from the invading ST.

Multiple modeling studies have published data on the extent of serotype competition between pairs of serotypes – VT-NVT or mixed  [4, 6, 11, 12, 19, 49, 56, 59, 60]. The geographically diverse data differs broadly in values. For example, while Bottomley *et al*. [12] estimates the competition parameter between any two serotypes (defined as the reduction in the rate of acquisition among carriers relative to non-carriers) using the pre-vaccination data from The Gambia in a dynamic transmission model to be 0.63 (i.e., carriage with one serotype reduces the likelihood of acquiring carriage with another serotype by 37%), Numminen *et al*. [60] report a competition between any two serotypes of 0.09 using carriage data among daycare children in Norway (observing a relatively similar group of serotypes as Bottomley *et al*. [12]).

Other researchers have considered serotype competition associated with VT versus NVT. For example, Melegaro *et al*. [4] report a competition parameter value of 0.85 between PCV7 VTs and NVTs (regardless of which group the residing serotype versus the invading serotype belongs), using longitudinal carriage data from England and Wales. On the other hand, Van Effelterre *et al*. [11] estimate a competition parameter value of 0.267 between any two PCV7 types, using US carriage data in children under 2 years of age.

Hoti *et al*. [61] estimate the competition parameter value of 0.68 between any two serotypes among a group of serotypes based on data from Finland, in a population of varying age, in daycare center and family settings. The list of serotypes considered here is comparable to that in Bottomley *et al*. [12] which reports a competition parameter value of a 0.63 based on the US data covering all ages.

Most modeling studies calibrate for a competition parameter. In a triple colonization event, we assume independence of competition encountered by an invading third ST with either of the two resident STs (i.e., $\theta_{i,j,k} = \theta_{i,k}\theta_{j,k}$) or through simultaneous acquisition of two STs in presence of one (i.e., $\theta\theta_{i,j,k} = \theta_{i,j}\theta_{i,k}$).

###### 3.2.5.2 *Disease parameters*

The model assessed the epidemiological analyses of different vaccination strategies with IPD, NBPP and AOM outcomes.

###### *3.2.5.2.1* Probability of pneumococcal disease from carriage

The model incorporates the disease proportion (proportion of individuals that develop a pneumococcal disease given carriage) as the fraction $\rho_{a,i}$ of colonized individuals in age group *a* who develop the disease with serotype *i.* The progression to disease occurs shortly after carriage acquisition [62, 63]. In literature, case-to-carrier ratio [64] (also known as invasiveness, invasive ratio or invasive capacity) of a specific serotype is generally reported as disease cases per 100,000 colonized individuals (or person-years) with that serotype. The estimates of $\rho_{a,i}$ for IPD obtained from calibration are reported in Section 5.3.1.3.

###### 3.2.5.3 *Vaccine parameters*

The model distinguishes between the efficacy of a vaccine against carriage acquisition and the efficacy against disease development. PPSV23 is known to protect against pneumococcal disease, but it does not prevent carriage acquisition. We assumed that all PCVs protected against carriage and disease. For a cohort of vaccinated and placebo, we define $p_A$ to be the probability of carriage acquisition, $p_D$ to be the probability of disease given acquisition, $\epsilon_v$ as the effectiveness against acquisition and $\epsilon_d$ to be the effectiveness against disease given acquisition. Then the probability of disease in the placebo arm and vaccine arm, is given, respectively, by $p_A \times p_D$ and $p_A(1-\epsilon_v) \times p_D(1-\epsilon_d)$. VEd is given by

$VEd = 1 - \frac{p_A(1-\epsilon_v)\times p_D(1-\epsilon_d)}{p_A \times p_D} = 1-(1-\epsilon_v)\times(1-\epsilon_d)$, so that, $\epsilon_d = \frac{VEd-\epsilon_v}{1-\epsilon_v}$.

Furthermore, vaccine efficacy is different from one serotype to another and varies for each disease.

The mean duration of protection for a PCV or PPSV23 is based on [65], which assumed no decline of protection from a PCV for the first 5 years, followed by a linear decline to 0 over the next 10 years. This leads to an average duration of 10 years of protection. Meanwhile, PPSV23 protection was assumed to linearly decline to 0 over 15 years, which yields an average duration of protection of 7.5 years at the population level.

VE in adults is further adjusted by the risk proportions in the population. The proportions of healthy, at-risk and high-risk adults (Table S9) are based on a retrospective study by Pelton *et al*. [33].

Table S9. Adult populations by risk groups (Pelton, 2019).

| | **Healthy** | **At-risk** | **High-Risk** |
|---|---|---|---|
| **18–49 years** | 87.30% | 10.40% | 2.30% |
| **50–64 years** | 69.20% | 23.40% | 7.40% |
| **65+** | 49.1% | 36.2% | 14.7% |

###### *3.2.5.3.1* Vaccine Efficacy against IPD

VE against IPD is listed in Table 1 in the main text. VEd of PCV13 against STC1-type IPD in children is assumed to be 96% (CI: 93-98), identical to PCV7 VEd published in literature [66]. Taking ST-specific VEds for the additional PCV13 types from Moore *et al*. [67] the VEd for each STC in children is aggregated as the average of constituent STs, weighted by the number of IPD cases recorded in the ABCs data in children under 5 during the years 2010-2014 (to match with the age group and study period of the Moore *et al*. [67] data).

VEd of PCV13 against non-ST3 IPD in the adult age groups is assumed to be 75% in healthy and low-risk population (95% CI: 41.4-90.8) (68) and 25% in the high-risk population (95% CI:13.8-30.3), assuming 1/3 of healthy/low-risk adults [68]. VEd against ST3 IPD in health and low-risk population is assumed to be 26% (95% CI: 0-53.4) [69] whereas in the high-risk population it is assumed to be 8.7% (95% CI: 0-17.8) [68]. VEd of PCV13 in the adult age groups is adjusted by applying the risk distribution (Table S9) in adults. VEs of PCV15 and PCV20 are assumed to be the aggregate VEd of PCV13.

VEd of PPSV23 against IPD is assumed to be 59.7% (95% CI: 47.4-69.1) in healthy and low-risk adults and 7.9% (95% CI: 0-34.2) in high-risk adults [65].

###### *3.2.5.3.2* Vaccine Efficacy against NBPP

The VEd of PCV13 against NBPP in children is assumed to be 87.2% [70]. The VEd in healthy adults is 66.7% and 40.3% in CMC [71]. These assumptions are aligned with the estimates used for the ACIP meeting of September 2021 [65]. For the IC individuals also, we assumed a VEd of 15% in line with [65] (1/3 of the VEd in healthy/CMC adults). Finally, VEd against ST3 NBBP is assumed to be 15.6% in the healthy/CMC individuals and 5.2% in the IC individuals based on [72], which calculates this estimate by applying the ratio of IPD VEd/Pneumonia VEd for all PCV13 types to the corresponding point estimate for ST3 IPD VEd. Using risk proportions of Table S9, the adjusted VEd of PCV13 against NBPP is given by Table S10. VEd of PCV15 and PCV20 is assumed identical to PCV13 for common serotypes (STC1-STC5) and additionally against STC6 for PCV15 and against STC6, STC8, STC9 for PCV20.

Table S10. Vaccine efficacies against NBPP by age and serotype class. (PCV20 was only considered for adult vaccination).

| | | Age (years) | | | |
|---|---|---|---|---|---|
| | **STC** | **0-17** | **18-49** | **50-64** | **65+** |
| **PCV13** | **1,2,4,5** | 0.872 | 0.628 | 0.567 | 0.495 |
| | **3** | 0.872 | 0.154 | 0.148 | 0.141 |
| | **6-11** | 0 | | | |
| **PCV15** | **1-5** | Identical to PCV13 VE | | | |
| | **6** | 0.872 | 0.628 | 0.567 | 0.495 |
| | **7-11** | 0 | | | |
| **PCV20** | **1-6** | Identical to PCV15 VE | | | |
| | **7** | 0 | | | |
| | **8, 9** | NA | 0.628 | 0.567 | 0.495 |
| | **10, 11** | 0 | | | |
| **PPSV23** | **1-4, 6-9** | 0 | 0.197 | 0.190 | 0.180 |
| | **5,10,11** | 0 | 0 | 0 | 0 |

##### *3.2.5.3.3* Vaccine Efficacy against AOM

Serotype-specific VEd of PCV7 against AOM is obtained from Eskola *et al*. [73] and is used as VEd of PCV13 against STC1. VEd of PCV13 against the additional STs is obtained from Pichichero *et al*. [74]. VEd against AOM with each STC is calculated from the average of constituent VEds in that STC and is listed in Table S11. VEd of PCV15 against AOM with the additional STs (STC6) was assumed to be that of PCV7.

Table S11. Vaccine efficacies against AOM by age and serotype class.

| STC | PCV13 | PCV15 |
|---|---|---|
| **1** | 0.57 | 0.57 |
| **2** | 0.86 | 0.86 |
| **3** | 0.15 | 0.15 |
| **4** | 0.885 | 0.885 |
| **5** | 1 | 1 |
| **6** | 0 | 0.57 |
| **7-11** | 0 | 0 |

Using these VEd values, we derived the effectiveness against disease given carriage acquisition by also taking into account effectiveness against carriage VEc.

###### *3.2.5.3.4* Vaccine coverage data in age group 0-1

For age group 0-1, the VCR data from 2002 to 2018 is taken from CDC's ChildVaxView website [75]. This data is based on the National Immunization Survey (NIS-Child) to record PCV coverage among children 19-35 months old. It entails a telephone survey of parents/guardians of children aged 19-35 months in the U.S., and the coverage estimates are based on the provider-reported vaccination history. Vaccine types are confirmed from the vaccination providers who are contacted with the parents'/guardians' permission.  PCV7 was included in the children's NIP from 2000-2010 and PCV13 after 2010 in the 0-1 age group. Coverage estimates are presented by birth year (birth cohort), given by Table S12.

Table S12. VCR in age group 0-1.

| | **Coverage** | | **Coverage** |
|---|---|---|---|
| **2002** | 35.0 | **2012** | 79.5 |
| **2003** | 58.5 | **2013** | 80.3 |
| **2004** | 62.6 | **2014** | 80.9 |
| **2005** | 71.1 | **2015** | 82.1 |
| **2006** | 65.2 | **2016** | 80.1 |
| **2007** | 73.7 | **2017** | 80.6 |
| **2008** | 77.9 | **2018** | 82.1 |
| **2009** | 78.1 | **2019** | 82.6 |
| **2010** | 81.1 | **2020** | 84.4 |
| **2011** | 81.4 | | |

###### *3.2.5.3.5* Vaccine coverage data in age groups 18-49 and 50-64

VCR in 18-64 adults at increased pneumococcal disease risk is taken from the MMWR report of the CDC [76]. Given that 12.7% of all adults in age group 18-49 and 30.8% of all adults in 50-64 group is risk population [33], VCR estimates in the 18-49 and 50-64 age groups are obtained as summarized in Table S13.

Table S13. VCR in age groups 18-49 and 50-64.

| | Coverage among 18-64 at increased risk | Coverage among 18-49 regardless of risk | Coverage among 50-64 regardless of risk |
|---|---|---|---|
| **2010** | 18.5 | 2.3 | 5.7 |
| **2011** | 20.1 | 2.6 | 6.2 |
| **2012** | 20 | 2.5 | 6.2 |
| **2013** | 21.2 | 2.7 | 6.5 |
| **2014** | 20.3 | 2.6 | 6.3 |
| **2015** | 23 | 2.9 | 7.1 |
| **2016** | 24 | 3 | 7.4 |
| **2017** | 24.5 | 3.1 | 7.5 |
| **2018** | 23.3 | 2.96 | 7.2 |

###### *3.2.5.3.6* Vaccine coverage data in age group 65+

Vaccine coverage in 65+ is based on CDC's data [77] where the percent population vaccinated is estimated – for each respective year – from the proportion of those 65 and above who submit Medicare claims for a pneumococcal vaccination. VCR in 65+ adults is summarized in Table S14.

Table S14. VCR for different adult vaccinations in the 65+population. VCR is based on annual Medicare claims.

| | PCV13 | PPSV23 | PPSV23+PCV13 |
|---|---|---|---|
| **2010** | 0 | 41 | 0 |
| **2011** | 0 | 42 | 0 |
| **2012** | 0 | 44 | 0 |
| **2013** | 1 | 45 | 0 |
| **2014** | 1 | 43 | 2 |
| **2015** | 10 | 30 | 14 |
| **2016** | 14 | 22 | 22 |
| **2017** | 17 | 18 | 26 |
| **2018** | 17 | 15 | 30 |
| **2019** | 17 | 14 | 32 |

### 4 MODEL CALIBRATION TO VACCINATION COVERAGE DATA AND PROJECTED COVERAGE

We first calibrated the model to the vaccination coverage data in isolation from carriage/disease dynamics to ensure that it captured the implementation of interventions most accurately over the calibration time-period. Doing so warranted the model calibration with the overall IPD incidence in the presence of serotype-specific and age-specific vaccines, along with the recommendation changes during the calibration period.

#### 4.1 History of ACIP recommendations of pneumococcal vaccines

An account of the times of vaccine recommendations in the US is provided in Figure S7. PPSV23 was approved for use in 65+ in the year 1983. In addition, it was approved for use in children 2 years and older at increased risk for disease [78] and in 5+ at increased risk for serious disease [78]. It was recommended by the ACIP for 65+ routine immunization.

In the year 2000, PCV7 was introduced in infant series at 2,4, and 6 months, with a booster dose at 12-15 months [78] and in 24-59 months old with immunocompromised (IC) and chronic medical conditions (CMC) [78]. In children 2 years to 59 months at increased risk for PD the recommendation was PCV7+PPSV23 sequence (2 months apart) [78].

PCV13 replaced PCV7 in 2010. IC/CMC conditions were clarified in 2012 [79], accompanied by recommendation to 19+ with some IC/CMC to take PCV13+PPSV23 [79] and 19+ other IC/CMC to take PCV13+PPSV23 with PPSV23 revaccination [79].

All adults 65+ were recommended to take PCV13+PPSV23 sequential vaccination in the year 2015. This recommendation was revised in 2018 to either PPSV23-only or PCV13+PPSV23 sequence in all 19+, based on the shared clinical decision-making (SCDM) [80].

#### 4.2 Implications of ACIP recommendations for the model

- Vaccination of 65+ age group with PPSV23 is assumed to start in 2000 (before 2000 we assume no vaccine to obtain an initial steady-state IPD incidence).
- 2-64 high-risk individuals were also recommended PPSV23 since the time of PPSV23 introduction, so it is likely that there was a small fraction of this age group taking PPSV23 in the 2000-2017 period. However, it is worth highlighting that the model does not consider vaccination of age groups 2-4 and 5-17, nor is there any vaccine coverage data available for these age groups. Note, however, that vaccinated

population in 2-4 and 5-17 age groups includes a fraction that was vaccinated in the 0-1 age group and has aged into these age groups.

- The NHIS data show that there was considerable VCR in 18-64 age group before 2012 when sequential recommendation was made for the 19+ risk group. Therefore, the model assumes nonzero VCR in the 18-64 age group starting 2000 to obtain model fit with the available 2010-2018 data.

Time-varying cohort vaccination rates and continuous vaccination rates were adjusted in such a way that vaccination coverage predicted by the model matched the available historic data on vaccination coverage rates in different age groups. For this purpose, we first constructed a simplified vaccination only model, given by (5), comprising compartments corresponding to vaccine states and excluding the epidemiology of pneumococcal carriage transmission and disease. The model is calibrated by iteratively adjusting vaccination rates so that the vaccination coverage of each vaccine in corresponding age groups matches the data.

#### 4.3 Vaccine-only model

We denote the unvaccinated and vaccinated individuals in age group *a* by $\mathcal{U}_a$ and $\mathcal{V}_{\sigma,a}$ respectively, with the subscript $\sigma = v, w, vw$ denoting vaccination status with a PCV, PPSV23, and sequential vaccine PCV+PPSV23, respectively. Let $\phi_{v,1}$ and $\phi c_{\sigma,a}$ denote cohort vaccination in age group 1 and *a* > 1, respectively and $\psi_{\sigma,a}$ denote continuous vaccination rate in age group *a*. Let $\psi u_{vw,a}$, $\psi v_{w,a}$, and $\psi w_{v,a}$ denote, respectively, sequential vaccination of the unvaccinated, PPSV23 vaccination of those with PCV vaccination history and PCV vaccination of those with PPSV23 history, all three resulting in the sequential vaccination status. The age-structured ODE system (5) gives age- and PCV/PPSV23-specific vaccination coverage dynamics in the population. Other parameters are as defined in the transmission model.

$$
\begin{aligned}
\mathcal{U}_a{}' &= \delta_{1,a}\Lambda(1-\phi_{v,1}) + (1-\delta_{1,a})(1-(\phi c_{v,a}+\phi c_{w,a}))m_{a-1}\mathcal{U}_{a-1} - (m_a+\psi_{v,a}+\psi_{w,a} \\
&\quad + \mu_a)\mathcal{U}_a, \\
\mathcal{V}_{v,a}{}' &= \delta_{1,a}\Lambda\phi_{v,1} + (1-\delta_{1,a})m_{a-1}\phi c_{v,a}(\mathcal{U}_{a-1}+\mathcal{Q}_{v,a-1}+\mathcal{Q}_{vw,a-1}+\mathcal{Q}_{w,a-1}) + (1 \\
&\quad - \phi cv_{w,a})\mathcal{V}_{v,a-1} + \psi_{v,a}(\mathcal{U}_a+\mathcal{Q}_{v,a}+\mathcal{Q}_{vw,a}+\mathcal{Q}_{w,a}) - (m_a+\psi v_{w,a}+\mu_a \\
&\quad + \omega_{v,a})\mathcal{V}_{v,a}, \qquad (7) \\
\mathcal{V}'_{w,a} &= (1-\delta_{1,a})m_{a-1}\phi c_{w,a}(\mathcal{U}_{a-1}+\mathcal{Q}_{v,a-1}+\mathcal{Q}_{vw,a-1}+\mathcal{Q}_{w,a-1}) + (1-\phi cw_{v,a})\mathcal{V}_{w,a-1} \\
&\quad + \psi_{w,a}(\mathcal{U}_a+\mathcal{Q}_{v,a}+\mathcal{Q}_{vw,a}+\mathcal{Q}_{w,a}) - (m_a+\psi w_{v,a}+\omega_{w,a}+\mu_a)\mathcal{V}_{w,a}, \\
\mathcal{V}_{vw,a}{}' &= \psi v_{w,a}\mathcal{V}_{v,a} + (1-\delta_{1,a})m_{a-1}(\phi cv_{w,a}\mathcal{V}_{v,a-1}+\phi cw_{v,a}\mathcal{V}_{w,a-1}+\mathcal{V}_{vw,a-1}) - (m_a+\mu_a \\
&\quad + \omega_{vw,a})\mathcal{V}_{vw,a} + \psi w_{v,a}\mathcal{V}_{w,a},
\end{aligned}
$$

(5)

$$\mathcal{Q}_{v,a}{}' = \omega_{v,a}\mathcal{V}_{v,a} - (m_a + \psi_{v,a} + \psi_{w,a} + \mu_a)\mathcal{Q}_{v,a} + (1-\delta_{1,a})m_{a-1}(1-\phi c_{v,a} - \phi c_{w,a})\mathcal{Q}_{v,a-1},$$

$$\mathcal{Q}_{w,a}{}' = \omega_{w,a}\mathcal{V}_{w,a} - (m_a + \psi_{v,a} + \psi_{w,a} + \mu_a)\mathcal{Q}_{w,a} + (1-\delta_{1,a})m_{a-1}(1-\phi c_{v,a} - \phi c_{w,a})\mathcal{Q}_{w,a-1},$$

$$\mathcal{Q}_{vw,a}{}' = \omega_{vw,a}\mathcal{V}_{vw,a} - (m_a + \psi_{v,a} + \psi_{w,a} + \mu_a)\mathcal{Q}_{vw,a} + (1-\delta_{1,a})m_{a-1}(1-\phi c_{v,a} - \phi c_{w,a})\mathcal{Q}_{vw,a-1}.$$

Vaccination coverage of the vaccine $\sigma$ in age group $a$ at a given time $t$ is given by

$$VCR(a,\sigma) = \frac{\mathcal{V}_{\sigma,a}}{\mathcal{P}_a(t)}.$$

We fitted the vaccine-only model to the VCR data that led to the estimation of vaccination rates. Figure S6 shows model fit to data.

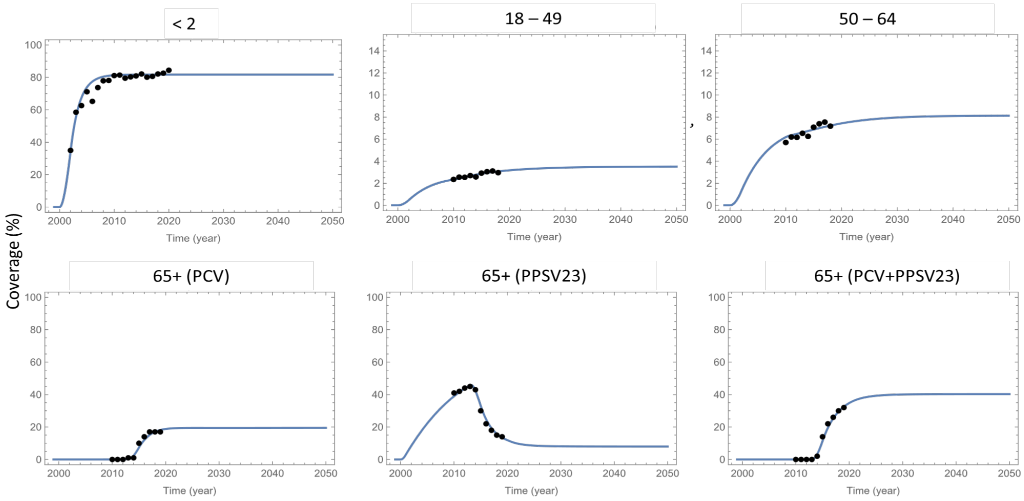

Figure S6. Fitted and projected vaccination coverage in respective age groups.

### 5 MODEL CALIBRATION TO IPD DATA

The model is calibrated to the historic data on IPD incidence in the US over the period 1998-2019 [81] by fitting model parameters which are not readily available from literature. The steady-state values are used as the initial condition for the epidemiological model. Calibration entails using the Nelder-Mead method to minimize a weighted sum of squared error's objective function over the annual IPD data and model outcome for each age group and each STC for each time-point. The Nelder-Mead method is implemented as one of the methods of the function NMinimize, a function to find a local minimum of a real-valued function, in Mathematica® 14.0 (https://reference.wolfram.com/language/ref/NMinimize.html).

#### 5.1 IPD incidence

Using the method of nonlinear least squares, we fit the incidence rate of IPD in each of the six age groups with each of the 11 serotype classes projected by the model to the incidence of IPD per 100,000 population. Mean age- and STC-specific IPD incidence from 1998-2000 was used to generate the steady-state IPD incidence in the year 2000 for each respective STC- and age-group combination. After the steady state was generated, the model was calibrated to the historical IPD data, which included multiple vaccine introductions and policy changes.

#### 5.2 Calibration procedure

Figure S7 shows timings of vaccine recommendation switches, with the overall IPD incidence (i.e., aggregated over all serotypes and across all age groups) overlaid on these timings (orange curve). We divide these timings into three periods for the sake of calibration. $T_0$ marks the pre-PCV period; $T_1$, the first PCV calibration period (2000-2010), marks the duration of PCV7 in pediatric immunization program (thus PPSV23 in 65+ and PCV7 in <2); $T_2$, the second calibration period (2010-2017), is the interval when PCV13 was in use in children under 2 replacing PCV7 in the year 2010. It overlaps the duration of recommendation of PCV13+PPSV23 in 19+ risk groups (in the year 2012), therefore sequential recommendation PCV13+PPSV23 in the 19+ individuals is assumed over the whole period $T_2$, for the purpose of calibration.

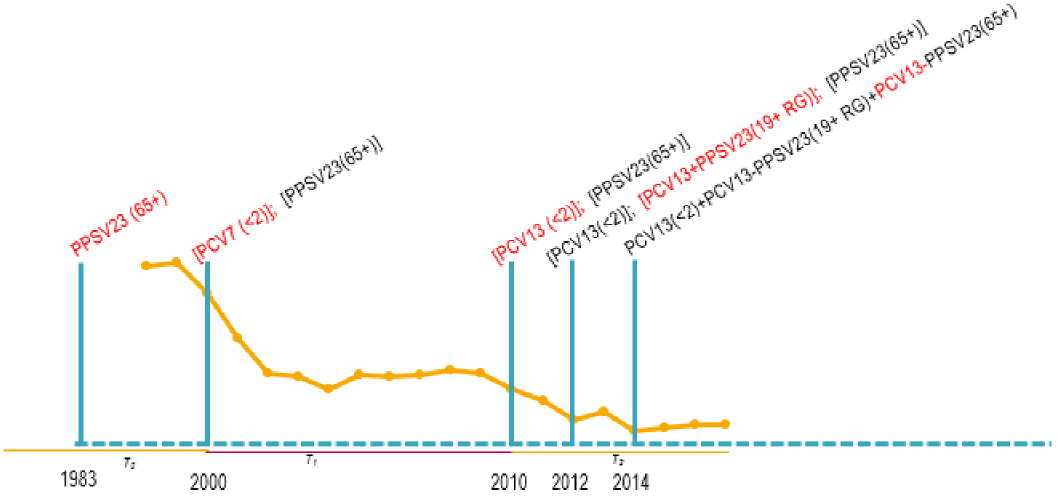

Figure S7. US pneumococcal vaccine recommendation timings and IPD incidence. $T_0$, $T_1$ and $T_2$ represent calibration periods. Red denotes new or changed recommendation. RG: Risk Group.

Calibration parameters include per contact carriage transmission probability ($\beta_{a,i}$), competition parameters ($\theta_{i,j}$), probability of developing pneumococcal disease after carriage (case-to-carrier ratio, $r_{a,i}$), and vaccine efficacy against acquisition ($\epsilon_{\sigma,a,i}$).

The first challenge in calibrating the model was to obtain the coexistence of the 11 different STCs at the no-vaccine steady-state. In other words, in a fully endemic steady state, all the carriage compartments in the model must be populated, which then yields positive IPD outcomes for all the serotype classes. This co-existence equilibrium is very delicate, and it is not reachable by a random search as it is often done with other models. For this model, initial manual adjustment guided by intuition and automated adjustment was needed to arrive an initial set of carriage parameters $\beta_{a,i}$.

Carriage prevalence refers to the prevalence of nasopharyngeal pneumococcal carriage in the individuals who are carriers of either single or multiple strains of the bacteria. The age-dependent and serotype specific prevalence of carriage is given by

$$
\begin{aligned}
CP(a,i) = \frac{1}{P_a(t)} \Bigg[ & \mathcal{C}_{u,a,i} + \sum_{\sigma \in \{v,w,vw\}} (\mathcal{C}_{\sigma,a,i} + \mathcal{Q}_{\sigma,a,i}) \\
& + \sum_{j=i+1}^{nS} \left( \mathcal{CC}_{u,a,i,j} + \sum_{\sigma \in \{v,w,vw\}} \left( \mathcal{CC}_{\sigma,a,i,j} + \mathcal{QQ}_{\sigma,b,i,j} \right) \right) \\
& + \sum_{j=1}^{i-1} \left( \mathcal{CC}_{u,a,j,i} + \sum_{\sigma \in \{v,w,vw\}} \left( \mathcal{CC}_{\sigma,a,j,i} + \mathcal{QQ}_{\sigma,b,j,i} \right) \right) \\
& + \sum_{j=i+1}^{nS-1} \sum_{k=j+1}^{nS} \left( \mathcal{CCC}_{u,a,i,j,k} + \sum_{\sigma \in \{v,w,vw\}} \left( \mathcal{CCC}_{\sigma,a,i,j,k} + \mathcal{QQQ}_{\sigma,b,i,j,k} \right) \right) \\
& + \sum_{j=1}^{i-1} \sum_{k=i+1}^{nS} \left( \mathcal{CCC}_{u,a,j,i,k} + \sum_{\sigma \in \{v,w,vw\}} \left( \mathcal{CCC}_{\sigma,a,j,i,k} + \mathcal{QQQ}_{\sigma,b,j,i,k} \right) \right) \\
& + \sum_{j=1}^{i-2} \sum_{k=j+1}^{i-1} \left( \mathcal{CCC}_{u,a,j,k,i} + \sum_{\sigma \in \{v,w,vw\}} \left( \mathcal{CCC}_{\sigma,a,j,k,i} + \mathcal{QQQ}_{\sigma,b,j,k,i} \right) \right) \Bigg].
\end{aligned}
$$

Carriage acquisition by unvaccinated individuals is given by

$$
\begin{aligned}
CA(u,a,i) = {} & \left(\lambda_{a,i} + \sum_{j=i+1}^{nS} \lambda_{a,i,j} + \sum_{j=1}^{i-1} \lambda_{a,j,i}\right)\left(\mathcal{N}_{u,a} + \sum_{\sigma\in\{v,w,vw\}} \mathcal{M}_{\sigma,a}\right) \\
& + \left(\sum_{\substack{j=1\\ j\neq i}}^{nS} \theta_{j,i}\lambda_{a,i} + \sum_{\substack{j=1\\ i<j}}^{nS} \theta_{j,i}\lambda_{a,i,j} + \sum_{\substack{j=1\\ i>j}}^{nS} \theta_{j,i}\lambda_{a,j,i}\right)\left(\mathcal{C}_{u,a,j} + \sum_{\sigma\in\{v,w,vw\}} \mathcal{Q}_{\sigma,a,j}\right) \\
& + \sum_{\substack{j=1\\ i\neq j}}^{nS} \sum_{\substack{k=i+1\\ j\neq k}}^{nS} \theta\theta_{j,i,k}\lambda_{a,i,k}\left(\mathcal{C}_{u,a,j} + \sum_{\sigma\in\{v,w,vw\}} \mathcal{Q}_{\sigma,a,j}\right) \\
& + \sum_{\substack{j=1\\ i\neq j}}^{nS} \sum_{\substack{k=1\\ j\neq k, k<i}}^{i-1} \theta\theta_{j,k,i}\lambda_{a,k,i}\left(\mathcal{C}_{u,a,j} + \sum_{\sigma\in\{v,w,vw\}} \mathcal{Q}_{\sigma,a,j}\right) \\
& + \Bigg(\sum_{\substack{j=1\\ i\neq j}}^{nS} \sum_{\substack{k=1\\ i\neq k, j<k}}^{nS} \theta_{j,k,i}\lambda_{a,i} + \sum_{\substack{j=1\\ j<i}}^{nS} \sum_{\substack{k=1\\ k\neq i, j<k}}^{nS} \theta_{j,k,i}\lambda_{a,j,i} + \sum_{\substack{j=1\\ i<j}}^{nS} \sum_{\substack{k=1\\ k\neq i, j<k}}^{nS} \theta_{j,k,i}\lambda_{a,i,j} \\
& + \sum_{\substack{j=1\\ j\neq i}}^{nS} \sum_{\substack{k=1\\ j<k<i}}^{nS} \theta_{j,k,i}\lambda_{a,k,i} + \sum_{\substack{j=1\\ j\neq i}}^{nS} \sum_{\substack{k=1\\ i<k, j<k}}^{nS} \theta_{j,k,i}\lambda_{a,i,k}\Bigg)\left(\mathcal{CC}_{u,a,j,k} + \sum_{\sigma\in\{v,w,vw\}} \mathcal{QQ}_{\sigma,a,j,k}\right),
\end{aligned}
$$

and by vaccinated individuals is given by (for $\sigma = v, w, vw$)

$$
\begin{aligned}
CA(\sigma,a,i) = {} & \left(\lambda v_{\sigma,a,i} + \sum_{j=i+1}^{nS} \lambda v_{\sigma,a,i,j} + \sum_{j=1}^{i-1} \lambda v_{\sigma,a,j,i}\right)\mathcal{N}_{\sigma,a} \\
& + \left(\sum_{\substack{j=1\\ j\neq i}}^{nS} \theta_{j,i}\lambda v_{\sigma,a,i} + \sum_{\substack{j=1\\ i<j}}^{nS} \theta_{j,i}\lambda v_{\sigma,a,i,j} + \sum_{\substack{j=1\\ i>j}}^{nS} \theta_{j,i}\lambda v_{\sigma,a,j,i}\right)\mathcal{C}_{\sigma,a,j} \\
& + \sum_{\substack{j=1\\ i\neq j}}^{nS} \sum_{\substack{k=i+1\\ j\neq k}}^{nS} \theta\theta_{j,i,k}\lambda v_{\sigma,a,i,k}\mathcal{C}_{\sigma,a,j} + \sum_{\substack{j=1\\ i\neq j}}^{nS} \sum_{\substack{k=1\\ j\neq k, k<i}}^{i-1} \theta\theta_{j,k,i}\lambda v_{\sigma,a,k,i}\mathcal{C}_{\sigma,a,j} \\
& + \left(\sum_{\substack{j=1\\ i\neq j}}^{nS} \sum_{\substack{k=1\\ i\neq k, j<k}}^{nS} \theta_{j,k,i}\lambda v_{\sigma,a,i} + \sum_{\substack{j=1\\ j<i}}^{nS} \sum_{\substack{k=1\\ k\neq i, j<k}}^{nS} \theta_{j,k,i}\lambda v_{\sigma,a,j,i} + \sum_{\substack{j=1\\ i<j}}^{nS} \sum_{\substack{k=1\\ k\neq i, j<k}}^{nS} \theta_{j,k,i}\lambda v_{\sigma,a,i,j}\right. \\
& \left. + \sum_{\substack{j=1\\ j\neq i}}^{nS} \sum_{\substack{k=1\\ j<k<i}}^{nS} \theta_{j,k,i}\lambda v_{\sigma,a,k,i} + \sum_{\substack{j=1\\ j\neq i}}^{nS} \sum_{\substack{k=1\\ i<k, j<k}}^{nS} \theta_{j,k,i}\lambda v_{\sigma,a,i,k}\right)\mathcal{CC}_{\sigma,a,j,k}, \qquad \sigma \in \{v, w, vw\}.
\end{aligned}
$$

Disease incidence per 100,000 with serotype $i$ at any time $t$ in unvaccinated individuals of age group $a$ is given by

$$D_{U,a,i}(t) = \rho_{a,i}\frac{CA(u,a,i)}{P_a(t)} \times 100{,}000,$$

while breakthrough disease incidence with serotype $i$ in individuals of age group $a$ who have been vaccinated with PCV, PPSV23 or sequential vaccination, is given by

$$D_{\sigma,a,i}(t) = \rho_{a,i}\left(1 - \epsilon_{D,\sigma,a,i}\right)\frac{CA(\sigma,a,i)}{P_a(t)} \times 100{,}000, \sigma \in v, vw,$$

$$D_{w,a,i}(t) = \rho_{a,i}\left(1 - \epsilon_{D,w,a,i}\right)\frac{CA(w,a,i)}{P_a(t)} \times 100{,}000.$$

To obtain carriage for all STC/AG combinations at steady state, target values about vaccine type distribution of the carriage and carriage prevalence by age group were sought. We set the target value for pre-vaccine carriage level in 0-1 age group to be about 40% and assumed carriage prevalence in other age groups, with the highest levels in extreme age-groups (Figure S8, left). Carriage distribution by vaccine types was assumed to roughly match the pre-PCV (2006/07) distribution in a UK study reported in Cleary *et al*. [82] (Figure S8, right). Figure S9 shows preliminary no-vaccine model fitting. Note that these target levels are achieved at the

2000 pre-vaccine steady state through the initial adjustment of the carriage parameters $\beta_{a,i}$ and $\theta_{i,j}$.

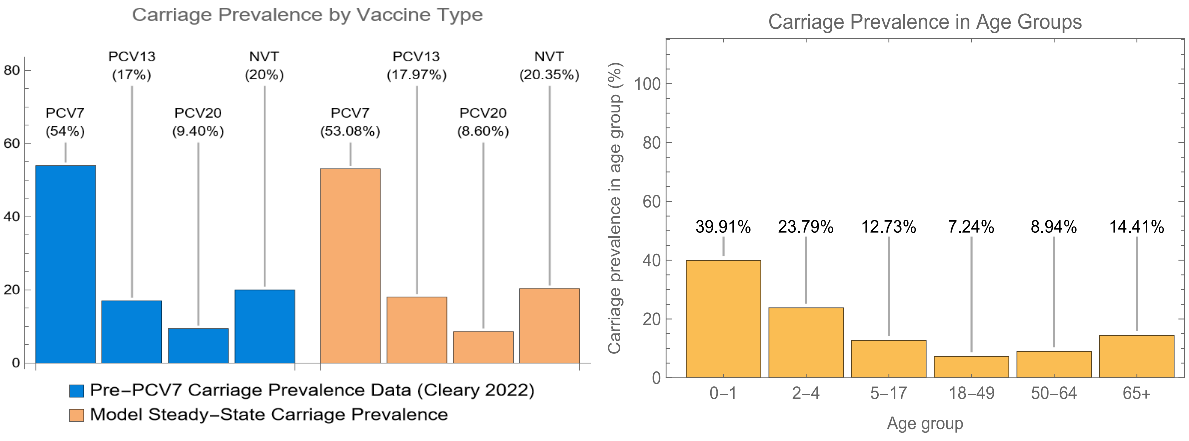

Figure S8. Carriage prevalence at steady state. Left: Carriage data from Cleary *et al*. [82] was used as a reference point for carriage values by vaccine type (blue). This was compared to the carriage levels in the model at pre-PCV steady state (orange). Right: Carriage levels assumed in model age groups at pre-PCV steady state.

Since IPD incidence data is available starting in 1998, the average of incidence in 1998, 1999 and 2000 in each age group is taken to represent the steady-state incidence in 2000. Once the suitable carriage levels by each STC in each group are obtained, one can adjust the invasiveness so that the model-predicted IPD incidence fits with the steady-state incidence data perfectly. That is due to the fact that IPD Incidence is computed as an outcome of the transmission model outside the ODE system, and this is where the invasiveness parameters $r_{a,l}$ appear.

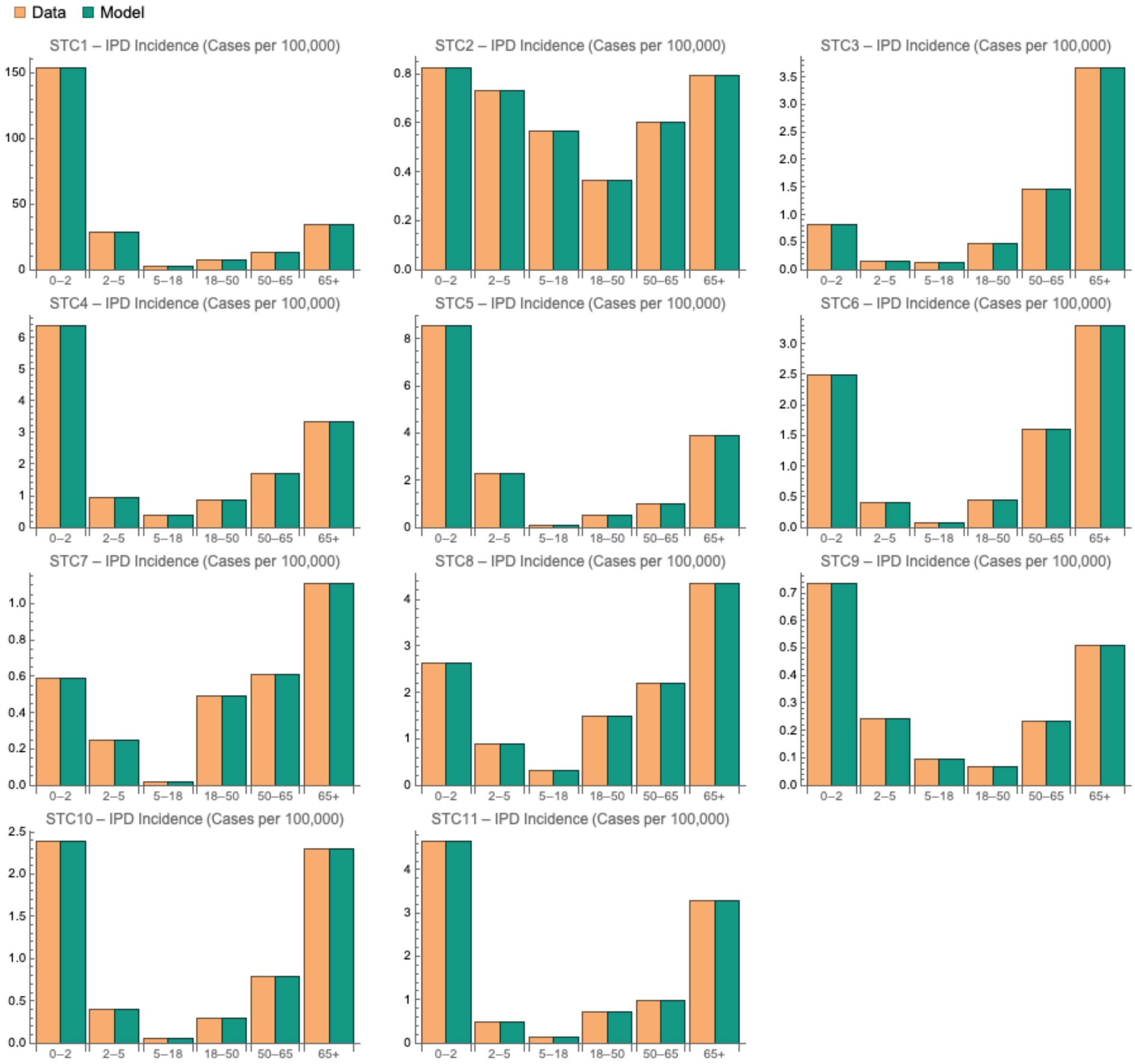

Figure S9. Initial fitting of the steady-state IPD incidence (per 100,000) by age group for each STC. Orange denotes the data and green denotes the model.

Figure S11 shows the model fit to IPD incidence data for each STC in each age group (AG). The model was calibrated to individual IPD incidence per 100,000 profiles for each STC/AG combination. Using the steady-state values (Figure S9) as the initial condition for the vaccine model, PCV7 was implemented in 2000 in the first model age group. This resulted in a steep decline of STC1 as illustrated by the green curve in the first plot of Figure S11 (a) (0-1 STC1), matching closely with the orange curve which represents the corresponding data. This decline of STC1 in the first model age group triggered similar decline in the later age groups through indirect effect and is shown in the rest of the first column plots in Figure S11 (a) and the first column plots of Figure S11 (c). Most other STCs – with the exception of STCs 4, 7 and 10 – were not impacted by PCV7 and remained at steady state. However, STCs 4, 7, and 10 showed replacement of STC1 (PCV7 types). Replacement of STC1 by STC 4, until PCV13 was implemented in 2010 causing a decline, is particularly noteworthy. This trend, that is clear in data (orange curve) was captured closely by the model (green curve).

Appropriate time-dependent parameters are defined to merge the periods $T_1$ and $T_2$ (Figure S7) in a unique ODE model solution. For example, the efficacy and waning rate, in the time period 2000-2010 have values that reflect PCV7 implementation in age group 0-1 and PPSV23 in the adult age groups. These parameters continuously change in 2010, with a transition time of 3 months for simplicity, to reflect the switch from PCV7 to PCV13. For the vaccine periods we use the coverage profiles for each relevant group based on the available data as explained before.

The model is calibrated by minimizing a weighted sum of squared errors objective function over the annual IPD data and model outcome for each age group and each serotype class at each time-point.

In the calibration period $T_1$ (2000-2010) of Figure S7 the subscript *v* stands for PCV7 and *w* represents PPSV23. In the second calibration period $T_2$ (2010-2019), PCV13 replaces PCV7 (so that the subscript *v* now stands for PCV13) in the first age group (i.e., children under 2) as well as all age groups 4, 5 and 6 where a PCV is administered. This switch is reflected by continuously changing the efficacy against acquisition and IPD, respectively, $\epsilon_{v,a,i}$ and $\epsilon_{D,v,a,i}$ with the additional six corresponding serotype classes *i = 2-5* from zero to non-zero for *a* = 1,4,5,6. Furthermore, in 2012 a new recommendation, namely the sequential regimen PCV13-PPSV23, was introduced in individuals 19 and above with risk conditions. This is implemented by assuming nonzero values for continuous vaccination rates $\psi w_{v,a}$ and $\psi v_{w,a}$ for a = 4,5,6 over the period $T_2$. It is worth highlighting that after the formal implementation of the minimization of mean squared error, visual inspection was used to choose between different available model fits. For example, data shows a steep decline in STC2 (STs 1 and 5) at the start of the calibration period, across almost all age groups (Figure S11 (c)). However, these STs were not in PCV7 and therefore it was hard to capture the initial decline through model dynamics. Therefore, model fits to STC2 data (all of which were more-or-less flat over the 1998-2010 period as expected) which tried to match the higher initial levels of STC2, were discarded through visual inspection, despite that some of those fits produced lower overall fit error.

#### 5.3 Calibration results

##### 5.3.1 Calibrated parameters

Model calibration led to the estimation of multiple parameters.

###### *5.3.1.1 Serotype competition $\theta_{i,j}$*

The competition parameters between STCs found clear relationships between specific serotype classes and how they behaved following vaccine introductions, which led to replacement. Following the introduction of PCV7, IPD incidence with some STCs – most noticeably STC4, STC7 and STC10 – increased as they faced less competition with PCV7 serotypes (STC1). In the pre-PCV7 period, STs in these STCs could not establish themselves at a steady state in presence of STC1. This is reflected in the low-valued competition parameters estimated between STC1 and STC4, STC1 and STC7, and STC1 and STC10 (Table S15). In addition, competition between STC1 and SCT2 and that between STC1 and STC3 were less than but closer to one, indicative of the presence of a weaker competition. As STC1 declined in response to PCV7, these STCs started to increase. Data shows an increase in both ST 7F and ST 19A (which comprise STC4) until 2009, when PCV13 was implemented that targeted these STs, which caused STC4 to decline. Likewise, competition parameters between PCV13-unique STCs and STC7 and STC10 are estimated to be less than one, which means that the PCV13-unique STCs (STC2, STC4 and STC5) also offer competition against STC7 (STs 9N, 17F, 20) and STC10 (the V116-unique STs). Therefore, this decline in response to PCV13 introduction results in ST replacement by STC7 and STC10.

In comparison, serotype competition estimates are reported as 63% [12] and 74% [4] using model structures different from the current model.

Table S15. Serotype competition. Likelihood of acquisition of 2nd ST (j, represented by columns) if currently colonized with one (i, represented by rows).

| | **1** | **2** | **3** | **4** | **5** | **6** | **7** | **8** | **9** | **10** | **11** |
|---|---|---|---|---|---|---|---|---|---|---|---|
| **1** | 0 | 0.9899 | 0.9901 | 0.025 | 1 | 1 | 0.8033 | 1 | 1 | 0.6981 | 1 |
| **2** | 1 | 0 | 1 | 1 | 1 | 1 | 0.8996 | 1 | 1 | 0.9899 | 1 |
| **3** | 1 | 1 | 0 | 1 | 1 | 1 | 1 | 1 | 1 | 1 | 1 |
| **4** | 1 | 1 | 1 | 0 | 1 | 1 | 0.9003 | 1 | 1 | 0.9904 | 1 |
| **5** | 1 | 1 | 1 | 1 | 0 | 1 | 0.9002 | 1 | 1 | 0.9898 | 1 |
| **6** | 1 | 1 | 1 | 1 | 1 | 0 | 1 | 1 | 1 | 1 | 1 |
| **7** | 1 | 1 | 1 | 1 | 1 | 1 | 0 | 1 | 1 | 1 | 1 |
| **8** | 1 | 1 | 1 | 1 | 1 | 1 | 1 | 0 | 1 | 1 | 1 |
| **9** | 1 | 1 | 1 | 1 | 1 | 1 | 1 | 1 | 0 | 1 | 1 |
| **10** | 1 | 1 | 1 | 1 | 1 | 1 | 1 | 1 | 1 | 0 | 1 |
| **11** | 1 | 1 | 1 | 1 | 1 | 1 | 1 | 1 | 1 | 1 | 0 |

###### 5.3.1.2 *Probability of carriage acquisition per contact $\beta_{a,i}$*

The model estimated that age- and serotype-specific carriage acquisition rates were highest in the youngest and oldest age groups. The calibrated probability of carriage acquisition of STC *i* per contact in each model age group is given by Table S16.

Table S16. Probability of carriage transmission of ST i (represented by columns) per contact in age group a (represented by rows).

| | | STC | | | | | | | | | | |
|---|---|---|---|---|---|---|---|---|---|---|---|---|
| | | 1 | 2 | 3 | 4 | 5 | 6 | 7 | 8 | 9 | 10 | 11 |
| Age group | 1 | 0.01243 | 0.01554 | 0.02036 | 0.00672 | 0.00615 | 0.00203 | 0.00871 | 0.00495 | 0.00435 | 0.01175 | 0.00697 |
| | 2 | 0.00755 | 0.01708 | 0.0044 | 0.00739 | 0.00673 | 0.00337 | 0.01629 | 0.00684 | 0.00617 | 0.0145 | 0.00326 |
| | 3 | 0.00285 | 0.01163 | 0.00461 | 0.00506 | 0.00463 | 0.00176 | 0.00353 | 0.00415 | 0.00372 | 0.00359 | 0.00438 |
| | 4 | 0.00276 | 0.00358 | 0.00104 | 0.0017 | 0.00117 | 0.00062 | 0.00365 | 0.002 | 0.002 | 0.00241 | 0.00128 |
| | 5 | 0.0039 | 0.00454 | 0.00174 | 0.00201 | 0.00138 | 0.00132 | 0.00385 | 0.00212 | 0.00211 | 0.0038 | 0.0016 |
| | 6 | 0.01185 | 0.01452 | 0.00807 | 0.00669 | 0.00554 | 0.00419 | 0.01308 | 0.0077 | 0.00713 | 0.01649 | 0.00802 |

###### 5.3.1.3 *Probability of disease from carriage acquisition $\rho_{a,i}$*

Age- and serotype-specific carriage acquisition rates were also highest in the youngest and oldest age groups. The calibrated probability of IPD with ST *i* in each age group from carriage acquisition is given by Table S17.

Table S17. Probability of IPD with ST i (represented by columns) in age group a (represented by rows).

| | | STC | | | | | | | | | | |
|---|---|---|---|---|---|---|---|---|---|---|---|---|
| | | 1 | 2 | 3 | 4 | 5 | 6 | 7 | 8 | 9 | 10 | 11 |
| Age group | 1 | 0.00093 | $1.2 \times 10^{-5}$ | $6.6 \times 10^{-5}$ | 0.00043 | 0.00027 | 0.00146 | $1.3 \times 10^{-5}$ | 0.0003 | 0.00014 | $4.1 \times 10^{-5}$ | 0.00018 |
| | 2 | 0.00023 | $8 \times 10^{-6}$ | 0.00016 | $9.4 \times 10^{-5}$ | $6.6 \times 10^{-5}$ | 0.00023 | $2 \times 10^{-6}$ | $6.7 \times 10^{-5}$ | $3.2 \times 10^{-5}$ | $1.1 \times 10^{-5}$ | $7.9 \times 10^{-5}$ |
| | 3 | $8.6 \times 10^{-5}$ | $3 \times 10^{-6}$ | $1.3 \times 10^{-5}$ | 0.00001 | $6 \times 10^{-6}$ | $2.5 \times 10^{-5}$ | $4 \times 10^{-6}$ | $1.3 \times 10^{-5}$ | $5 \times 10^{-6}$ | $1.4 \times 10^{-5}$ | $5 \times 10^{-6}$ |
| | 4 | 0.00011 | $1.6 \times 10^{-5}$ | 0.00025 | 0.00013 | 0.00015 | 0.00065 | $2.4 \times 10^{-5}$ | 0.00028 | $1.9 \times 10^{-5}$ | $5.1 \times 10^{-5}$ | $7.8 \times 10^{-5}$ |
| | 5 | 0.00015 | $2.9 \times 10^{-5}$ | 0.00067 | 0.00038 | 0.00038 | 0.00116 | $7.5 \times 10^{-5}$ | 0.00083 | $7.5 \times 10^{-5}$ | 0.00015 | 0.00021 |
| | 6 | 0.00027 | $2.5 \times 10^{-5}$ | 0.00064 | 0.00047 | 0.00062 | 0.00146 | $6.1 \times 10^{-5}$ | 0.00064 | 0.0001 | 0.00015 | 0.00019 |

###### 5.3.1.4 *Vaccine efficacy against carriage acquisition* $\epsilon_{\sigma,a,i}$

Calibrated VE against carriage acquisition is given by Table S18.VE of PCV13 against acquisition of PCV7 types is assumed to be identical to that of PCV7. Estimates of vaccine efficacy of PCV7 against carriage acquisition in the literature from the calibrations of different dynamic transmission models is reported to be 0.599 (CI: 0.4-0.6) [11] and 0.756 (CI: 0.69-0.84) [4] by fitting to the US data, a range of 0.08 (CI: 0.05-0.1) to 0.43 (CI: 0.35-0.5) by fitting to six different countries [21], and 0.93 using the derivation method laid out by Andrews *et al*. [62] for the UK data [24].

Table S18. Vaccine efficacy against carriage acquisition.

| Vaccine | Children age groups | Adult age groups |
|---|---|---|
| PCV7 | 28.0% | 20.0% |
| PCV13 (1,5) | 5.8% | 2.8% |
| PCV13 ST3 | 0.02% | 0.01% |
| PCV13 (7F, 19A) | 16.0% | 9.0% |
| PCV13 (6A, 6C) | 3.0% | 1.5% |

##### 5.3.2 Results of model fitting

The model outcome of steady-state IPD incidence given by Figure S9 is perturbed when calibration is run over the vaccine period 2000-2019, because now the dynamics are impacted by the vaccine. Vaccine efficacy against carriage acquisition is adjusted to align model output with the declining STC1 following PCV7 implementation, and serotype-competition parameter $\theta_{i,j}$ is re-adjusted to account for the resulting serotype-replacement of STC1 by STCs like STC4. These adjustments impact $\beta_{a,i}$ and $\rho_{a,i}$, so that the perfect fit of the steady state is altered. Figure S10 shows the updated steady-state IPD incidence which corresponds now to the calibrated parameter values consistent with the vaccine period. Figures S11 and S12 show that the model recapitulates the levels of IPD incidence per 100,000 in each model STC for each age group, the initial declines after the introductions of PCV7 and PCV13 in the pediatric age groups, and the steady-state following these declines. Figure S13 shows the resulting fit to the overall IPD incidence data in the US.

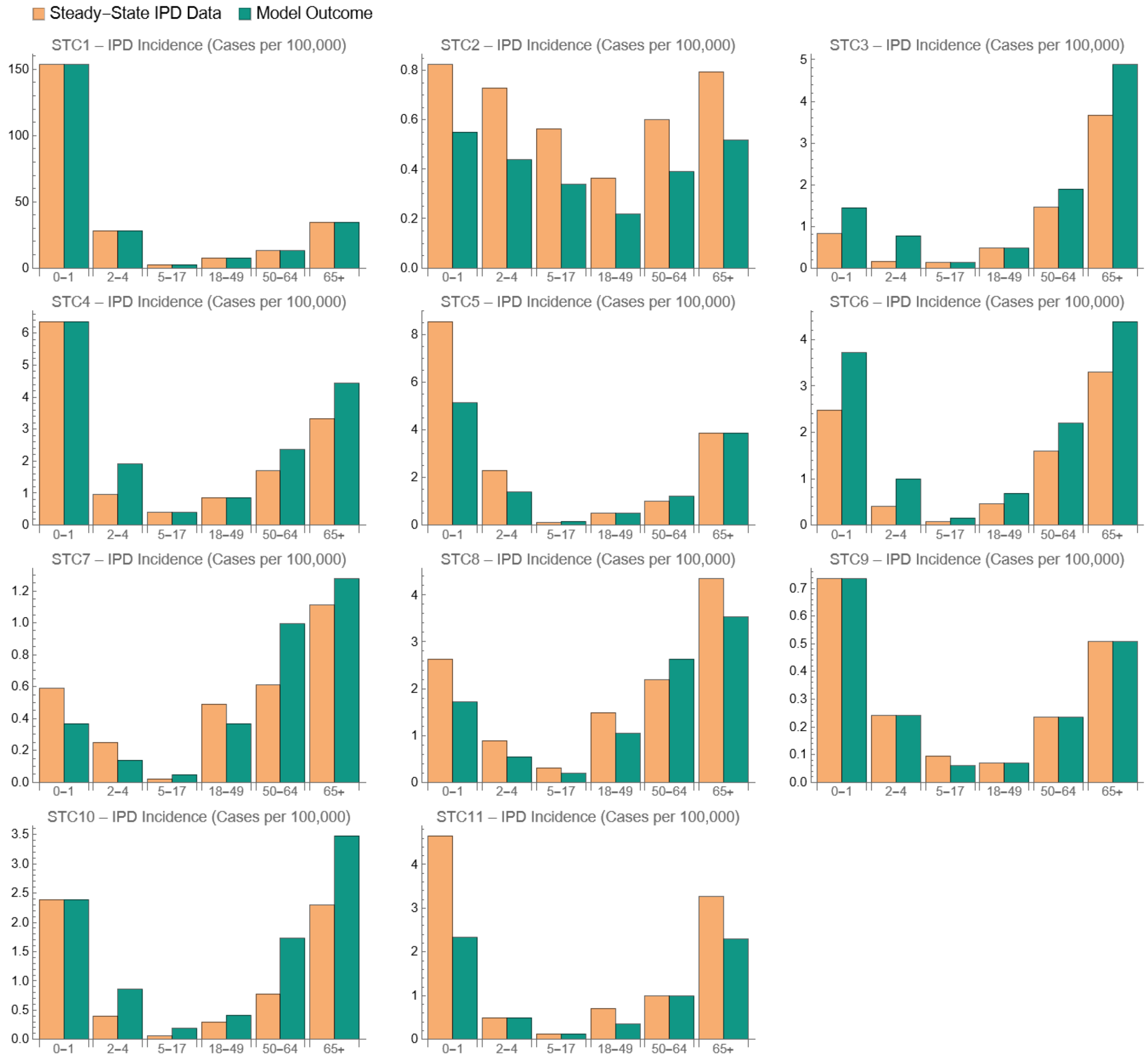

Figure S10. Model calibration versus IPD incidence (per 100,000) in 2000 by age group for each STC. Orange denotes the data and green denotes the model outcome.

(a): STC1-6, pediatric age groups.

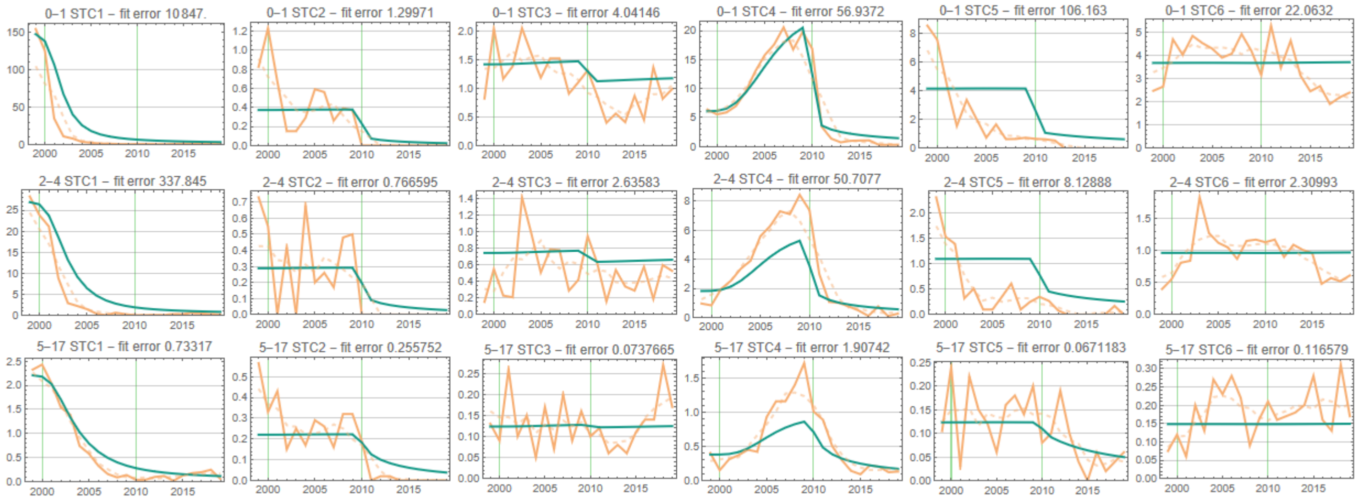

(b): STC7-11, pediatric age groups.

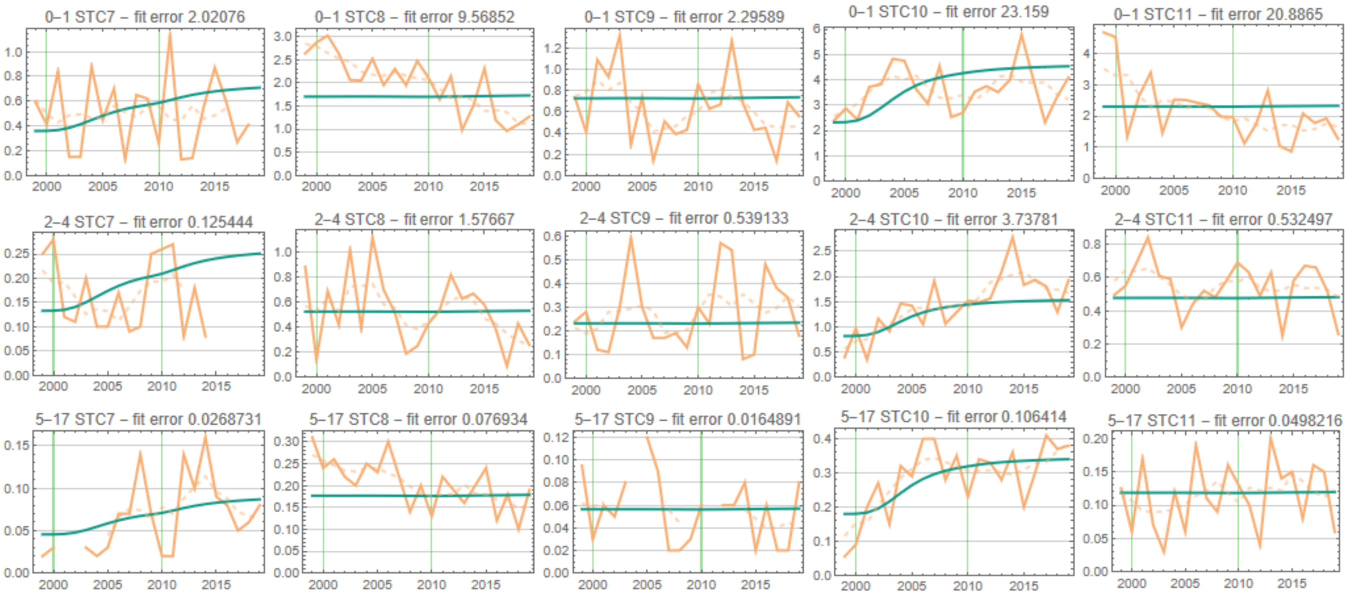

(c): STC1-6, adult age groups.

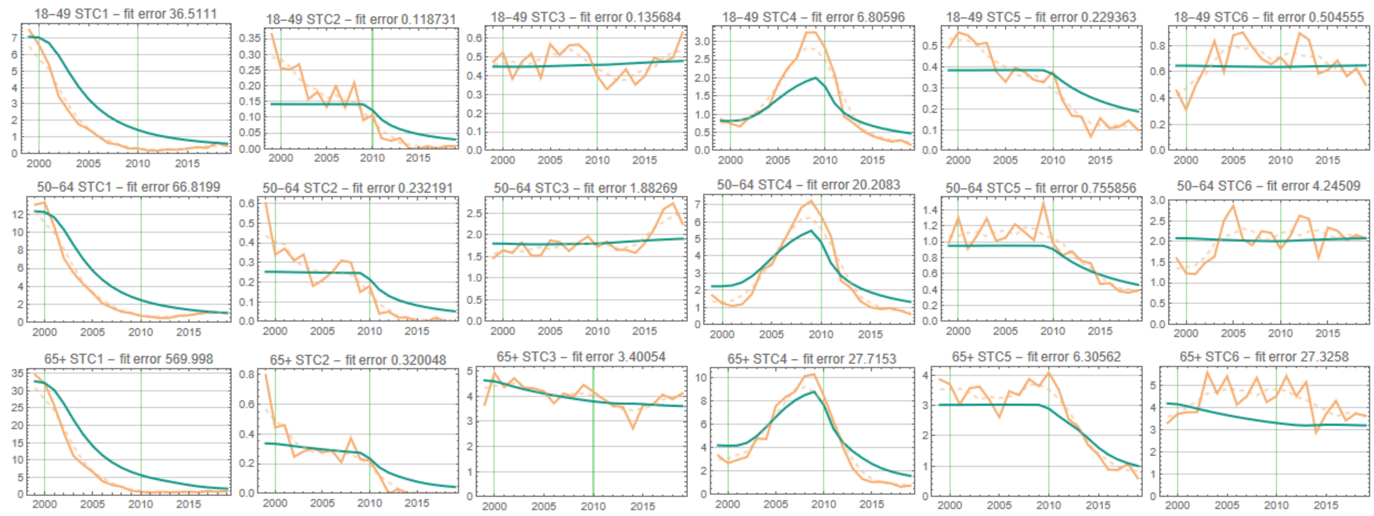

(d): STC7-11, adult age groups.

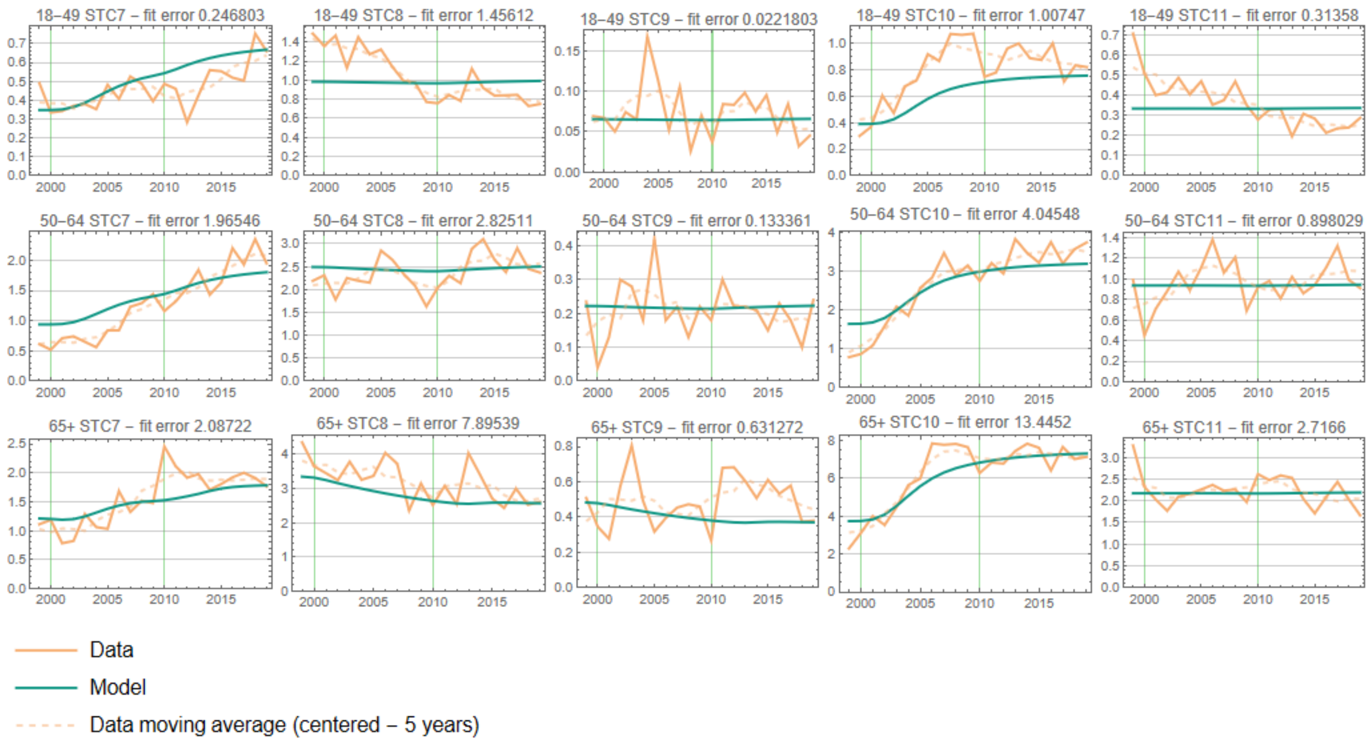

Figure S11. Model outputs versus IPD incidence (per 100,000) data by year for each STC and each age group. Vertical green lines denote the time of implementation of the PCVs (PCV7 in 2000 and PCV13 in 2010). Mean square difference for each plot was minimized. The 'fit error' shows this mean square difference at the fit.

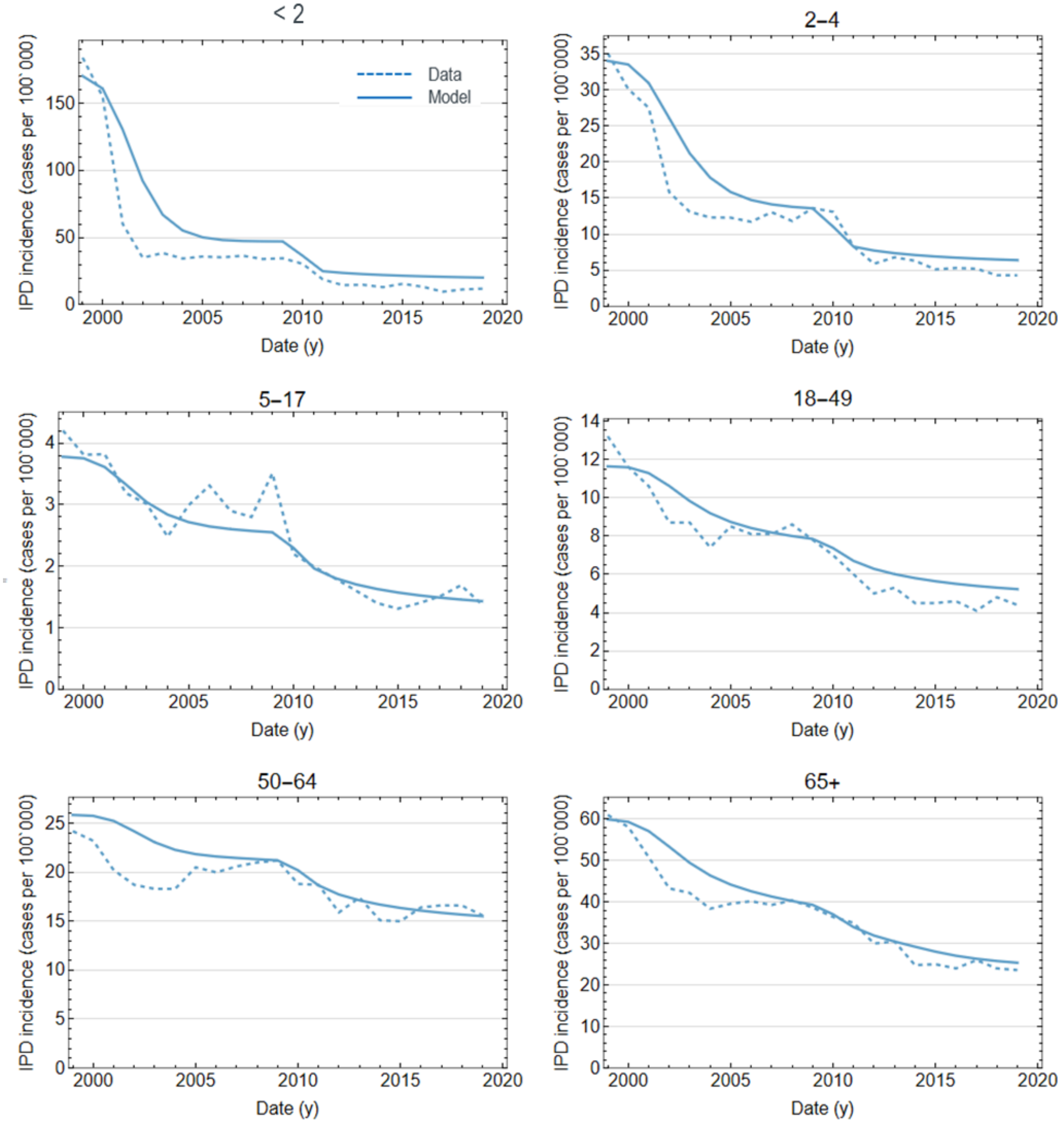

Figure S12. Model calibration versus IPD incidence (per 100,000) data by year for each age group. Dashed denotes the data and solid line denotes the model outcome.

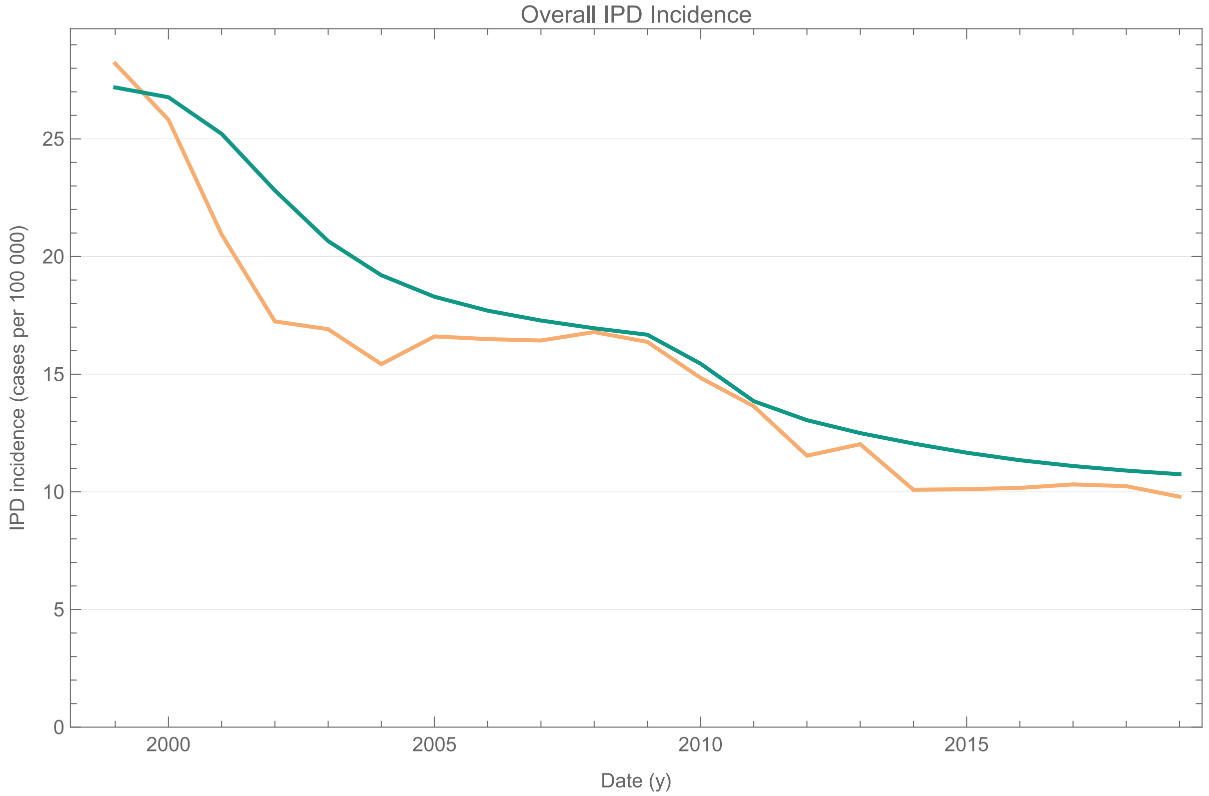

Figure S13. Model fitting of the overall IPD incidence (per 100,000) aggregated over age group and STC. Orange denotes the data and green denotes the model.

###### ***5.3.2.1 Model validation with carriage prevalence data***

Carriage prevalence data in the US are not reliably available for the broad population, therefore, they could not be used for model calibration. Much of the carriage prevalence data in the US are available for special populations, such as the Navajo Nation [83] and White Mountain Apache [84]. There are a few studies that capture nasopharyngeal carriage data for pediatric populations in the US that we have included as a source of external validation for our model (Figure S 14). Specifically, we compared the estimated percent of the pediatric population that is colonized over time to three key papers: Sharma et al. 2013 ( [85]), Lee et al. 2014 ( [86]), and Desai et al. 2015 ( [87]). The populations in these papers are from two different metropolitan areas (Atlanta, GA and Boston, MA) and there is variation across studies regarding the percentage of the pediatric population colonized. Carriage prevalence (the percentage of enrolled children who were colonized) estimated from these studies vary according to time, age, and geographical location.

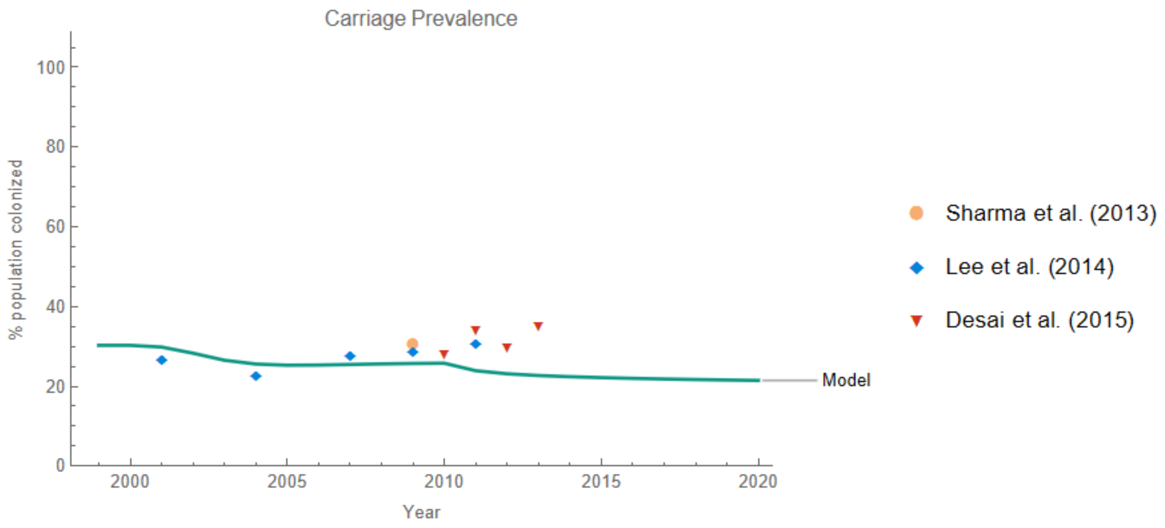

Figure S 14. Model validation: Comparison of carriage prevalence in the US children under 5 as generated by the model with data from Sharma et al. and Desai et al. for children under 5, and Lee et al. for children under 7.

### 6 MODEL CALIBRATION TO NBPP AND AOM DATA

Once the model has been calibrated with the IPD incidence data, we use the carriage solution to estimate the case-to-carrier ratios for the non-invasive diseases. That is, we now fix the calibrated parameters related to carriage – namely carriage acquisition probability ($\beta_{a,i}$), the competition parameter ($\theta_{i,j}$) and the vaccine efficacy against carriage acquisition ($\epsilon_{v,a,i}$) – and run another calibration to estimate the values of the CCR ($\rho_{a,i}$) for each of NBPP and AOM. To this end we use the historic respective data and fit the model outcome for the corresponding disease incidence.

#### 6.1 NBPP incidence

The overall NBPP incidence per 100k in the US children over the years 1998-2018 was obtained from Hu *et al*. [88] as shown in Table S19. Assuming serotype-distribution identical to IPD, the NBPP calibration target data was constructed.

To obtain the counterpart data in adults, we acquired ACP incidence for the years 2012-2020 (data were only available for this time period) from the Truven Health Analytics MarketScan Commercial Claims database. Adults with a pneumonia-related claim with International Classification of Diseases (ICD)-10 codes (J09.X1, J12-J18, J11.0, A22.1, A37.X1, B25.0, and B44.0) were identified (Table S19). We then assumed 11% of ACP had pneumococcal etiology—NBPP—in US adults [89].

To obtain the serotype distribution for NBPP in adults, we used two different papers [90, 91] reporting data from one multicenter surveillance study investigating radiographically confirmed CAP in 10 US cities between October 2013 and September 2016 and identifying the CAP with SP etiology and the serotypes associated with those NBPP cases for 18-64-year-olds and 65+ [92]. The key differences between the two papers were 1) the addition of a new, more sensitive assay (UAD2) designed to detect additional PCV20 serotypes (6A, 10A, 11A, 12F, 15B, 17F, 20, 22F, 8, and 9N), and 2) the 2019 paper [91] reported data on 39 serotypes while the 2021 paper only reported data on 24 serotypes (the serotypes reported in the first study but not the second include: 13, 15A, 15C, 16F, 21, 23A, 23B, 24F, 25F, 31, 34, 35B, 28, 6C, 7C, and 7C). Many of the reported serotypes in the 2019 paper but not the 2021 paper were those NVTs (mostly falling within STC10 and STC11) for which a sensitive assay does not exist. For those serotypes that were reported in both papers but changed in the number of NBPP presented from 2019 to 2021 (due to increased assay sensitivity), we estimated the multiplier between 2019 and 2021, for example, if there were 10 NBPP cases attributed to serotype 11A reported in 2019 and 40 NBPP cases attributed to serotype 11A in 2021, then the multiplier was 4. We calculated this multiplier for each overlapping serotype between the two papers, obtained the mean multiplier of 5.1, and then applied the mean multiplier to those serotypes reported in 2019 but not 2021, to mimic the increased NBPP cases that would be observed if those serotypes had been included in the more sensitive assay.

We used the serotype-specific NBPP data from the more sensitive UAD2 assay with the inferred serotype-specific NBPP data to calculate the percent of NBPP cases attributed to each STC for 18-64- and 65+ year-olds. We then applied this percentage to the overall NBPP data and obtained the STC- and age-specific calibration target.

Table S19. NBPP incidence (per 100,000). Pediatric data was obtained from Hu 2023. Adult data was extracted as 5% of the ACP obtained from the MarketScan database [77].

| **year** | **0-2** | **2-4** | **5-17** | **18-49** | **50-64** | **65+** |
|---|---|---|---|---|---|---|
| **1998** | 123.2777 | 73.89175 | 31.41537 | | | |
| **1999** | 137.3072 | 79.31542 | 30.85738 | | | |
| **2000** | 136.7242 | 77.09411 | 28.26775 | | | |
| **2001** | 110.5826 | 72.25302 | 34.78506 | | | |
| **2002** | 134.9783 | 95.49202 | 37.69829 | | | |
| **2003** | 105.1522 | 84.43184 | 34.92703 | | | |
| **2004** | 102.2389 | 78.80616 | 27.38574 | | | |
| **2005** | 111.5175 | 86.67696 | 33.56542 | | | |
| **2006** | 90.19012 | 73.05591 | 27.14055 | | | |
| **2007** | 84.55953 | 83.8845 | 31.75692 | | | |
| **2008** | 94.17509 | 71.33592 | 32.37657 | | | |
| **2009** | 94.48607 | 74.58573 | 34.91333 | | | |
| **2010** | 74.88648 | 66.50573 | 27.90969 | | | |
| **2011** | 67.91659 | 61.52647 | 28.92582 | | | |
| **2012** | 68.38477 | 61.87165 | 28.02154 | 83.7259 | 104.9155 | 288.4718 |
| **2013** | 53.2491 | 44.752 | 19.40757 | 109.9072 | 125.6285 | 295.8635 |
| **2014** | 40.84723 | 40.34029 | 16.10468 | 116.8404 | 131.693 | 293.0393 |
| **2015** | 43.52732 | 40.70034 | 18.11461 | 101.7649 | 125.2366 | 323.0925 |
| **2016** | 25.15567 | 22.11363 | 7.543041 | 45.05201 | 82.63699 | 274.6858 |
| **2017** | 18.108 | 17.73633 | 4.985948 | 40.93328 | 81.36261 | 291.3509 |
| **2018** | 14.16337 | 13.39164 | 4.653581 | 42.00799 | 81.74102 | 249.1763 |
| **2019** | | | | 44.61206 | 80.99799 | 242.452 |
| **2020** | | | | 43.72181 | 91.88969 | 221.7068 |

These data are used to calibrate the model for the NBPP-specific CCR ($\rho_{a,i}$). It is worth highlighting that the incidence data are available in different years in children and adult (with some overlap), and the all the available data in each age group were used to inform the CCR. This model fit is shown in Figure S14.

1252

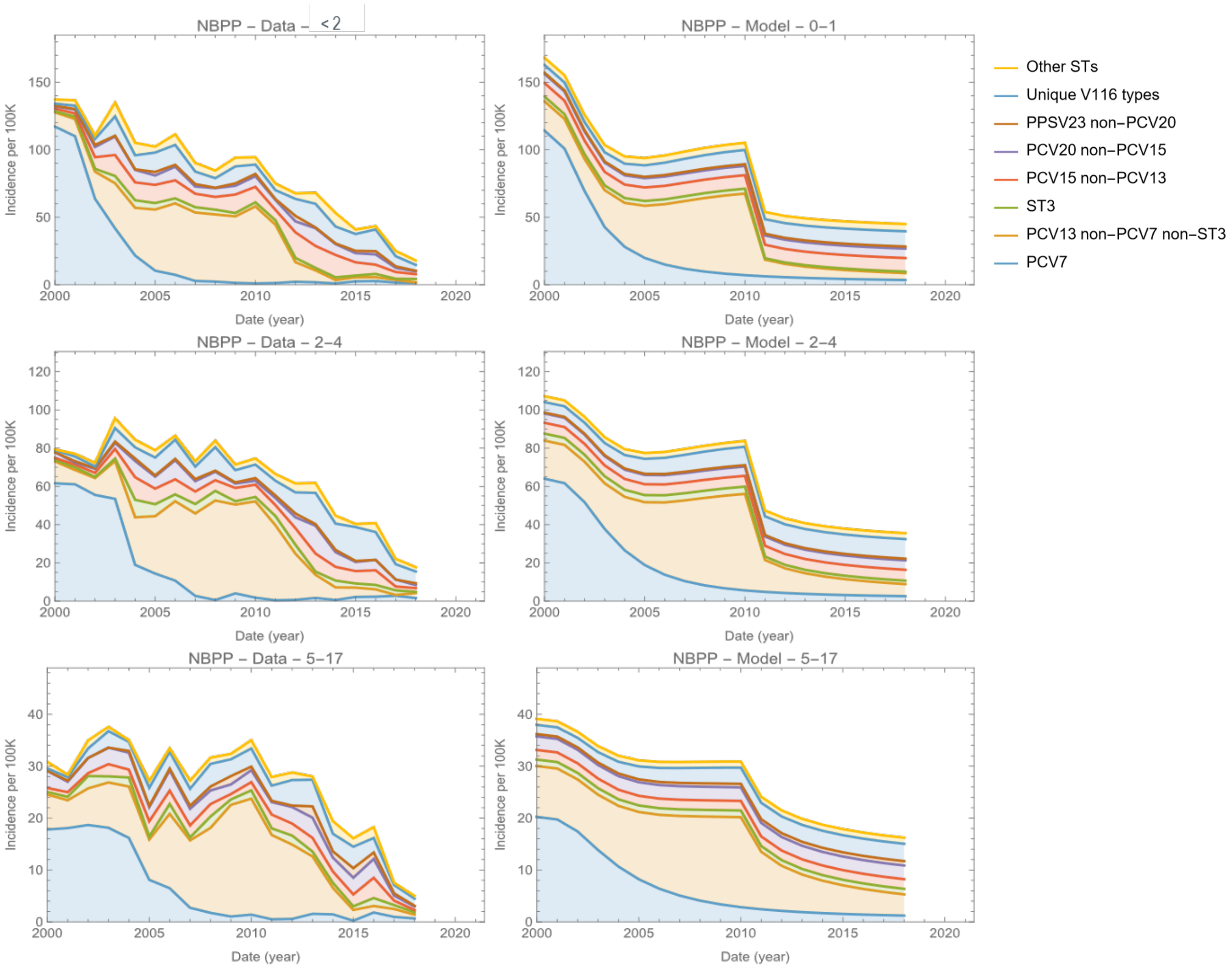

NBPP – Data – < 2
NBPP – Model – 0–1
NBPP – Data – 2–4
NBPP – Model – 2–4
NBPP – Data – 5–17
NBPP – Model – 5–17
Incidence per 100K
Date (year)
Other STs
Unique V116 types
PPSV23 non–PCV20
PCV20 non–PCV15
PCV15 non–PCV13
ST3
PCV13 non–PCV7 non–ST3
PCV7

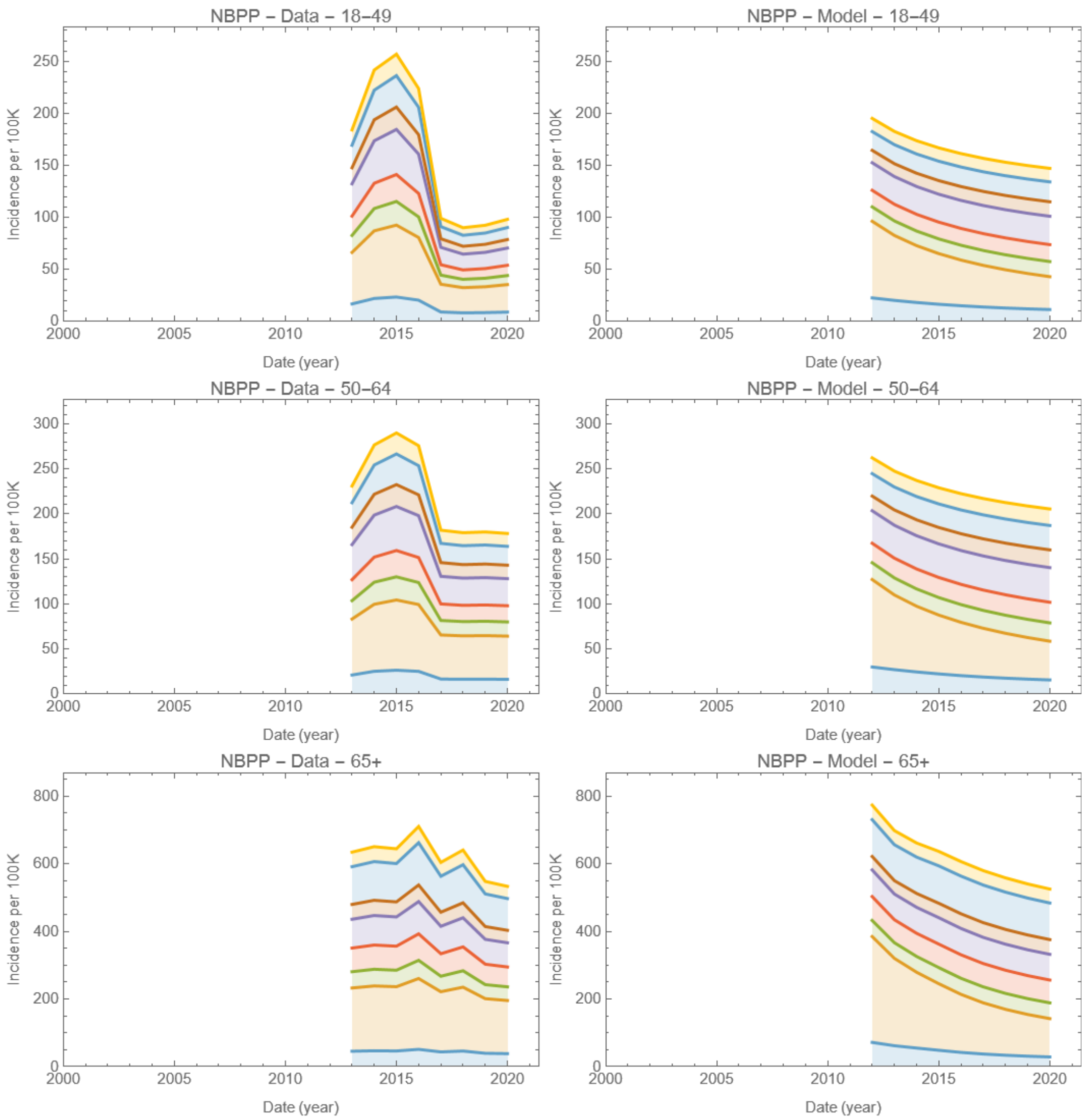

Figure S15. Model calibration to the NBPP incidence data by year for each age group. Pediatric data from 1998 was used for calibration, but adult data was only available beginning in 2012, which was used for calibration.

#### 6.2 AOM incidence

All-cause AOM incidence data over the years 1998-2018 was obtained from Hu *et al*. [93], of which 23.8% was pneumococcal attributable [94].

We split the 1998-2018 time series into PCV regimes, 1998-1999 as the pre-PCV regime, 2000-2009 as the PCV10 regime, and 2010-2018 as the PCV13 regime. We then identified studies with data on ST distribution available for AOM for the concomitant regimes: Block *et al*. [95] for the pre-PCV regime, Casey *et al*. [96] for the PCV10 regime, and Kaur *et al*. [94] for the PCV13 regime. We then calculated the percent of AOM cases attribute to each STC for pediatrics in the three different PCV regimes [94, 95, 96] and then applied these percentages to the overall AOM data and obtained the STC- and age-specific calibration target (Table S20).

Table S20. AOM incidence (per 100,000). Data was extracted from Hu et al. [88].

| | 0-1 | 2-4 | 5-17 |
|---|---|---|---|
| 1998 | 26446.34 | 12296.71 | 2663.833 |
| 1999 | 29223.58 | 13738.24 | 2834.016 |
| 2000 | 27427.32 | 12235.46 | 2525.796 |
| 2001 | 24590.48 | 11427.44 | 2493.741 |
| 2002 | 22864.25 | 11022.24 | 2282.915 |
| 2003 | 21178.5 | 10979.68 | 2355.586 |
| 2004 | 19596.61 | 9255.057 | 1915.185 |
| 2005 | 20379.05 | 9819.31 | 2146.442 |
| 2006 | 20137.71 | 10331.42 | 2117.454 |
| 2007 | 20048.97 | 10455.23 | 2200.458 |
| 2008 | 20525.43 | 10216.07 | 2193.744 |
| 2009 | 20613.68 | 10231.72 | 2398.227 |
| 2010 | 19238.42 | 9963.417 | 2226.815 |
| 2011 | 19300.07 | 10515.75 | 2441.999 |
| 2012 | 19796.69 | 10349.16 | 2365.735 |
| 2013 | 18865.89 | 9746.098 | 2235.579 |
| 2014 | 17330.99 | 9319.264 | 2110.849 |
| 2015 | 19099.51 | 10355.53 | 2313.519 |
| 2016 | 19143.98 | 10051.58 | 2214.284 |
| 2017 | 19023.5 | 9688.737 | 2213.807 |
| 2018 | 17570.99 | 9703.604 | 2118.343 |

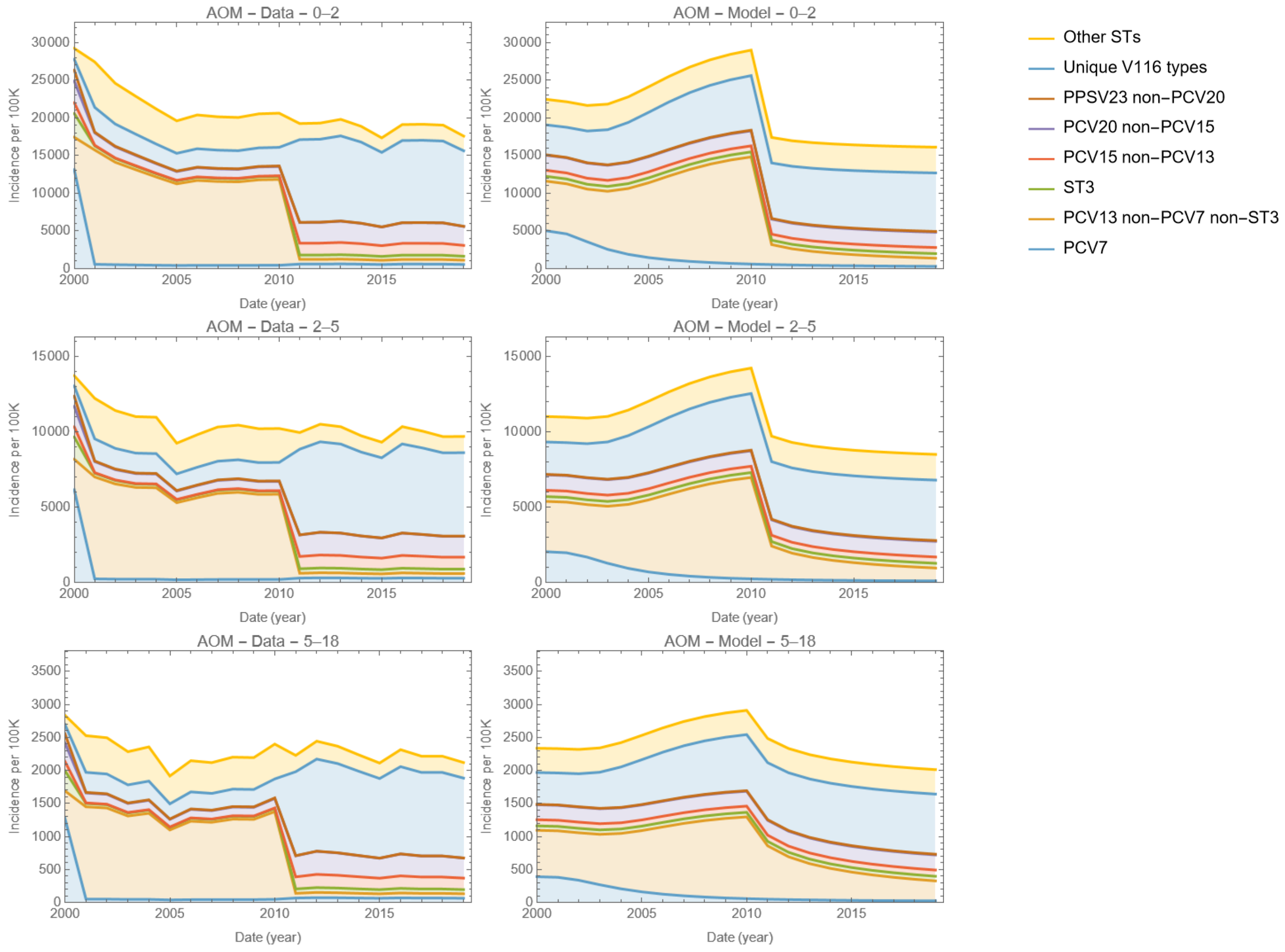

Figure S16. Model calibration to the AOM incidence data by year for each age group.

These data are used to calibrate the model for the AOM-specific CCR ($\rho_{a,i}$). This model fit is shown in Figure S15.

### 7 EPIDEMIOLOGIC PROJECTIONS

The calibrated model was used to project pneumococcal disease. These projections can be made at different levels of data aggregation, up to the 11 STCs and 6 age groups considered in the model. In the following projections we chose to aggregate STCs according to inclusion in vaccines relevant to the pediatric population.

#### 7.1 IPD projections

Projections of the overall IPD in all model age groups with the two scenarios in base case are presented in main text. Here, we present projections of IPD burden assuming VEd of PCV15 for each ST common with PCV13 is higher or lower than that of PCV13 if the immune response induced by PCV15 observed in clinical trials was respectively higher or lower for that ST.

###### 7.1.1 Scenario VE of PCV15 scaled by immunogenicity.

For this scenario VEd of PCV15 against IPD in children age group < 2 was based on Ryman *et al*. [97]. Left end point of the 95% CI was used as VEd of PCV15 against ST3 IPD. Overall VEd of PCV13 reported by Moore *et al*. [67] was used as VEd of PCV15 against STC6-type IPD in children. PCV13 VEd in adults was not ST-specific and was extended to additional PCV15 and PCV20 STCs (Table S21).

Table S21. Vaccine effectiveness against IPD by age and serotype class used for scenario analysis. (PCV20 was only considered for adult vaccination).

| | | **Age (years)** | | | |
|---|---|---|---|---|---|
| **PCV13** | **STC** | **0-17** | **18-49** | **50-64** | **65+** |
| | **1** | 0.96 | 0.739 | 0.713 | 0.677 |
| | **2** | 0.87 | 0.739 | 0.713 | 0.677 |
| | **3** | 0.303* | 0.256 | 0.247 | 0.235 |
| | **4** | 0.883 | 0.739 | 0.713 | 0.677 |
| | **5** | 0.86 | 0.739 | 0.713 | 0.677 |
| | **6-11** | 0 | | | |
| **PCV15** | **STC** | **0-17** | **18-49** | **50-64** | **65+** |
| | **1** | 0.96 | 0.739 | 0.713 | 0.677 |
| | **2** | 0.76 | 0.739 | 0.713 | 0.677 |
| | **3** | 0.67* | 0.256 | 0.247 | 0.235 |
| | **4** | 0.817 | 0.739 | 0.713 | 0.677 |
| | **5** | 0.59 | 0.739 | 0.713 | 0.677 |
| | **6** | 0.86 | 0.739 | 0.713 | 0.677 |
| | **7-11** | 0 | | | |
| **PCV20** | **1-6** | NA | Identical to PCV15 VE | | |
| | **7** | | 0 | | |
| | **8, 9** | | 0.739 | 0.713 | 0.677 |
| | **10, 11** | | 0 | | |
| **PPSV 23** | **1-4, 6-9** | NA | 0.585 | 0.559 | 0.521 |
| | **5,10,11** | | 0 | | |

IPD: Invasive Pneumococcal Disease; STC: Serotype Class; NA: Not Applicable;
PCV13: 13-valent pneumococcal conjugate vaccine; PCV15: 15-valent pneumococcal conjugate vaccine; PCV20: 20-valent pneumococcal conjugate vaccine; PPSV23: 23-valent pneumococcal polysaccharide vaccine; ST3 = serotype 3; STCs = serotype classes.

*: left end point of 95%CI.

Figure S17 shows model-projected IPD incidence per 100,000 in the US population with PCV15 (solid green line) and PCV13 (dashed orange line) in children using immunogenicity-based VEs from Table S21. The black line shows IPD incidence generated by the model over the calibration period.

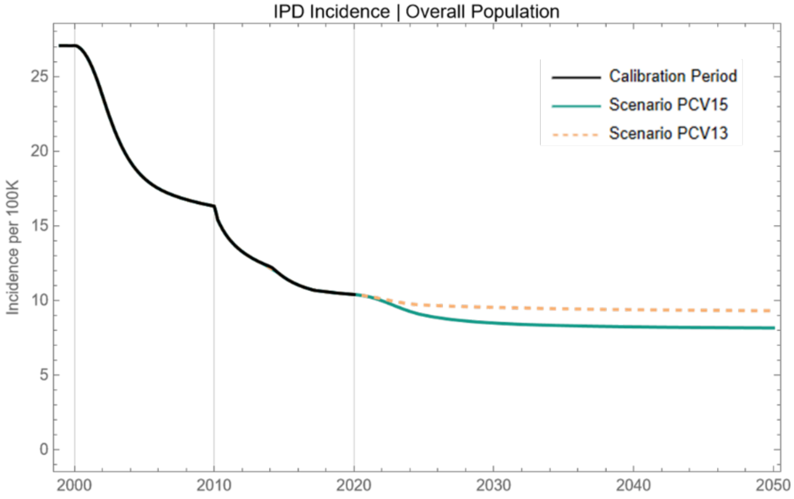

Figure S17. Calibrated and projected IPD incidence per 100,000 in the US population.

a) PCV13 STs, children < 5

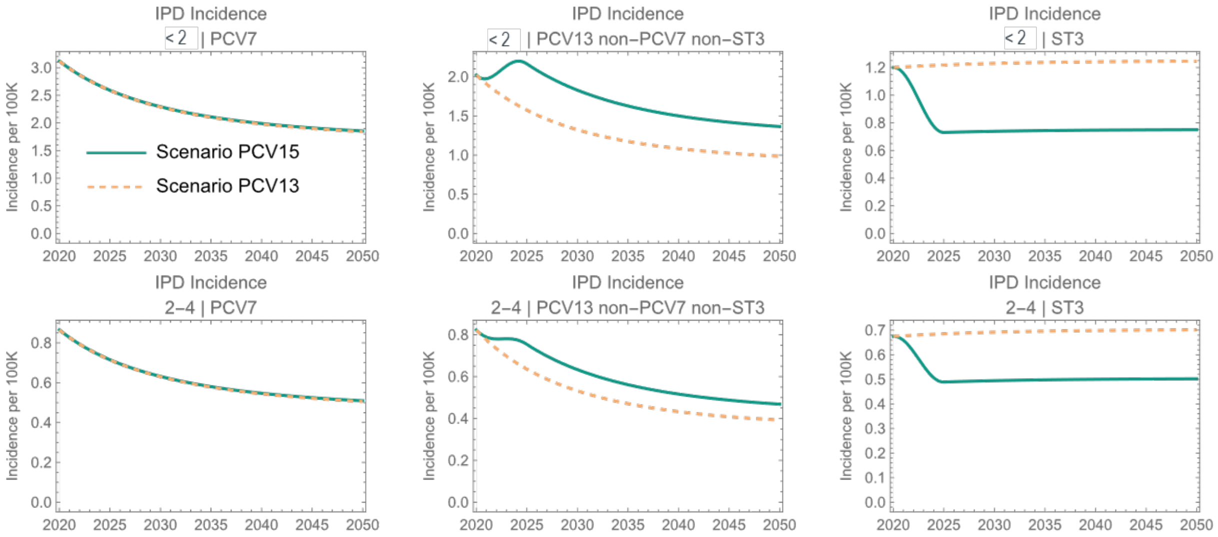

(b) non-PCV13 STs, children < 5

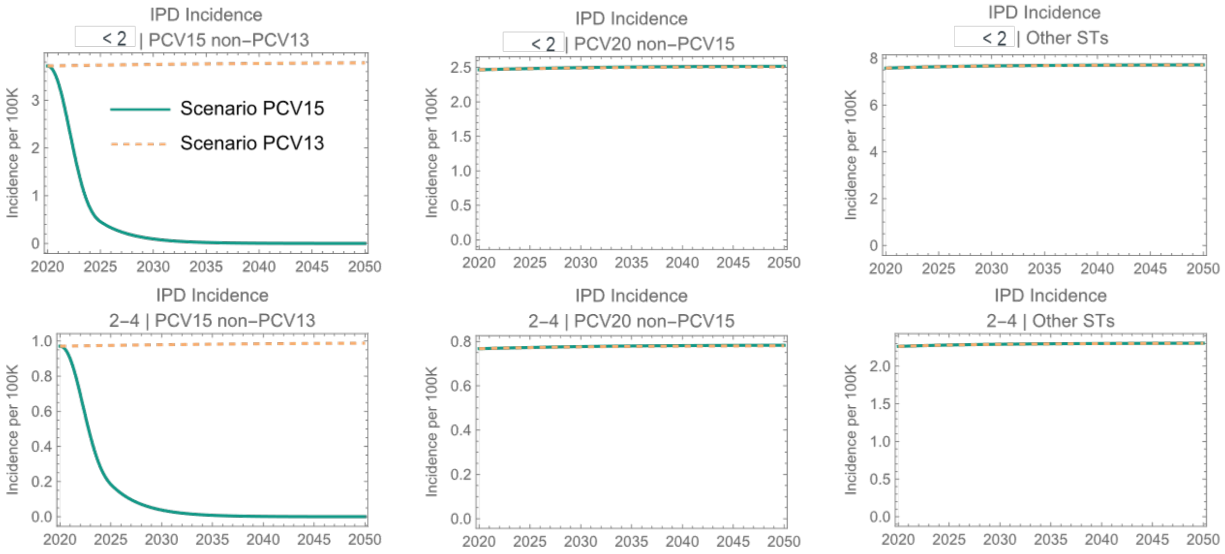

(c) Overall IPD burden in children < 5

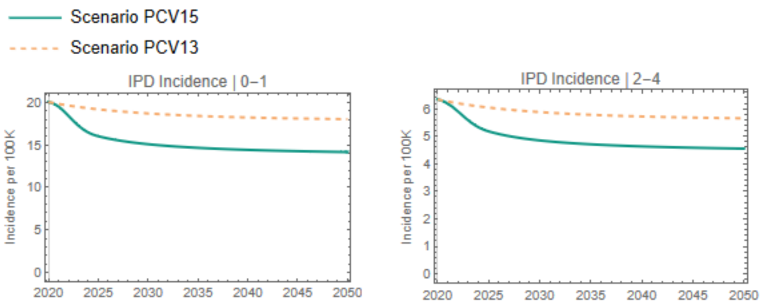

Figure S18. IPD projections.

PCV15 reduces a greater number of IPD cases due to STC3 and STC6 (Figure S18(a) and (b)). It has lower VEd for STC 2, 4 and 5, therefore more cases of IPD due to PCV13 non-PCV7 non-ST3 are attributed to PCV15. However, there is a net greater reduction of IPD with PCV15 compared to PCV13 (13.2% over 10 years in children under 2 and 11.47% in children 2-4 years old) (Figure S18(c)). Indirect protection also leads to about 4.8% more reduction in IPD among 65+.

#### 7.2 NBPP projections

(a) NBPP burden, children < 5

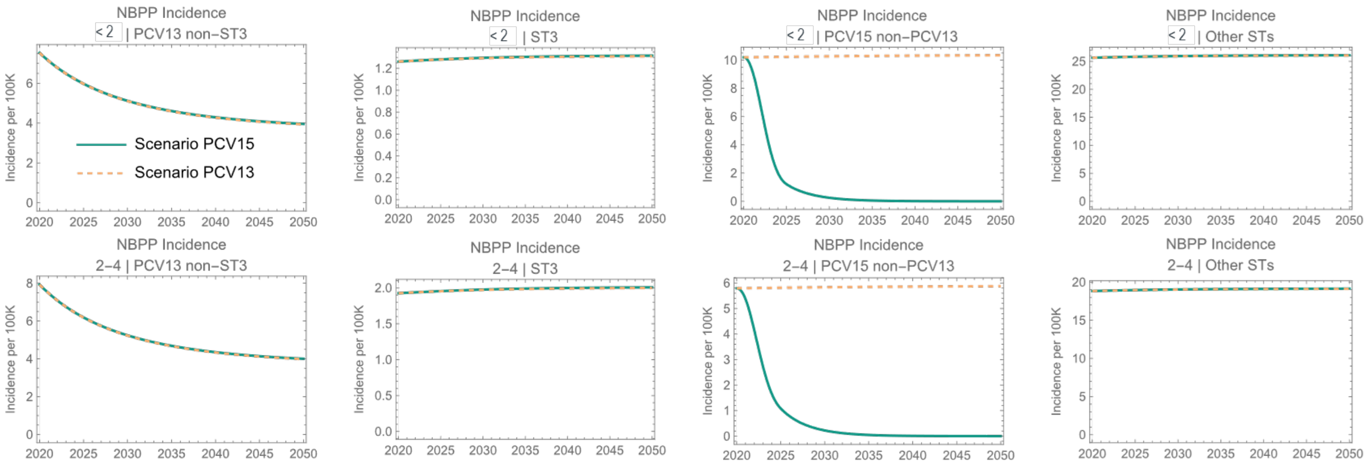

(b) NBPP burden, 65+

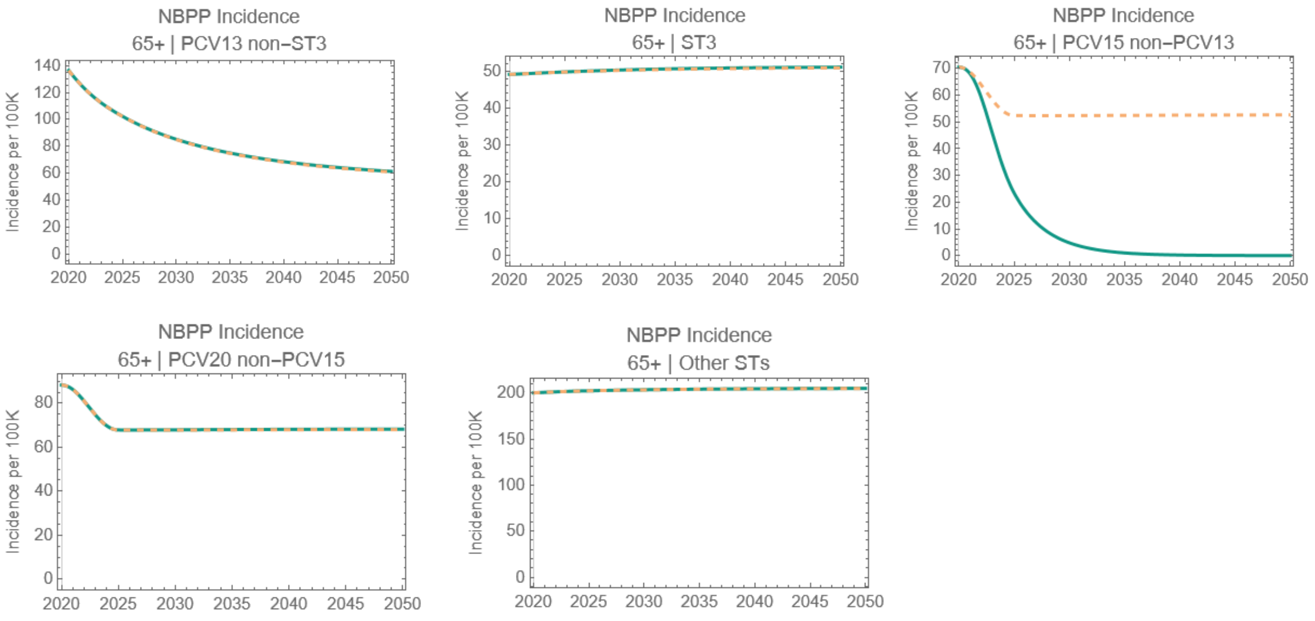

Figure S19. NBPP incidence (per 100,000) projected by the calibrated model. Solid line denotes Scenario 15 (i.e., PCV15 in children < 2) and dashed line denotes Scenario PCV13 (i.e., PCV13 in children < 2). Adult vaccination is identical in both scenarios: 80% receive PCV20, 10% receive PPSV23, and 10% receive PCV15+PPSV23.

Over 10 years, PCV15 prevents around 16.5% more cases of NBPP than PCV13 in children under 2, 11.5% more cases in 2–4-year-old children, 7.37% more cases in 5-17, around 5.49% more cases in 18-49, 5.43% more cases in 50-64 and 5.22% more cases of NBPP in 65+ (Figure S19).

#### 7.3 AOM projections

AOM burden, children < 5

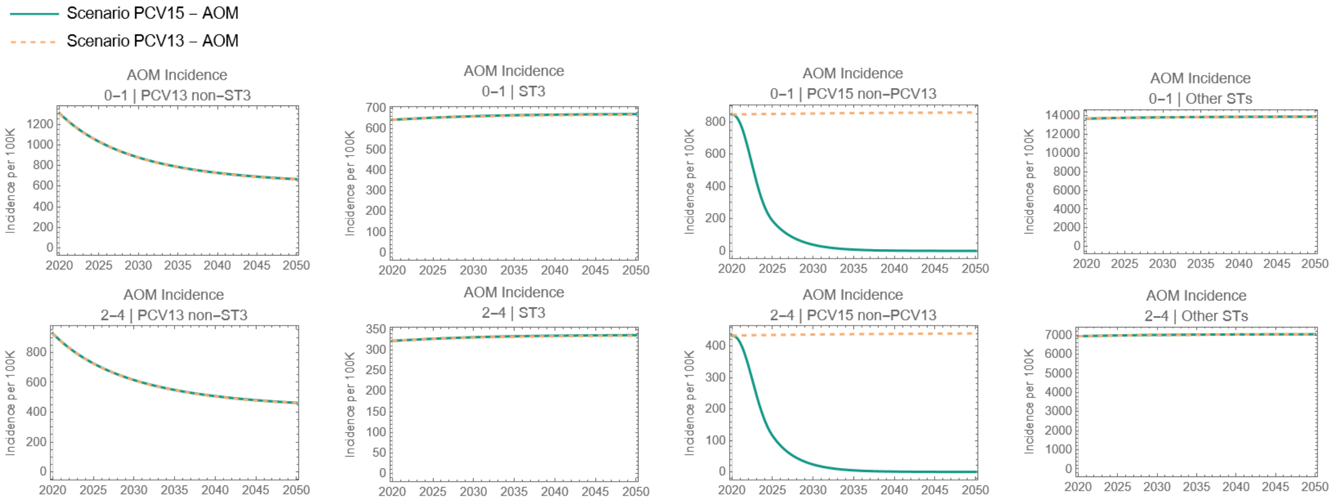

Figure S20. AOM incidence (per 100,000) projected by the calibrated model. Solid line denotes Scenario 15 (i.e., PCV15 in children < 2) and dashed line denotes Scenario PCV13 (i.e., PCV13 in children < 2).

Over 10 years, PCV15 prevents about 3.26% more cases of AOM in < 2 children, 3.07% more cases in 2-4 and 2.58% more cases of AOM in 5–17-year-olds compared to PCV13 (Figure S20).

[9] S. Snedecor, D. Strutton, V. Ciuryla, E. Schwartz and M. Botteman, "Transmission-dynamic model to capture the indirect effects of infant vaccination with Prevnar (7-valent

pneumococcal conjugate vaccine (PCV7)) in older populations," *Vaccine,* vol. 27, no. 34, pp. 4694-703, 2009.

[17] S. Flasche, J. Ojal, O. L. P. d. Waroux, M. Otiende, K. L. O'Brien, M. Kiti, D. J. Nokes, W. J. Edmunds and J. A. G. Scott, "Assessing the efficiency of catch-up campaigns for the

introduction of pneumococcal conjugate vaccine: a modelling study based on data from PCV10 introduction in Kilifi, Kenya," *MBC Med,* vol. 15, no. 113, 2017.

[37] A. Parker, J. N. A. S. K. H. T. Alwan, A. Wyllie, K. Kogut, N. M. A. Holland, B. Eskenazi, L. Riley and J. Lewnard, "Predictors of upper respiratory <em>Streptococcus pneumoniae</em> colonization among working-age adults with prevalent exposure to overcrowding.," *medRxiv,* 2024.

[38] H. Rinta-Kokko, R. Dagan, N. Givon-Lavi and K. Auranen, "Estimation of vaccine efficacy against acquisition of pneumococcal carriage.," *Vaccine,* vol. 27, no. 29, pp. 3831-7, 2009.

[39] P. C. Hill, Y. B. Cheung, A. Akisanya, K. Sankareh, G. Lahai, B. M. Greenwood and R. A. Adegbola, "Nasopharyngeal carriage of Streptococcus pneumoniae in Gambian infants: a longitudinal study.," *Clin Infect Dis,* vol. 46, no. 6, pp. 807-14, 2008.

[40] C. Chaguza, M. Senghore, E. Bojang, S. W. Lo, C. Ebruke, R. A. Gladstone, P.-E. Tientcheu, R. E. Bancroft, A. Worwui, E. Foster-Nyarko, F. Ceesay and C. Okoi, "Carriage Dynamics of Pneumococcal Serotypes in Naturally Colonized Infants in a Rural African Setting During the First Year of Life," *Front Pediatr,* vol. 8, p. 587730, 2021.

[41] O. Abdullahi, A. Karani, C. C. Tigoi, D. Mugo, S. Kungu, E. Wanjiru, J. Jomo, R. Musyimi, M. Lipsitch and J. A. G. Scott, "Rates of acquisition and clearance of pneumococcal serotypes in the nasopharynges of children in Kilifi District, Kenya," *J Infect Dis,* vol. 206, no. 7, pp. 1020-9, 2012.

[42] O. Abdullahi, A. Karani, C. C. Tigoi, D. Mugo, S. Kungu, E. Wanjiru, J. Jomo, R. Musyimi, M. Lipsitch and J. A. G. Scott, "The prevalence and risk factors for pneumococcal

colonization of the nasopharynx among children in Kilifi District, Kenya," *PLoS One,* vol. 7, no. 2, p. e30787, 2012.

[49] K. Auranen, J. Mehtala, A. Tanskanen and M. S. Kaltoft, "Between-strain competition in acquisition and clearance of pneumococcal carriage--epidemiologic evidence from a

longitudinal study of day-care children.," *Am J Epidemiol,* vol. 171, no. 2, pp. 169-76, 2009.

https://www.cdc.gov/vaccines/imz-managers/coverage/adultvaxview/pubs-resources/pcv13-medicare-beneficiaries-2010-2019.html. [Accessed 11 04 2024].

[84] L. R. Grant, L. L. Hammitt, S. E. O'Brien, M. R. Jacobs, C. Donaldson, R. C. Weatherholtz, R. Reid, M. Santosham and K. L. O'Brien, "Impact of the 13-Valent Pneumococcal

Conjugate Vaccine on Pneumococcal Carriage Among American Indians," *Pediatr Infect Dis J,* vol. 35, no. 8, pp. 907-914, 2017.

[91] R. E. Isturiz, J. Ramirez, W. H. Self, C. G. Grijalva, F. L. Counselman, G. Volturo, L. Ostrosky-Zeichner, P. Peyrani, R. G. Wunderink, R. Sherwin, J. S. Overcash, S. P. Oliva

and T. File, "Pneumococcal epidemiology among us adults hospitalized for community-acquired pneumonia," *Vaccine,* vol. 37, no. 25, pp. 3352-61, 2019.

[98] N. J. Andrews, P. A. Waight, R. C. George, M. P. Slack and E. Miller, "Impact and effectiveness of 23-valent pneumococcal polysaccharide vaccine against invasive

pneumococcal disease in the elderly in England and Wales," *Vaccine,* vol. 30, pp. 6802-6808, 2012.
